# A Systematic Scoping Review and Conceptual Analysis of New-Onset Fibromyalgia Manifestations After Non-Hospitalized COVID-19: Empirics, Definitions, Methodologies, Pathophysiology, Mapping of Literature, and Knowledge Gaps

**DOI:** 10.1101/2025.10.23.25338705

**Authors:** Shiloh Plaut

## Abstract

The global coronavirus pandemic has led to a quiet wave of a chronic illness referred to as ‘Long/Post Covid-19 syndrome’ (LC) which bears a notable resemblance to functional somatic or ‘fibromyalgia-type’ syndromes, and whose pathophysiology is undetermined. The lack of effective therapies for LC is straining healthcare systems worldwide and causing widespread public health and socioeconomic concerns. “Fibromyalgia” is a controversial chronic pain condition of unknown etiology largely attributed to generalized sensory hypersensitivity due to dysregulated central pain processing pathways (i.e., central sensitization). Despite intense research and growing attention in the scientific community, the clinical overlap of fibromyalgia, somatic symptom disorder, and post-viral chronic fatigue, is a medical puzzle yet to be solved, especially when occurring in non-severe infections and previously healthy individuals. This systematic scoping review covers the empirical findings on new-onset fibromyalgia manifestations after non-hospitalized covid-19. MEDLINE, Web of Science, and APA PsycINFO were searched in a systematic scoping approach for empirical studies on new-onset fibromyalgia after non-severe non-hospitalized covid-19, charting study characteristics and outcome data. A total of 228 records were included. Various types of methods, tools, and study designs are being used for LC research, with inconsistency in key concepts and definitions. This leads to a fragmented understanding of the relationship between SARS-CoV-2 infection and LC. Prevalence studies of post-Covid fibromyalgia are ongoing and susceptible to bias. The empirical evidence supports an overlap between LC, chronic fatigue syndrome, and fibromyalgia but the molecular mechanisms still remain unclear. There are conflicting findings regarding presence of viral particles, central sensitization, autoantibodies, and more. This review highlights the need for standardized definitions and rigorous methodologies in research on LC. Future research should focus on epidemiological population-based studies with representative sampling and improving methodology, refining evolving definitions, harmonization of research, elucidating neurological mechanisms in hypothesis driven studies, and developing effective therapeutic strategies. The discussion synthesizes findings and offers an integrative mechanism for the pathophysiology of fibromyalgia and multisystem medically unexplained manifestations of LC as a non-autoimmune connective tissue disease and is used to make testable theory-based predictions for future investigations.

## 1. Introduction

As the aftermath of the coronavirus pandemic continues to unravel, many convalescent patients have remained with long-term multi-symptom illness following infection with severe acute respiratory syndrome coronavirus 2 (SARS-CoV-2) and coronavirus disease 2019 (COVID-19) (1). Terms such as ‘long COVID-19’ (LC), post-acute sequelae of COVID-19 (PASC), and post-COVID-19 condition (PCC) are used somewhat interchangeably in the literature to describe the persistent symptoms and sequelae that can last for weeks, months, and even years, following the acute phase of SARS-CoV-2 infection. Although it had initially received less attention, LC is lately becoming recognized as a global public health challenge (1–4), to such an extent that the National Institute of Health (NIH) allocated more than 1 billion dollars for LC research in the year 2021 (5). The incidence of “post-acute COVID-19 condition” is estimated to be approximately 10–35% of individuals positive for SARS-CoV-2 and is said to occur more frequently in COVID-19 cases that involved hospitalization (2,4,6–8) but these estimates vary with the timeframe of data collected since initial acute COVID-19 and the definitions used. Despite ongoing extensive investigations and lots of speculations, the pathophysiological mechanisms of the post-acute sequelae of COVID-19 are poorly understood, thus impeding the development of effective treatments. Interestingly, the medically unexplained multisite symptoms of LC have symptomatologic overlap and a surprising resemblance to functional psychosomatic syndromes such as myalgic encephalomyelitis/chronic fatigue syndrome (ME/CFS) and fibromyalgia and may even share a similar pathophysiology (9–14). Post-viral infection syndromes are already known to be characterized by persistent disabling fatigue, arthralgia/myalgia, neurocognitive difficulties, and mood disturbances (15).

New-onset fibromyalgia syndrome is being identified as a prevalent condition in convalescent individuals following acute COVID-19 (16,17). According to a study in Sweden, many of convalescent COVID-19 patients who initially only had mild SARS-CoV-2 infection and were previously healthy that are affected by LC have generalized chronic pain and fatigue and fulfill the 2016 diagnostic criteria for “fibromyalgia” (16). What’s more, LC patients often fulfill the diagnostic criteria for ME/CFS (18). Many individuals who experience LC exhibit persistent pain despite having experienced mild initial infection, therefore effective pain management strategies in LC syndrome are needed (16,19). In this paper findings are presented from a systematic scoping review covering the empirical findings on new-onset fibromyalgia after non-hospitalized Covid-19, followed with a synthesis of data for offering a pathophysiological mechanism for the syndrome of fibromyalgia.

“Fibromyalgia syndrome” (20) is a heterogenous chronic pain condition of unknown etiology characterized by widespread musculoskeletal pain, post-exertional malaise, fatigue, and cognitive difficulties, and considerably overlaps with ME/CFS whose predominant feature is relentless fatigue while musculoskeletal pain is implied in its name (21). Fibromyalgia, which lies within the spectrum of medically unexplained symptoms (22,23) (sometimes regarded as a “non-disease”) and whose chronic pain is frequently said to be unattributable to any identifiable organic pathology (24), has an estimated prevalence of ∼1-6 percent of the general population and leads to a significant burden on the healthcare system and considerably impacts patients’ quality of life and emotional health (20,25–27).

Despite extensive research in the recent decades (28), the patho-mechanisms underlying fibromyalgia are still disputed and poorly elucidated (29) and the field remains in relative stagnation in terms of translation to therapeutic clinical impact, in what can be termed “a huge unmet medical need” (30). After having been regarded as a connective tissue disorder in the nineteenth and twentieth centuries (31), that idea has since then been abandoned and the most accepted and widely investigated theory nowadays for the pain of fibromyalgia is central sensitization (i.e., a dysfunction of ascending and descending somatosensory processing neural pathways in the spinal cord and brain, facilitated by structural and functional alterations in the central nervous system or a “nociplastic” malfunction) although, there are some authors who question this thesis (14,32–37). The international association for the study of pain (IASP) explains that it’s a pain disorder of its own, not a symptom of any other underlying organic disease (38). Remarkably, in a cross-sectional survey of Canadian Rheumatologists, 30 percent of them asserted that fibromyalgia was a psychosocial condition (39). Even though the pathophysiology of fibromyalgia is still under much dispute by researchers and practitioners in the field (16,40), the therapeutic strategy for fibromyalgia is usually derived from this theory of “central sensitization” (32,41–44) and is generally considered by clinicians to be ineffective (30,33,45–47).

As with fibromyalgia, results in medical tests that are offered as standard care are often unremarkable in patients with LC (8,12,48,49) and some authors nowadays describe a subset of patients as having “functional long-COVID” or “ME/CFS subtype of long COVID” or a “central sensitization phenotype” in cases that present with subjective complaints and functional impairment yet no apparent organ damage (11,50–52). Some authors argue that LC is likely a disorder of the domain of psychology and psychosomatics (30,53).

This manuscript is divided into two parts: Part 1 presents the findings from the systematic scoping literature review and the empirical evidence on new-onset fibromyalgia after non-hospitalized SARS-CoV-2 infection. The motivation behind this review was to find out what the evidence is on new-onset fibromyalgia manifestations post-Covid-19, and the goal was to clarify key concepts and definitions, map existing evidence, identify gaps in knowledge, and report on the extent and types of evidence, the methods being used to research it, and report on the methodological consistency or inconsistency across studies in this new emerging field of research. For this reason, a scoping-type review was conducted. In Part 2, building on a synthesis of data, a connective-tissue-based theoretical model is presented for the pathophysiology of fibromyalgia syndrome. The objective is to apply this model to fibromyalgia features of LC and reconcile the findings and anomalies encountered in part 1. The model depicts a neuromechanobiological disorder of the musculoskeletal system driven by the cascade of myofibroblast extracellular matrix remodeling and their natural tensile force generation in soft tissue, which drive peripheral and central pain mechanisms. Implications and predictions of this model are discussed.

## 2. Methods

The review follows recommendations of the Preferred Reporting Items for Systematic Reviews and Meta-Analysis Extension for Scoping Reviews (PRISMA-ScR) (54).

### 2.1. Search strategy

A systematic search was conducted using key phrases on fibromyalgia and covid-19 in Web of Science, MEDLINE, and PsycInfo, in all fields for all study types since inception until December 31, 2024. Detailed documentation of the search phrases that were used for each database can be found in the supplementary material. An example of the search phrase used in MEDLINE (via PubMed) is as follows: ((“fibromyalgi*” OR “myofascial pain*” OR “central sensitization*” OR “central sensitisation” OR “nociplastic pain”) OR (“Fibromyalgia”[Mesh] OR “Central Nervous System Sensitization”[Mesh] OR “Myofascial Pain Syndromes”[Mesh])) AND ((“covid*” OR “coronavirus” OR “sars-cov-2”) OR (“COVID-19”[Mesh] OR “Coronavirus”[Mesh] OR “SARS-CoV-2”[Mesh])).

Inclusion criteria: A record was to be included if its full text was accessible for review, and related to new-onset fibromyalgia features after sars-cov-2 infection (confirmed, self-reported, or suspected), in individuals who did not have fibromyalgia prior to infection, in the absence of a reported clear pathology or well-defined organ damage to account for symptoms after non-severe non-hospitalized Covid-19. Specifically, this included primarily studies related (but not limited) to epidemiology, symptoms, pathophysiology, disease course, patient surveys and experiences, diagnostics, and interventions, as well as studies reporting empirical evidence in the context of a theoretical mechanistic link to the putative pathophysiology of fibromyalgia. “Fibromyalgia features” were defined for this purpose as either clinical diagnosis of fibromyalgia, fulfilling criteria for fibromyalgia, stated suspicion of fibromyalgia, positive screening for fibromyalgia, new onset widespread musculoskeletal or “non-specific”/myofascial pain in the absence of a clear pathology or well-defined organ damage to account for it after non-severe non-hospitalized Covid-19. Empirical studies (e.g., clinical trials, qualitative or quantitative observational studies, case reports, etc.) both basic/translational research and clinical, and reviews (topical, narrative, systematic, meta-analyses) to be eligible.

Various terms are being used in literature to refer generally to long-lasting symptoms after COVID-19, e.g., long-haul COVID-19, chronic COVID, long COVID, post COVID, post-acute COVID-19, etc. The a-priori Long COVID-19 definition for the review’s purpose is described in the supplementary material, taking a similar approach to Phillips & Williams (2021) (8) that assert that *“long covid is not a condition for which there are currently accepted objective diagnostic tests or biomarkers, i.e., it is not blood clots, myocarditis, multisystem inflammatory disease, pneumonia, or any number of well-characterized conditions caused by covid-19.”*

Exclusion criteria: Studies published in journals whose highest ranking was Q4 in their specialty field(s) in the year of the record’s publication according to the journal citation reports (JCR) (using the Journal Citation Indicator ranking) were excluded. For papers published during the year 2024 whose ranking was unavailable in JCR, the quartile given in the previous year was taken. Studies published in journals not listed in JCR were excluded from the systematic part. Non-English language records were excluded.

After title and abstract read, items deemed either “relevant” or “possibly relevant” (i.e., not irrelevant or off-topic) based on title and abstract underwent a full text inspection for final inclusion or exclusion based on the abovementioned criteria. During the full text read of an included paper, the reference list was also inspected to identify additional records on post-Covid fibromyalgia that were potentially eligible for inclusion.

### 2.2. Data charting

For the included records, data abstracting and charting was conducted on a charting form for the documentation of essential information from each record (title, first author, publication date, journal, type of study, aims of the study, summary of study design and methods, population characteristic if relevant, main findings, main theme(s), JCR quartile for peer-reviewed publications). Each article was tagged by its focus according to its main theme(s) developed during the review of the literature.

### 2.3. Additional non-systematic searches for subtopics and preprints

Additional literature was added as part of the scoping search from pubmed and web of science and from the preprint database medRxiv. This part was not done systematically and included subtopics such as post-covid, myofascial tissue in long covid, fascia, myofibroblasts and fibrosis in post-covid-19, acupuncture in long covid, lifestyle and fibromyalgia-type syndromes, medically unexplained symptoms after covid-19, long/post covid treatment, chronic fatigue syndrome in LC (due to the significant clinical overlap with fibromyalgia), and acupuncture in chronic fatigue syndrome, and other subtopics identified during the review process such as joint hypermobility syndrome. The literature from this non-systematic narrative part will be presented separately in the findings section. Figure 1 shows a diagram summarizing the scoping review process.

**Figure 1:**
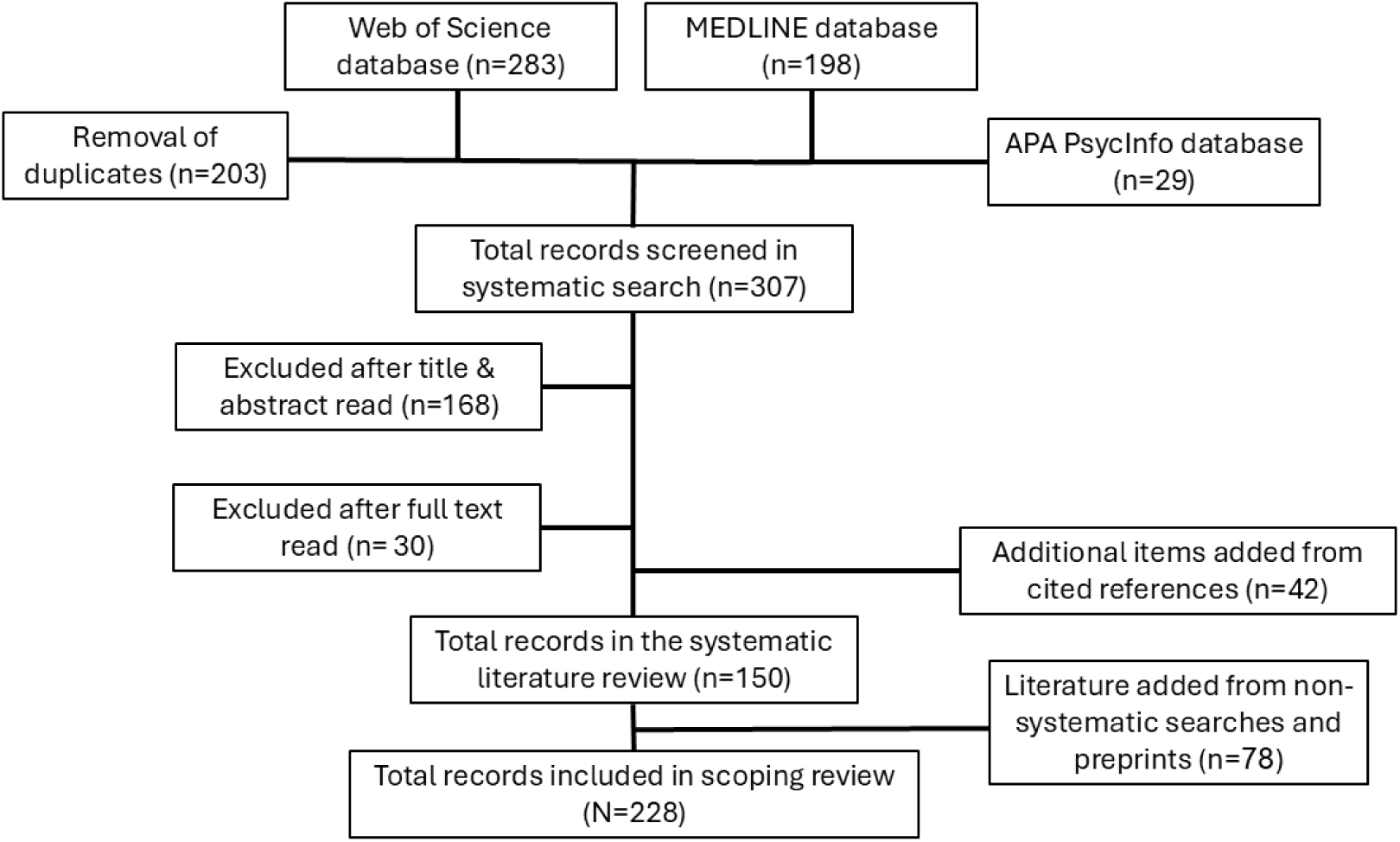
Schematic outline of the systematic scoping review process.

## 3. Part one-findings from the systematic scoping review

After removal of duplicates, 307 records were initially screened, afterwards 198 were excluded based on title/abstract or full text read (of them three records were excluded because of no access to their full text). 42 records were added from the cited reference list of included records. 150 records were included in the systematic literature review, and an additional 78 from non-systematic searches, for a total of 228 records finally included in the scoping review.

Records whose main topic was ME/CFS even without involving fibromyalgia were included due to the close symptomatologic overlap (55) between the two syndromes. Whenever there was ambiguity whether the sampled population in a study was hospitalized or not, the record was included in the review. If both hospitalized and non-hospitalized patients were recruited in a study, the record was included. Also, non-empirical studies on fibromyalgia and covid (such as opinion, viewpoints, speculations and hypotheses, etc.) were included. Few older studies on sars-cov-1 were found but none fit the inclusion criteria.

Common themes were (a single record may be allocated to more than one theme):

- Chronic fatigue or chronic fatigue syndrome after viral infection (49,55–71)
- Putative LC relevant patho-mechanisms (11,13,14,21,24,36,43,51,55,58,61,67,72–101)
- Chronic pain after sars-cov-2 infection or during the covid pandemic (30,49,68,102–124)
- Observational studies of fibromyalgia syndrome after COVID-19 (10,16,48,52,104,114,125–134)
- Overlap between fibromyalgia and/or chronic fatigue syndrome and/or LC (13,21,43,51,53,55,64,66,71,86,123,124,128,132,135–141)
- Speculation on emotional/psychosocial stress as a trigger of fibromyalgia post-COVID-19 (21,53,102,105,124,142,143).
- Speculations or empirical investigations on possible central sensitization in LC (52,53,72,77,92,103,104,110–112,117,137,139,142–148)
- Hypothetical and speculative papers on potential treatments for LC (14,56,58,109,124,147,149,150)
- Interventions in LC or post-viral infection fatigue (68,137,151–161)
- SARS-CoV-2 and myofascial-type pain (19,121,127,145,162,163)
- Other (e.g., study protocols, post-COVID manifestations, LC phenotyping, rheumatoid arthritis, hypermobility syndrome) (2,8,65,68,87,126,135,136,138,140,141,164–186).

The distribution of publication ranking according to JCR quartile (ranging from Q1 to Q3) was as follows: Q1-80/148 (54%), Q2-54/148 (36%), Q3-14/148 (9%). Two additional records were included: a clinical guideline (186) and the full paper of a study that was presented as a meeting abstract (183).

The records were of the following types:

- Observational human studies (excluding case series, case reports, and conference abstracts) (n=60) (2,10,16,48,49,52,61,64,65,67–69,75,77,81,85,88,91,93,99–101,104,105,108,111–115,118,121,127–130,132–137,140,141,144–146,148,154,155,163,167,170,173,177–179,182,183,185)
- Case reports or case series (n=6) (19,126,160–162,171)
- Interventional studies (n=8) (89,150–153,156,157,159)
- Non-systematic reviews (including narrative reviews, speculative reviews, and topical articles) (n=37) (11,13,14,21,36,51,55–58,72–74,76,79,80,82–84,90,92,95,96,102,103,109,110,117,138,142,147,149,166,176,180,181,184)
- Systematic reviews (with or without meta-analysis) (n=12) (24,59,60,62,70,94,106,107,116,124,143,158)
- Comments, editorials, viewpoints, and letters (n=14) (8,30,43,53,66,78,86,87,97,98,119,139,169,174)
- Congress/meeting abstracts (n=8) (63,71,120,122,123,125,131,175)
- Official clinical guidelines (n=1) (186)
- Other (descriptive papers, study protocols) (n=4) (164,165,168,172)

Figure 2 shows the distribution of record types included in the systematic part.

**Figure 2:**
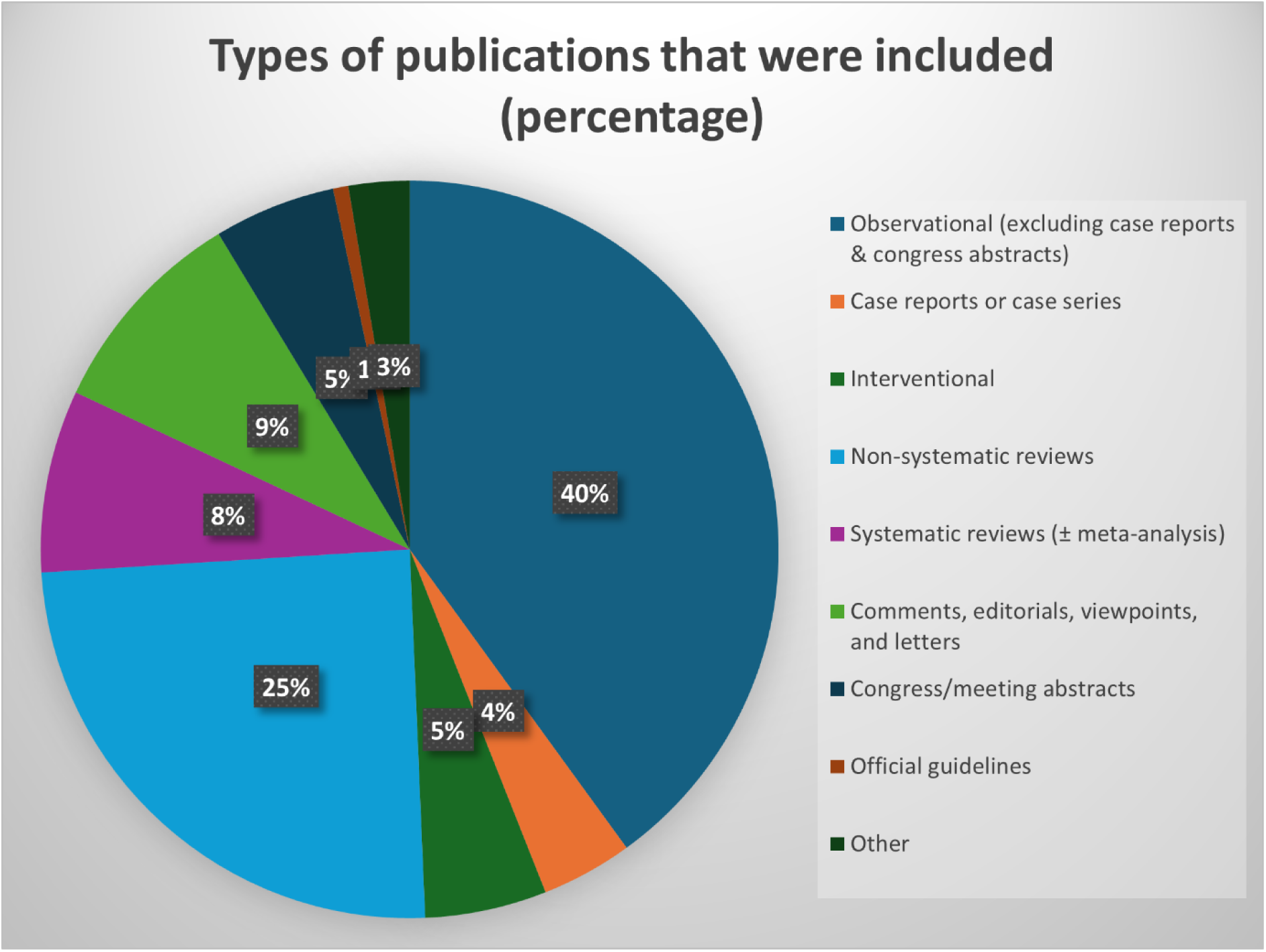
Distribution of records according to types of publications.

A brief description of types of records excluded e.g., (17,187–206), is provided in the supplementary material.

The following sections present the main findings of the review.

### 3.1. Definitions, Research Inclusion/Exclusion Criteria in LC studies, and Measurement Tools

#### 3.1.1. Definitions & Research Inclusion/Exclusion Criteria

An analysis of the methodologies employed in LC studies reveals significant heterogeneity in LC definitions and the inclusion and exclusion criteria for LC research including LC patients, individuals with a prior SARS-CoV-2 infection, and control subjects. This variability underscores the evolving understanding of LC and the challenges that stem from studying such a novel and complex condition. Various approaches to inclusion and exclusion criteria were found in studies as follows:

- <u>COVID-19 individuals</u>: COVID-19 or SARS-CoV-2 infected individuals were seen defined differently across studies and involved different inclusion and exclusion criteria. A “COVID-19” individual was established either by self-report of the participant (e.g., based on symptoms consistent with COVID-19, a self-reported physician diagnosis of COVID-19, or self reported positive COVID-19 test), positive immunoglobulin response, a documented positive test in healthcare databases or COVID registries (e.g., PCR test), or by logical combinations of these conditions, for example. To give a specific example, in a study by Peterson et al. (75), symptomatic COVID-19 individuals were those who self-reported the symptoms that they had experienced during active COVID-19 infection from a list modified from the CDC and provided evidence of a previous positive PCR or antibody ELISA test indicating infection. On the other hand, the asymptomatic COVID-19 group consisted of those participants who self-reported no symptoms and had a previous positive PCR and/or positive antibody test (or self-reported that they had no symptoms but had a positive antibody test).

In case of a non-LC (i.e., recovered COVID-19) infected individual, absence of persistent symptoms beyond a certain timeframe or the absence of symptoms altogether defined the convalescent group (91,185). Such participants may have undergone a brief verbal screening to confirm no active symptomatology.

Severity of acute illness: few studies stratified the COVID-19 group based on the severity of their acute COVID-19 illness (e.g., hospitalized vs. non-hospitalized).

Additionally, unclear onset of symptoms was sometimes used as an exclusion criteria for self-reported COVID-19 (153).

- <u>Controls and Healthy controls:</u> were also seen defined differently across studies, depending on the study (81,91,104,115). The heterogeneity in defining these crucial comparator groups has significant implications for interpreting research findings and understanding the true impact of SARS-CoV-2 infection. Healthy uninfected controls were often defined as individuals who have no prior history of COVID-19 infection, often confirmed through PCR and antibody testing, while other studies defined control groups as those with no active symptomatology or implementing both conditions (91). Naturally, the longer the duration into the pandemic the more difficult it would have been for investigators to find non-infected individuals. Damasceno et al. (2023) (115) chose adults who had COVID-19 for at least 3 months prior to the data collection and without a chronic pain syndrome. Examples of inclusion criteria are (i) individuals that reported they did not have a confirmed objective COVID-19 test (e.g., PCR or home kit) (127), (ii) individuals that do not have a previous history of medical conditions as self-reported, (iii) no previous symptoms self-reportedly and negative result on the PCR and antibody test (75,118), and (iv) based on (absence of) diagnoses in medical records or healthcare registries. The non-infection healthy control group in Peterson et al., for example, were those who self-reported no previous symptoms and were negative on the PCR and antibody test administered immediately prior to carrying out the study’s investigation (75).
- <u>Long COVID, Post-COVID condition, Persistent COVID symptoms, and other parallel terms:</u> A fundamental aspect of LC research is the definition used to identify affected individuals. Various studies employ different criteria, often aligning with guidelines from organizations like NICE and WHO, or developing their own definitions.

A consistent inclusion criterion across many studies was, naturally, the persistence of symptoms for a defined duration following the acute phase of SARS-CoV-2 infection (e.g., 4, 6, 12 weeks). Another additional inclusion condition often used for LC was confirmation of prior SARS-CoV-2 infection (by positive PCR, serology, self-reportedly, independent clinician, rapid antigen test with documentary proof from a health authority (159), or documentation in electronic health records). Some LC studies included only previously healthy individuals. Self-reported history of confirmed or probable COVID-19 infection according to WHO guidelines was also seen integrated into the inclusion criteria (91). Certain studies focused on individuals experiencing a particular set of new persistent symptoms after acute COVID-19 (133), such as musculoskeletal pain (114) or neurological symptoms (182) whereas others focused on evident reduction in the level of functioning and activity or participation in daily life compared to before the infection (16). Exclusion of alternative etiologies: some studies incorporated a process to rule out alternative medical etiologies for persistent symptoms, such as medical evaluations by physicians, or self-reportedly. Pre-existing chronic pain prior to COVID-19 infection, pre-existing chronic fatigue syndrome or fibromyalgia were sometimes part of the exclusion criteria (155).

Examples of inclusion criteria for Post COVID/Long COVID/Chronic COVID/Subacute COVID/“persistent symptoms” or “non-recovery from COVID”: (i) participant self-reporting not to have been fully recovered after COVID-19 (146), (ii) participant self-reported physician-made diagnosis of LC (150), (iii) self-reported physician made diagnosis combined with a previous positive COVID-19 test (128), (iv) persistent symptoms beyond a specified interval of time (e.g., 12 weeks) (141,165), (v) presence of any persistent symptom since SARS-CoV-2 infection (or any persistent symptom among a predetermined list of symptoms) (93,99,185), (vi) persistent symptoms and negative Covid test for excluding active infection (75,154), (vii) based on the world health organization’s consensus definition (101), (viii) Bierle et al.’s (2021) criteria (144), (ix) fulfilling the official 2015 diagnostic criteria for ME/CFS (155), (x) persistent post-exertional malaise for 3 or more months verified by the DePaul Symptom Questionnaire (89), (xi) referral to-or diagnosis by-a post-Covid clinic, and more (137,156).

A reader would notice correctly that some of the above examples can conflate “LC syndrome” and “persistent COVID symptoms,” which are not necessarily the same. Noteworthy, as opposed to simply including persistent symptoms, in case a syndrome is what investigators are aiming to investigate, defining LC for the purpose of a study as at least one persistent symptom, any symptom, even hyposmia, does not necessarily reflect a syndrome, in agreement with Phillips and Williams (8). Lau et al. (2024) (159), for example, included individuals fulfilling the Centers for Disease Control and Prevention criteria for post-acute COVID condition and at least one of 14 symptoms included in their post-acute COVID-19 syndrome 14-item improvement questionnaire (PACSQ-14) for four weeks or more after SARS-CoV-2 infection. Matta et al. (2022) (185), in their widely cited publication of whether belief in having had COVID-19 and actually having had the infection (when verified by SARS-CoV-2 serology testing) were associated with persistent physical symptoms after COVID-19, in the context of LC, included individuals with at least one persistent symptom among a list of symptoms present for the past 4 weeks and lasting more than 8 weeks. That list consisted of headache, back pain, joint pain, muscular pain, sore muscles, sleep problems, fatigue, sensory symptoms such as pins and needles, tingling or burning sensation, skin problems, poor attention or concentration, hearing impairment, stomach pain, constipation, breathing difficulties, palpitations, chest pain, dizziness, cough, diarrhea, anosmia, and other symptoms.

Eccles et al. (2024) (170) determined non-recovery from COVID-19 from a dichotomous self-reported response to the question “Thinking about the last or only episode of COVID-19 you have had, have you now recovered and are back to normal?” while Amsterdam et al. (2024) (133) recruited outpatients from a post-COVID clinic who, subsequent to non-hospitalized COVID-19, developed a prolonged illness, leading to a diagnosis of LC syndrome characterized by the persistence of one or more symptoms for over a month: dyspnea, cough, cognitive decline, brain fog, or fatigue, going by reference to the 2020 published NICE guidelines. Azcue et al. (141) took a similar approach and explicitly excluded respiratory symptoms persisting for 12 weeks post-infection, severe bilateral pneumonia, admission to an intensive care unit, or other manifestations necessitating hospitalization.

The Centers for Disease Control and the National Academies of Sciences, Engineering and Medicine offer their definitions for LC terminologies (80). Table 1 summarizes, in a non-exhaustive list, official definitions according to several national and international health bodies.

**Table 1:**
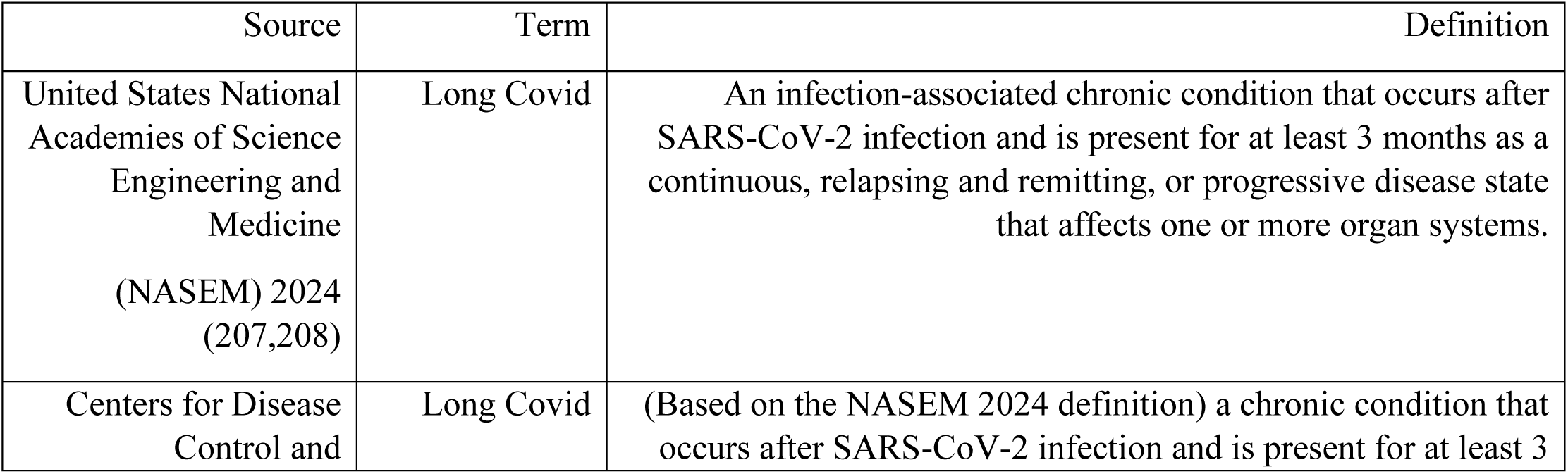
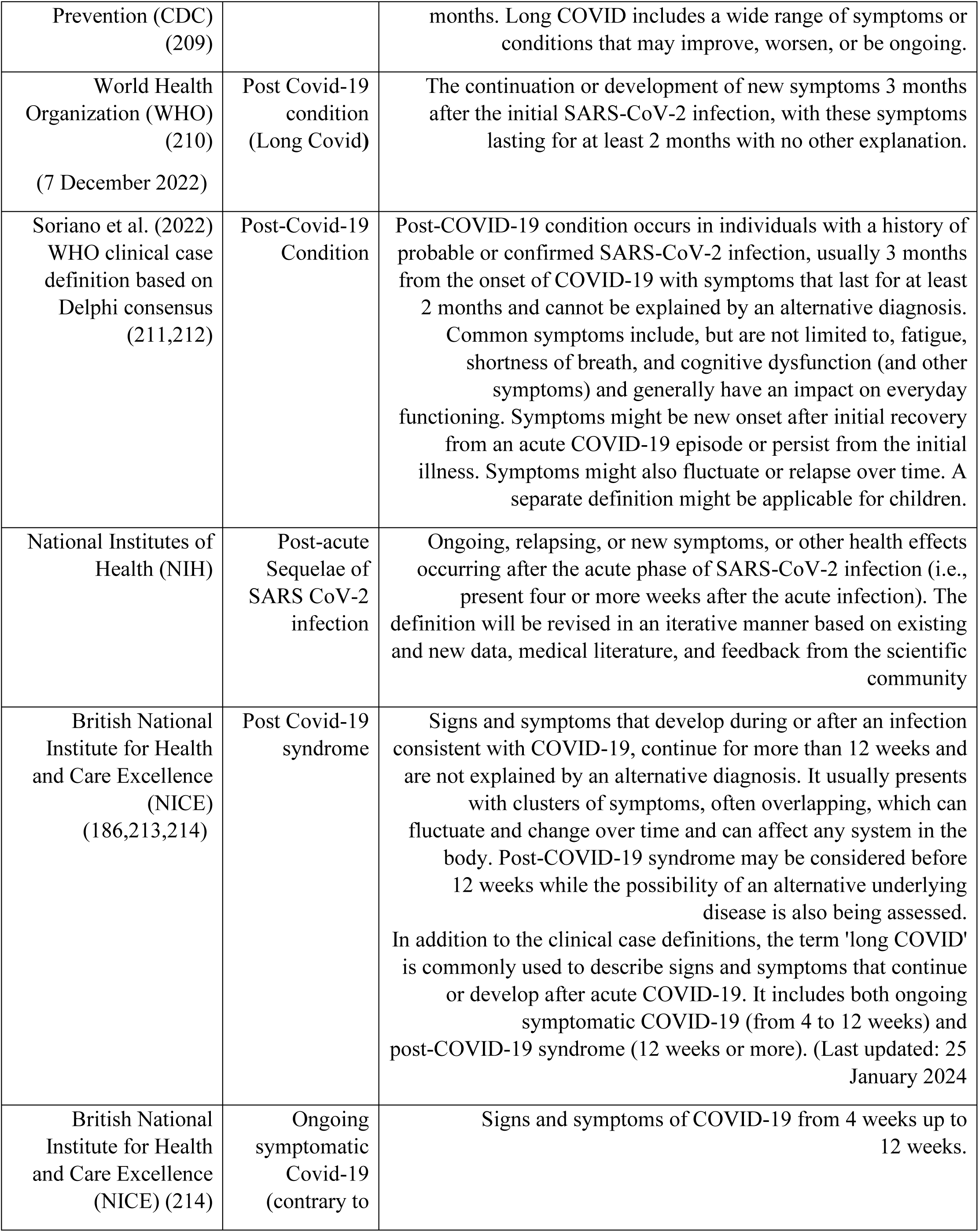
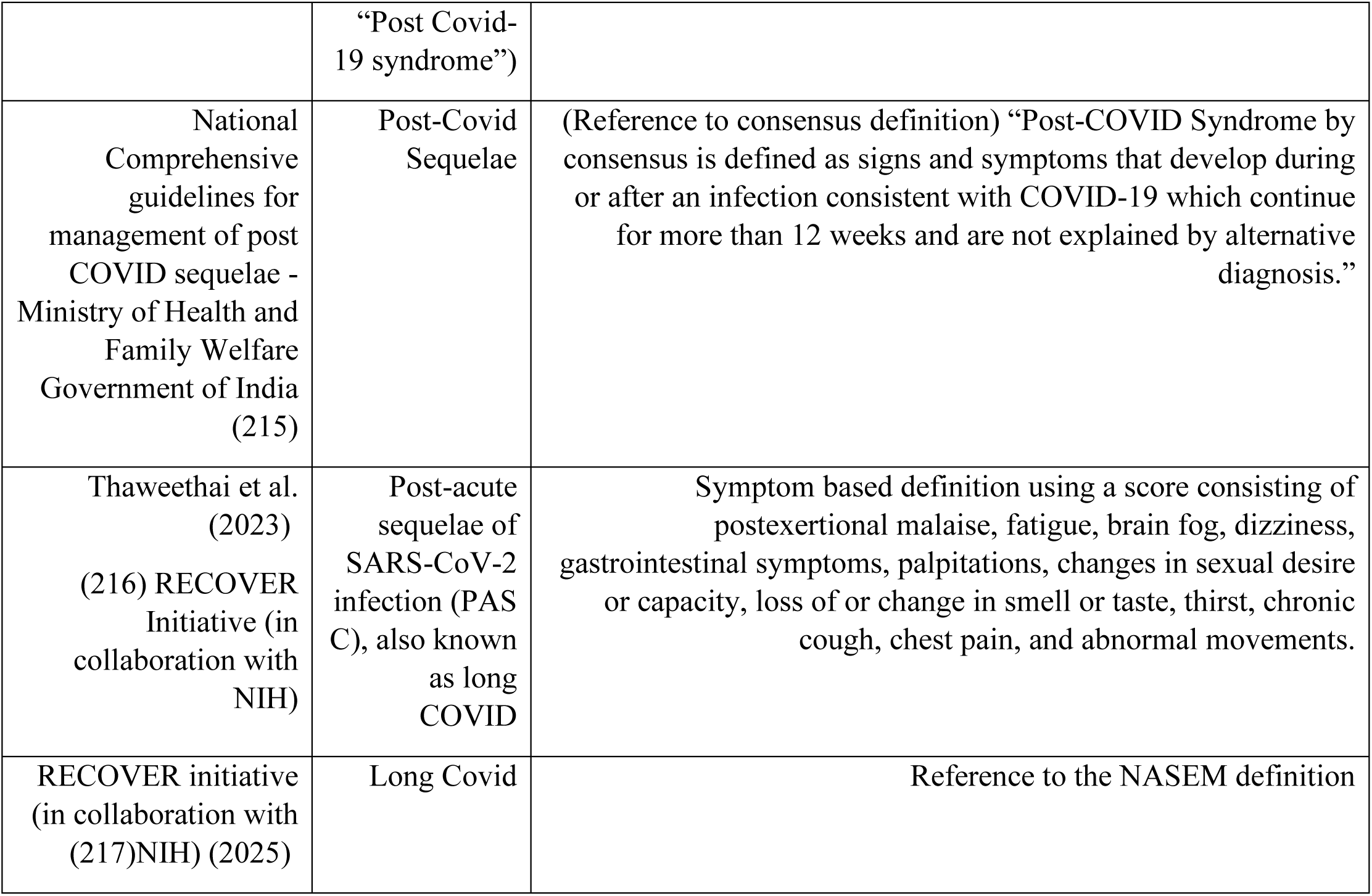
Selective example of definitions and terminologies used for persisting COVID symptoms.

| Source | Term | Definition |
| --- | --- | --- |
| United States National Academies of Science Engineering and Medicine (NASEM) 2024 (207,208) | Long Covid | An infection-associated chronic condition that occurs after SARS-CoV-2 infection and is present for at least 3 months as a continuous, relapsing and remitting, or progressive disease state that affects one or more organ systems. |
| Centers for Disease Control and | Long Covid | (Based on the NASEM 2024 definition) a chronic condition that occurs after SARS-CoV-2 infection and is present for at least 3 |
| Prevention (CDC) (209) |  | months. Long COVID includes a wide range of symptoms or conditions that may improve, worsen, or be ongoing. |
| World Health Organization (WHO) (210)<br>(7 December 2022) | Post Covid-19 condition (Long Covid) | The continuation or development of new symptoms 3 months after the initial SARS-CoV-2 infection, with these symptoms lasting for at least 2 months with no other explanation. |
| Soriano et al. (2022)<br>WHO clinical case definition based on Delphi consensus (211,212) | Post-Covid-19 Condition | Post-COVID-19 condition occurs in individuals with a history of probable or confirmed SARS-CoV-2 infection, usually 3 months from the onset of COVID-19 with symptoms that last for at least 2 months and cannot be explained by an alternative diagnosis. Common symptoms include, but are not limited to, fatigue, shortness of breath, and cognitive dysfunction (and other symptoms) and generally have an impact on everyday functioning. Symptoms might be new onset after initial recovery from an acute COVID-19 episode or persist from the initial illness. Symptoms might also fluctuate or relapse over time. A separate definition might be applicable for children. |
| National Institutes of Health (NIH) | Post-acute Sequelae of SARS CoV-2 infection | Ongoing, relapsing, or new symptoms, or other health effects occurring after the acute phase of SARS-CoV-2 infection (i.e., present four or more weeks after the acute infection). The definition will be revised in an iterative manner based on existing and new data, medical literature, and feedback from the scientific community |
| British National Institute for Health and Care Excellence (NICE) (186,213,214) | Post Covid-19 syndrome | Signs and symptoms that develop during or after an infection consistent with COVID-19, continue for more than 12 weeks and are not explained by an alternative diagnosis. It usually presents with clusters of symptoms, often overlapping, which can fluctuate and change over time and can affect any system in the body. Post-COVID-19 syndrome may be considered before 12 weeks while the possibility of an alternative underlying disease is also being assessed. In addition to the clinical case definitions, the term 'long COVID' is commonly used to describe signs and symptoms that continue or develop after acute COVID-19. It includes both ongoing symptomatic COVID-19 (from 4 to 12 weeks) and post-COVID-19 syndrome (12 weeks or more). (Last updated: 25 January 2024) |
| British National Institute for Health and Care Excellence (NICE) (214) | Ongoing symptomatic Covid-19 (contrary to | Signs and symptoms of COVID-19 from 4 weeks up to 12 weeks. |
|  | “Post Covid-19 syndrome”) |  |
| National Comprehensive guidelines for management of post COVID sequelae - Ministry of Health and Family Welfare Government of India (215) | Post-Covid Sequelae | (Reference to consensus definition) “Post-COVID Syndrome by consensus is defined as signs and symptoms that develop during or after an infection consistent with COVID-19 which continue for more than 12 weeks and are not explained by alternative diagnosis.” |
| Thaweethai et al. (2023) (216) RECOVER Initiative (in collaboration with NIH) | Post-acute sequelae of SARS-CoV-2 infection (PASC), also known as long COVID | Symptom based definition using a score consisting of postexertional malaise, fatigue, brain fog, dizziness, gastrointestinal symptoms, palpitations, changes in sexual desire or capacity, loss of or change in smell or taste, thirst, chronic cough, chest pain, and abnormal movements. |
| RECOVER initiative (in collaboration with (217)NIH) (2025) | Long Covid | Reference to the NASEM definition |

Overall, there is inconsistency in LC definitions and research criteria across studies, the formal international bodies vary in their exact definitions which aren’t necessarily operative and applicable for a research study, and empirical studies don’t always employ or refer to an official LC definition.

#### 3.1.2. Assessment and Measurement Tools

Methodological consistency in the field could be beneficial, and is important for generalizability and for better coherence in future research and meta-analyses. Recurring instruments used for assessments and measurements in empirical studies as found in the reviewed literature are as follows.

Questionnaires and scores such as:

- The visual analogue scale: for multiple measures such as pain and fatigue.
- The brief pain inventory (BPI) (128) and verbal numeric pain rating scale: for assessing pain (148).
- Fatigue Severity Scale (128): for assessing fatigue.
- Insomnia Severity Index: for the evaluation of insomnia (16).
- Fibromyalgia Symptom Scale (FSS) including the widespread pain index (WPI) and symptom severity scale (SSS) based on the ACR fibromyalgia diagnostic criteria and/or modified for self-administration (10,128), Fibromyalgia Rapid Screening (FIRST) questionnaire (135), and the central sensitization inventory (CSI) (146): for assessment of fibromyalgia-type features, screening, or diagnosis. It is worth noting here that the CSI has not been validated to assess or measure central sensitization or neural activity (29), despite several studies using it for this purpose.

Fibromyalgia Impact Questionnaire (or revised version) and its counterpart Symptom Impact Questionnaire (SIQ or SIQ-revised) for assessing psychosomatic disease burden (136,137). More data regarding fibromyalgia questionnaires can be found in a systematic review by Carrasco-Vega et al. (2023) (218).

- Post-COVID-19 Functional Status (PCFS) self-reporting version: for assessing functional status post-COVID-19 infection.
- Yorkshire Rehabilitation Scale (C19-YRS) questionnaire: for assessing LC impact and need for rehabilitation in LC patients.
- The European Quality of Life Instrument versions (16,146) and Short Form 36 (16): for evaluating health-related quality of life.
- Versions of the Patient Health Questionnaire (PHQ-2, 8, 9): for depression assessment.
- Patient Health Questionnaire 15 (PHQ-15) for assessing somatic symptoms.
- Hospital Anxiety and Depression Scale (16) and Generalised Anxiety Disorder-7 scale: for anxiety assessment.

Other measurement tools and methods more commonly used were:

- Quantitative sensory testing (88).
- Algometer for pressure pain threshold (75,148).
- Cold pressure test for conditioned pain modulation (75).
- Diagnostic codes of the international classification of diseases when using data from medical records.

Table 2 provides a comprehensive summary of the instruments and methods used in studies.

**Table 2:** Summary of tools and methods that were used in the reviewed studies. Some of these were used more often than others, and some only in one study. The studies cited for each tool are a selected example for reference. The tools are not listed by order of their frequency used in studies.

| Patient-filled questionnaires and scores |  | Other measurement tools and methods |
| --- | --- | --- |
| The visual analogue scale for multiple measures such as pain and fatigue | The brief pain inventory (BPI) (104,128), short form BPI (BPI-sf) (112) and numeric pain rating scale for assessing pain (148) | Physical exam (68,130) |
| Chronic Pain Grading Scale (CPGS) for intensity of chronic pain (172) | Patient-Reported Outcomes Information System (PROMIS) Pain Interference v1.1 (PROMIS-PI) computerized adaptive test were used to evaluate pain and pain interference (128) | Diagnostic codes of the international classification of diseases (134) |
| Fatigue Severity Scale (FSS) (128) and Multidimensional Fatigue Inventory (MFI-20) (16,67) for assessing fatigue | Multi-dimensional assessment of fatigue questionnaire and global fatigue index (156), Modified Fatigue Impact Scale (MFIS) (172), Chalder Fatigue Scale (69,124), Pichot scale (68) for fatigue | The world health organization's definition of post-COVID-19 (104,128) |
| PROMIS v1.0 Fatigue (PROMIS-Fatigue) computerized adaptive test questionnaire was used to evaluate self-reported daily fatigue symptoms (128) | The FACIT Fatigue Scale (81) for assessing severity of fatigue | LC definition according to the British NICE guidelines (112) |
| dePaul Symptom Questionnaire for post-exertional malaise (67) or for assessing the symptomatology and case definition fulfillment of ME/CFS (81) | General Symptom Questionnaire (GSQ) (64) to assess symptoms of people with irritable bowel syndrome, ME/CFS and fibromyalgia. | Rome III criteria (108) |
| Insomnia Severity Index (ISI) was used for the evaluation of insomnia (16) | The PROMIS Sleep Disturbance (PROMIS-SD) computerized adaptive test survey was used to measure self-reported quality of sleep for a 7-day period (128), Pittsburgh Sleep Quality Index (PSQI) for sleep quality (81). single-item sleep quality scale (91) | Bierle et al. (2021) criteria for indicating LC, developed by a modified Delphi process consisting of scores based on necessary conditions and major and minor criteria (144) |
| Fibromyalgia Symptom Scale including the widespread pain index and symptom severity scale based on the ACR criteria (81,130) and modified for self-administration (10,128), the central sensitization inventory (CSI) (146), Fibromyalgia Rapid Screening Tool (135), and at home fibromyalgia questionnaire (85), for assessment of fibromyalgia-type features, screening, or diagnosis | Fibromyalgia impact questionnaire (or revised version) for classifying fibromyalgia severity, and the Symptom Impact Questionnaire Revised for a fibromyalgia-neutral version (135) | The 2015 Institute of Medicine ME/CFS Diagnostic Criteria (128), and Fukuda et al. criteria for CFS (141) |
| Post-COVID-19 Functional Status self-reporting version, for assessing functional status post-COVID-19 infection (89,148) | Lau et al. (2024) (159) post-acute COVID-19 syndrome 14-item improvement questionnaire (PACSQ-14) | Algometer for pressure pain threshold in each limb, or trapezius, and/or center of rectus femoris, or other (75,148) |
| COVID-19 Yorkshire Rehabilitation Scale (112,114) for capturing the impact of LC and for the assessment of overall health status before and after infection. It is also used for assessment of rehabilitation needs in LC | Long COVID questionnaire (133) | Nerve conduction studies, quantitative sensory testing (e.g., cold detection threshold, warm detection threshold, thermal sensory limen to assess paradoxical heat sensation, heat pain threshold, mechanical detection threshold, mechanical pain threshold, mechanical pain sensitivity, dynamic mechanical allodynia, wind-up ratio, vibration |
|  |  | detection threshold, and pressure pain threshold.) (112), cold pressor test for conditioned pain modulation (75), and other neurological tests (101) |
| EuroQol with five dimensions and three or five levels (16,104,112,146), and 36-item Short Form health survey (SF-36) and its components (16,68,89) and SF-12 (133) for evaluating quality of life and/or health-related quality of life. | WHOQOL-BREF for assessing quality of life (130) | Exercise-Induced Hypoalgesia (EIH) protocol (75) |
| Patient Health Questionnaire 9 (PHQ-9), PHQ-8 and PHQ-2 were used for depression screening/assessment (16,112,128), Beck Depression Inventory (135), BDI-II (183), Center for Epidemiologic Studies of Depression (CES-D) scale for assessing depression (137) | Hospital Anxiety and Depression Scale for anxiety and Generalised Anxiety Disorder-7 scale (GAD-7) (16,112). The State-Trait Anxiety Inventory (183) | Peripheral venous blood sampling (for hormone concentrations, circulating mitochondrial DNA, and more) (67) |
| Depression, Anxiety and Stress Scale (DASS-21) for assessing stress, anxiety, and depression (104) and perceived stress scale (161) for assessing stress. | The Patient Health Questionnaire 15 (PHQ-15) for assessing somatic symptoms (93,219) | RT-PCR, ELISA, or serology for testing for SARS-CoV-2 (active or past infection) |
| Somatic Symptom Disorder-B criteria Scale (SSD-12) for psychological burden associated with the persistent symptoms (68) | The Symptom Impact Questionnaire-Revised (SIQR) (137) to assess symptom severity and functional ability in individuals with chronic pain symptoms (128) | Timed Up and Go test (112) for balance and mobility, Assessment of gait using wearable device (156) |
| Pain catastrophizing scale (PCS) (75,137) for assessing catastrophic thinking related to pain in adults | Pain attitudes questionnaire-revised (PAQ-R) (75) | Handgrip strength test using dynamometry (89,112) |
| Emotion Regulation Questionnaire (ERQ) (172) | Acceptance and Action Questionnaire-II (AAQ-II) (172) | Head-up tilt test for orthostatic intolerance testing (based on consensus criteria) (89) |
| Fear Avoidance Beliefs Questionnaire for assessing fear avoidance and beliefs (104) | The Coping Strategies Questionnaire-Catastrophizing subscale (CSQ-CAT) queried catastrophic thinking related to pain by quantifying a person's pain experience and analyzing thoughts and feelings pertaining to their pain experience (128) | Maximal incremental ramp exercise test (67) |
| Polysymptomatic Distress Scale (130) | Pain Self-Efficacy Questionnaire (112) for assessment of individual's confidence in their ability to perform activities despite pain. | 7-day step count (168), Accelerometer for physical activity monitoring (67,89) |
| Brief resilience scale (161) | Short form of Profile of Mood States (75) | Near-infrared spectroscopy (67) |
| DS-14 questionnaire for assessing Type D personality (133). | The Multi-dimensional Perceived Social Support scale for assessing a person's subjective perception of the extent of his or her social support (133) | Cardio-pulmonary tests such as spirometry, echocardiography, continuous gas analyzer (89) |
| Memory Failures of Everyday (MEF-30) (172) | Five Facet Mindfulness Questionnaire (FFMQ-24) (172) | Aortic pulse wave velocity using arteriography (89) |
| The Multidimensional Inventory of Subjective Cognitive Impairment (MISCI) was used to assess self-reported cognitive abilities related to memory, verbal language ability, general mental clarity, attention/concentration, and executive functioning (128), and NeuroTrax MindStreams Computerized Battery (157) for cognitive assessment. | Montreal Cognitive Assessment (MoCA) (80) and Mini-Mental State Examination (MMSE) (80) | Flow cytometry (81,99) |
| Cognitive Failure Questionnaire (81) | Subjective Cognitive Decline (SCD) questionnaire for assessing cognitive complaints (133) | Gene expression assay, proliferation assay, Elispot Assay (81), microarray and blood transcriptome, combining machine learning (77), RNA-sequencing and more (99) |
| The Tampa Scale of Kinesiophobia – 11 (TSK-11) assessed fear-related beliefs pertaining to physical movement and re-injury in individuals with chronic pain (128) | Godin-Shephard Leisure-Time Physical Activity (89) | Genomic studies including genome wide association study and a combinatorial analysis platform (61) |
| International Physical Activity Questionnaire-short form (112) | Physical activity readiness questionnaire (PAR-Q) (75), International Physical Activity Questionnaire (IPAQ) (75,104), The PROMIS Physical Function (PROMIS-PF) computerized adaptive test was used to measure self-reported physical function (128) | Metabolomics (67), Spectroscopy and chemometrics (135) |
| Physical activity scale of Ricci & Gagnon (68) previously used in studies of elderly population | Functional Impairment Checklist (124). | Immunoassay (81,112) |
| Work Ability Index (161) | Roland Morris Low-Back pain Disability Questionnaire (113) | Interviews (93) |
| BORG Rating of Perceived Exertion (RPE)<br>(112) for perceived effort during Timed Up and Go test | The Multisensory Amplification Scale (MSAS) was used to assess heightened sensitivity to vision, hearing, tactile, smell, and internal bodily perceptions (128) | Clinical evaluation by medical work-up (laboratory, imaging, referral to specialist, etc.) (118) according to guidelines (68) |
| Medical Research Council breathlessness scale (91).<br>Modified Medical Research Council Dyspnea Scale (89).<br>FACIT-Dyspnea (81), London chest activity of daily living (LCADL) scale was assessed to evaluate the level of dyspnea during activities of daily living (148) | The PROMIS Dyspnea Severity (PROMIS-DS) Computerized Adaptive Test questionnaire was used to assess the severity of shortness of breath in response to activity (128) | Skin biopsy for investigating small fibre pathology (220) |
| RA Health Assessment Questionnaire Disability Index (130) | The RA Disease Activity Score-28, Clinical Disease Activity Index, and Simplified Disease Activity Index for rheumatoid arthritis disease evaluation (130). | Muscle biopsy from vastus lateralis for investigating skeletal muscle (e.g., respirometry, immune-histochemistry, immunofluorescence, etc.) (67) |
| Patient Phenotyping Questionnaire Short Form (168), | PainDETECT questionnaire (113) | Neuroimaging, fMRI, Diffusion Tensor Imaging (157) |

### 3.2. Long COVID-19 mechanisms

Elucidating the mechanism of LC is still a matter of ongoing research. To give a brief overview, putative LC patho-mechanisms, as found in the literature, included: immune dysregulation and/or autoimmunity (73,78,86,95,99,140), stress-induced small fiber neuropathy (21), mitochondrial dysfunction (81), metabolic abnormalities (81), infection induced genetic or epigenetic changes (83), impaired hemopoiesis (81), blood-brain barrier damage, endothelial dysfunction (80), direct viral invasion and cytotoxicity, cytokine storm (142), persistence of viral particles in peripheral tissue (55,86), re-activation of latent pathogens (55,140), dysbiosis (159) and/or disruption of the gut-brain axis (86,159), dysautonomia, hormonal imbalance (55), amyloid-containing deposit accumulation in blood vessels causing local hypoxia (67), reduced cellular aerobic capacity, skeletal muscle injury and inflammatory myopathy (88), deconditioning or skeletal muscle atrophy (88), coagulation abnormalities or microthrombi, cerebral vasculopathy (90), psychosomatics (30,68,98), central sensitization, neuroinflammation, glial-cell reactivity, brainstem dysfunction or other central neurological pathology (24,51,72,86,90,139,142), each of these which may plausibly overlap with another in a multifaceted pathophysiology. Correspondences in the field of rheumatology regarding the parallels between LC and fibromyalgia, and discussions on LC being regarded as psychosomatic or non-physiological, were also found (8,30,53), as well as criticism of the dualistic psychological-physiological medical thinking of physical versus mental illness (97,98). An integrative framework of interaction of biological, social, experiential, and psychological factors in LC functional somatic symptoms was advocated (97).

There are some authors who suggest that all chronic pain must be considered in the context of the biopsychosocial model, although the evidence for this is still mixed (102). Other authors seem more inclined to suggest that emotional stress associated with COVID-19 could trigger the onset of post-COVID fibromyalgia (53,92). Several authors discuss the idea of coronavirus inducing central sensitization through neuroinflammation (51,77,79,112,114,146). Goldenberg (2024) (55) gives an overview of the overlap between LC and fibromyalgia-type syndromes, covering the latest empirics in relation to current theories, and argues for a refined definition of LC that’s limited to persistent multisystem symptoms in the absence of well-defined organ damage.

Calabrese and Mease (2024) (43) note that emotional dysregulation is often attributed to fibromyalgia pathogenesis in theory but argue against it being the primary pathogenic cause, pointing out a lack of strong evidence that psychological stress causes fibromyalgia, and there being few, if any, prospective longitudinal studies that show this to be the case. Instead, they suggest a two-way relationship where pain treatment can improve emotional issues, as was shown in empirical studies (43), implying pain itself significantly contributes to emotional dysregulation. Even if some commonality among mechanisms exists, fibromyalgia after COVID-19, they argue, shouldn’t be regarded as a synonym for LC but seen as a part of a more complex post-acute illness. Meanwhile, in a study by Appelman et al. (2024) that investigated skeletal muscle biopsies of 25 LC adult patients after inducing post-exertional malaise by maximal exercise test, several abnormalities were found including extracellular amyloid deposits, metabolic dysfunctions, and more, compared to non-LC individuals (67). These findings are suggestive of a muscle tissue involvement in LC.

Contrary to the mainstream literature on the subject, Shoenfeld and colleagues reiterate the possibility of expression of functionally active autoantibodies against epitopes belonging to the autonomic nervous system as a possible pathobiology to explain LC (14,74,76,78), seen by them as a syndrome of autonomic imbalance that overlaps with fibromyalgia and related syndromes. Empirical studies indeed found several distinct functionally active autoantibodies in LC patients (100), at least some of which seem to be associated with COVID-19 severity (14). Yet, the adequacy of such a theory, however appealing it may be, to mild and asymptomatic infection of coronavirus leading to LC, remains an open question (51,81,91,93). The authors also emphasized, rightfully so, that there is substantial controversy with regards to the etiology and pathophysiology of fibromyalgia syndrome (14). Recently, Yin et al. (2024) published compelling evidence underscoring immune dysregulation in LC (99) pointing towards improper crosstalk between cellular and humoral adaptive immunity.

A comprehensive essay on LC’s putative pathophysiology, which wasn’t the purpose of this scoping review, can be found in publications dedicated to this subject (1,51,90,95,221).

In summary, there are plenty of ongoing speculations on LC pathobiology and mechanism, some of which are substantiated more by empirical studies and some less, but a comprehensive theoretical explanation for LC, let alone for post-COVID-19 fibromyalgia, and a rational organic mechanism that offers an effective treatment and enables a-priori successful theory-based predictions are still lacking as research is ongoing.

### 3.3. Observational studies on widespread musculoskeletal pain and fibromyalgia after sars-cov-2 infection

#### 3.3.1. Cross-sectional and cohort studies on post-Covid fibromyalgia prevalence and incidence

Accumulating evidence indicates that persistent fibromyalgia-type symptoms, including widespread pain, fatigue, and cognitive impairments, can develop following COVID-19 infection as part of a post-viral infection syndrome. Early investigations on fibromyalgia post-COVID-19 are somewhat informative about epidemiology and prevalences. Studies used various methodologies and study designs, measurement tools, outcome measures, inclusion criteria, and, when relevant, control groups. Several of the studies that were found used online surveys and questionnaires inquiring into manifestations of chronic pain and fibromyalgia-type features (e.g., low mood, anxiety, myofascial-type pain, stress, fatigue, sleep impairment, functional impairment and decreased quality of life) after COVID-19, and multiple studies implemented a fibromyalgia self-reported questionnaire or used the ACR criteria as part of the outcome measures (10,16,81,85,104,111,114,115,118,125,131,132,146,148). Large population-based observational studies utilizing data from healthcare databases were also found. A few studies are described in more detail in this section to demonstrate the variety of methods and the finding that stem from them. The remaining observational studies on the subject of fibromyalgia post-COVID-19 that were included in this review are summarized in Table 3 below: A nationwide exploratory cross-sectional study out of Denmark and Spain by Ebbesen and colleagues (2024) (105) investigated the prevalence and risk factors of de novo widespread musculoskeletal pain after COVID-19 in non-hospitalized COVID-19 survivors. Demographic and medical data were collected through an online questionnaire from Danish adults with a confirmed SARS-CoV-2 infection that occurred at least 6 months prior to the study, between March 2020 and December 2021, a period mostly consisting of the first SARS-CoV-2 strains. Among 130,443 non-hospitalized respondents (58.2% women, mean age was 50.2 years), 5.3 percent of non-hospitalized COVID-19 survivors had new-onset widespread musculoskeletal pain at approximately 14 ± 6.0 months after infection, which was rated as moderate to severe in its intensity in 75.6% of cases. In a multivariate analysis, female sex, age, higher body-mass index (BMI), and previous history of migraine, whiplash, stress, type-2 diabetes, and comorbid chronic neurological disorders, were found as risk factors for de novo widespread post-COVID pain, with adjusted odds ratio of 1.549, 1.003, 1.043, 1.554, 1.562, 1.47, 1.56, and 1.532, respectively.

**Table 3:** Summary of observational studies on new-onset fibromyalgia-type or myofascial-pain manifestations after COVID-19 in the systematic scoping review.

| Topic/context | Study | Description of study | Main findings |
| --- | --- | --- | --- |
| Pain after COVID-19 | Amsterdam et al. (133) | A cross-sectional survey via self-reported questionnaires explored the association between distinctive personality profiles, particularly type D personality, and LC among convalescent asymptomatic to mild acute-COVID-19 cases without a need for hospitalization or oxygen supplementation. Adult participants were recruited from a pool of 750 individuals undergoing follow-up at the Tel Aviv Sourasky Medical Center post-COVID-19 clinic as outpatients. | 31% completion rate yielded 114 respondents (74.6% women), mean age was 44.5 years. 68.4% were healthy prior to contracting COVID-19 and developing LC. 37 of 114 met diagnostic criteria for fibromyalgia, and in 28 (24.5%) it was a new diagnosis after COVID-19. None of the patients reported experiencing prior mental health issues, nor did they have previous psychiatric diagnoses. Clustering into two groups showed an association between more pronounced fibromyalgia features, a higher burden of depression and anxiety, diffuse pain, attention deficit, memory problems, headaches, perception of lower quality of life, and type D personality, as well as a trend towards poor sleep quality. |
| Pain after COVID-19 | Damasceno et al. (115) | A case control study from Brazil that aimed to establish etiological factors associated with chronic pain syndromes in adult patients with post-COVID-19 conditions during 2021. Participants were adults who had COVID-19 at least 3 months prior to data collection with and without chronic pain syndromes. CSI was used to assess “central sensitivity” (i.e., fibromyalgia-type manifestations) | In total, 120 individuals were recruited (51 patients and 69 controls, average age ~30 years). CSI scores differed significantly between the groups with average scores of 50.51 in patients vs. 24 in controls ($p < 0.001$). |
| Pain after COVID-19 | Ebbesen et al. (105) | A nationwide cross-sectional study to investigate the prevalence and risk factors of de novo widespread musculoskeletal pain after COVID-19 in non-hospitalized COVID-19 survivors. Demographic and medical data were collected through an online questionnaire from Danish adults with a confirmed SARS-CoV-2 infection at least 6 months prior to the study, between March 2020 and December 2021. Widespread pain was defined as participants experiencing pain in at least 2 sites of the body, in the upper part of the body and 1 site on the lower part. | Among 130,443 nonhospitalized respondents (58.2% women, mean age was 50.2 years), 5.3 percent (n=6,875) of nonhospitalized COVID-19 survivors had new-onset widespread musculoskeletal pain at approximately $14 \pm 6.0$ months after infection, which was rated as moderate to severe in its intensity in 75.6% of cases. In a multivariate analysis, female sex, age, higher BMI, and previous history of migraine, whiplash, stress, type-2 diabetes, and comorbid chronic neurological disorders, were found as risk factors for de novo widespread LC pain, with adjusted odds ratio of 1.549, 1.003, 1.043, 1.554, 1.562, 1.47, 1.56, and 1.532, respectively. Also, among a few other factors found to be significant, higher income was associated with less development of widespread pain. Time elapsed since infection was also significantly positively correlated. Rates differed according to stratification by SARS-CoV-2 variant. |
| Pain after COVID-19 | Ketenci et al. (111) | A multicenter cross-sectional survey that was conducted during 2021 in physical and rehabilitative medicine outpatient clinics in Turkey categorized chronic pain after COVID-19 into predetermined categories. Diagnosis of pain phenotypes | In total, 437 patients were grouped by diagnosis into predetermined chronic pain phenotypes, and subjects with overlapping clinical features were excluded. 66.13% of the patients were diagnosed with nociceptive pain, 11.67% with neuropathic pain, and |
|  |  | (nociceptive, neuropathic, or nociplastic/central sensitization) was made by physicians according to data from outcome measures including Pain Numerical Rating Scale, CSI, BDI, and HADS, Self-Report Leeds Assessment of Neuropathic Symptoms and Signs, clinical examination, and their experience in musculoskeletal diseases. Patients with overlapping phenotypes were excluded. | 22.20% with central sensitization based on the CSI (i.e., fibromyalgia-type features). According to the authors, central sensitization was associated with females, hypertension, physical activity, and pre-existing chronic disease prior to COVID-19. |
| Pain after COVID-19 | Khoja et al. (112) | As part of a larger UK longitudinal study on musculoskeletal pain in LC (MUSLOC), cross-sectional data was reported on 30 adults with a history of COVID-19 with a diagnosis of LC and new onset musculoskeletal pain. The COVID-19 Yorkshire Rehabilitation Scale (C19-YRS) was used to capture the overall impact and health state before and after COVID-19 infection and the symptoms and their effect on individuals functioning. Other outcomes measures and self-assessment tools included QST and time up and go test, PHQ-9, GAD-7, PCS, EuroQoL, and additional other tools. Central sensitization in participants was recognized if there was one of the three specific criteria: abnormally increased mechanical pain sensitivity, a reduced mechanical pain threshold, or presence of dynamic mechanical allodynia. | 30 participants in total (19 female) were included. The mean duration from the onset of musculoskeletal pain to evaluation in the study was 519.1 days (± 231.7). Only three participants were hospitalized due to COVID-19. Forty percent had no pre-existing medical condition. New-onset chronic musculoskeletal pain was mostly reported by the participants as generalized widespread pain (90%), characterized predominantly as joint pain. Ninety percent of the participants experienced continuous pain that always remains present, though its intensity may vary. 82.8% reported a high interference score, 19 participants stated that their employment status was affected by the health consequences associated with LC. QST indicated mechanical hyperalgesia and gain of function for wind up ratio, suggesting enhanced temporal summation of pain. In total, 25 participants (83%) showed central sensitization signs. There was a variability in the cytokine profiles. Investigation into individual cytokine levels using univariate analysis revealed no association between pain scores and any individual cytokines or C-reactive protein (CRP). The authors conclude that chronic new-onset musculoskeletal pain in LC tends to be generalized, widespread, continuous and is associated with central sensitization, elevated pro-inflammatory cytokines, weakness, reduced function and physical activity, depression, anxiety, and reduced quality of life. |
| Pain after COVID-19 | Khoja et al. (114) | An observational study that was conducted as part of a larger Musculoskeletal Pain in Long COVID (MUSLOC) UK study. Participants were adults (18 years or older) that tested positive for COVID-19 or had COVID-19 symptoms confirmed by an independent clinician, received a clinical diagnosis of post-COVID-19 syndrome according to the NICE guidelines, and experienced new-onset musculoskeletal pain since COVID-19. LC-associated symptoms were assessed using the | In total 18 patients were recruited, mean age was 49.6 (± 11.8) years, comprising 12 females (66.7%), mean duration since the onset of COVID-19 infection to the data collection point was 27.9 (± 6.97) months. Fourteen (77.8%) patients reported experiencing generalized widespread pain, while the remaining patients, who did not report widespread pain, still experienced pain in at least four distinct body areas. LC symptoms interfered with daily living activities for 17 (94.4%) patients, 13 (72.2%) of the evaluated patients met the diagnostic |
|  |  | COVID-19 Yorkshire Rehabilitation Scale (C19-YRS) questionnaire. The assessment of fibromyalgia was conducted as part of the standard clinical examination and using the American College of Rheumatology (ACR) 2010 criteria. | criteria for fibromyalgia as defined by the ACR. The average WPI score among the patients was 8.8, indicating a high level of pain spread across multiple body regions. Additionally, the average SS score was 8.2, reflecting significant symptom severity related to fatigue, waking unrefreshed, cognitive symptoms, and the extent of other somatic symptoms. Patients that did not meet the high cut-off of the ACR criteria for diagnosis still had fibromyalgia features of widespread pain. |
| Pain after COVID-19 | Kim et al. (127) | A 2022 cohort study that used data from electronic medical records from a nationwide population of all persons with COVID-19 in South Korea. Included were only individuals who had been diagnosed with COVID-19 during the first four months of the pandemic (February to May 2020) by means of real-time reverse-transcription polymerase chain reaction (PCR). Individuals in the control group were chosen as those who did not receive PCR testing. The authors investigated incidence rates of pain diagnoses of unspecified or idiopathic pain (e.g., fibromyalgia, headache, etc.), using diagnostic codes of the international classification, as well as prescription of medication as the outcome measures. | The diagnoses of fibromyalgia, temporomandibular joint disorders, and atypical facial pain did not occur at any time during 90 days from the index date. Opioid prescribed medications were higher in the COVID-19 group. When performing subgroup analysis the results were reversed, indicating higher rates of idiopathic pain in the control group. |
| Pain after COVID-19 | Kim et al. (163) | A population-based cohort study to determine changes in the level of incidence of musculoskeletal disorders among the Korean population in pre-pandemic and during the pandemic (through the periods of 2018-2021), using electronic medical record registries of the Korean National Health Insurance Service. The incidence of orthopedic diseases was evaluated based on diagnostic codes of the international classification of diseases. | The incidence of myofascial pain had decreased during the pandemic compared to pre-pandemic levels, while gout and frozen shoulder increased.. |
| Pain after COVID-19 | Patel and Javed (19) | Case series from a pain clinic | Medical records of individuals who developed myofascial pain after a diagnosis of COVID-19 between March 2020 and December 2020 were obtained. Three patients with considerable pre-existing chronic pain conditions experienced worsening musculoskeletal symptoms after SARS-CoV-2 infection. The first patient, a 68-year-old female, developed post-COVID-19 myalgia and muscle spasms, which improved by 75% following trigger point injections and physical therapy. The second patient, a 35-year-old female with congenital scoliosis, developed bilateral shoulder pain post-COVID-19, with taut bands in the<br>infrapinatus muscle, showing moderate improvement with conservative treatment but refusing intervention. The third patient, a 71-year-old male with a substantial orthopedic lumbar medical history, developed new-onset neck pain and headaches post-COVID-19, with palpable taut bands in the trapezius bilaterally (a total of six) referring pain to the occipital region, achieving 40–50% immediate pain relief after myofascial trigger point injections, resulting in a numerical pain rating scale rating of 3/10 (compared to 6/10 pre-intervention) during the 4 week follow-up. |
| Pain after COVID-19 | Zha et al. (162) | Case report | A 59-year-old previously healthy Hispanic male developed persistent myalgia following COVID-19, with pain localized to trigger points in the neck, shoulders, upper back, arms, and legs, consistent with myofascial pain syndrome. Wet needling with lidocaine provided immediate but temporary relief, requiring multiple sessions over months. After experiencing a relapse associated with psychological stress, dry needling was introduced, leading to rapid and sustained pain reduction. |
| Pain after COVID-19 | Gouraud et al. (68) | A retrospective observational study from France investigated the characteristics, medical conclusions, and satisfaction of 286 patients with persistent symptoms after COVID-19 who attended a multidisciplinary day-hospital program. Evaluation was done by medical workup as recommended by official guidelines. | A total of 286 patients (of which 12.7% were hospitalized) were included in the study. The most common symptoms were fatigue, breathlessness, and joint/muscle pain. Cognitive and behavioral features that may contribute to the maintenance of physical symptoms were identified in 75.5% of patients after clinical evaluation and were considered as positive arguments in favor of a diagnosis of functional somatic disorder. Among these patients, 95.6% did not present any abnormal clinical findings or test results that could potentially explain the symptoms, and a diagnosis of functional somatic disorder was established for 72.2% of the patients after the multidisciplinary assessment. Patients with a diagnosis of functional somatic disorder had similar rates of major depression (32.8%) and anxiety disorders (25.0%) than in the whole sample, with no significant difference compared to those without ($\chi^2 = 0.24$, $p = 0.63$ and $\chi^2 = 2.22$, $p = 0.14$). |
| Pain after COVID-19 | Bakilan et al. (121) | A retrospective cross-sectional study aiming to evaluate frequency of musculoskeletal problems in post-acute COVID-19 patients. The study used medical records of LC adult patients who were admitted to the physical medicine and rehabilitation outpatient clinic in Tukey between December 2020 and May<br>and who reported musculoskeletal symptoms. | 280 LC patients were included in the study (65% women, mean age $47.45 \pm 13.92$, 70% not hospitalized). At admission to the outpatient clinic the frequency of symptoms of widespread myalgia was 3.9%, back pain 28.6%, and fatigue was 12.1%. Muscle pain in more than one site that was initiated or aggravated with COVID-19 was 51.1%. |
| Somatic symptoms in LC | Kachaner et al. (182) | A single-centre observational study from France that assessed the diagnosis of somatic symptom disorder (SSD) in patients with unexplained long-lasting neurological symptoms after mild COVID-19. Consecutive patients referred to a neurologist for post-COVID-19 consultation were reviewed. Main outcome was positive diagnosis of SSD according to the criteria of the Diagnostic and Statistical Manual of Mental Disorders- 5 (DSM- 5). Brain MRI findings were extracted from patient records | 32 of 50 patients (64%) met the DSM-5 criteria for SSD. In the remaining 36%, SSD was considered possible given the high scores on diagnostic scales. Physical examinations were normal for all patients. Brain MRI showed unspecific minor white matter hyperintensities in 17% (8/46) of patients, considered non-specific findings, consistent with prevalence in the general population of that age range. Neuropsychological assessment (in 15 patients) showed exclusively mild impairment of attention in 93% (14/15), in discrepancy with their major subjective complaint. A high proportion of patients (90%, n=45/50) met criteria for chronic fatigue syndrome. A significant number of patients screened positive for mood-anxiety disorders (32%, n=17/50), had a history of prior SSD (38%, n=19/50), and reported past trauma (54%, n=27/50). Self-survey results highlighted post-traumatic stress disorder in 28% (12/43), high levels of alexithymia traits (42%, n=18/43), and high levels of self-oriented perfectionism (79%, n=33/42) |
| Chronic fatigue syndrome | Das et al. (61) | A study that investigated genetic risk factors associated with ME/CFS using combinatorial analysis on genotype data from 2,382 ME/CFS patients reporting a diagnosis in the UK Biobank Pain Questionnaire, matching them against 4,764 controls. | The study stratifies ME/CFS patients genetically and correlates this stratification with clinical criteria. Biological analysis of identified genes reveals links to key cellular mechanisms hypothesized to underpin ME/CFS, such as vulnerabilities to stress and infection, mitochondrial dysfunction, sleep disturbance, and autoimmune development. |
| Fibromyalgia after COVID-19 | Akel et al. (132) | A 2022 web-based cross-sectional study “to investigate the prevalence and predictors” (associated factors) of fibromyalgia in individuals recuperating from COVID-19, based on the study of Ursini et al. (2021). The ACR survey criteria were used with a cutoff score $\geq 13$. | Out of 404 respondents (75% women, mean BMI 26.6, mean duration of COVID-19 infection was $12.8 \pm 5.3$ days) 89% were treated at home, while only six (1.5%) patients needed a ward admission and one (0.2%) an intensive care unit admission. 80 individuals (19.8%) satisfied the ACR survey criteria for fibromyalgia (out of them 93.8% were women). Females (OR: 6.557, 95% CI: 2.376 - 18.093, $p = 0.001$) and dyspnea (OR: 1.980, 95% CI: 1.146 - 3.420, $p = 0.014$) were associated with post-COVID-19 fibromyalgia. The fibromyalgia group had more pre-existing comorbidities. In bivariate correlation analysis age ($r = 0.200$, $p = 0.001$) and duration of COVID-19 infection ($r = 0.121$, $p = 0.015$) were said to be weakly correlated with fibromyalgia symptom score. |
| Fibromyalgia after COVID-19 | Bileviciute-Ljungar et al. (16) | A Swedish web-based survey combined with face-to-face interviews, using several questionnaires inquiring into mood, pain, fibromyalgia criteria, | A total of 100 individuals (82% female), at a mean of 47 weeks after SARS-CoV-2 infection, 90% were not hospitalized for COVID-19. Irritable stomach, pain in varied |
|  |  | functional status, and quality of life. The study included adults who had COVID-19 according to anamnesis or a positive test, and self-reported a significantly reduced level of functioning and persistent symptoms for more than 12 weeks. | sites, and widespread pain were reported by 75%, 15%, and 50%, respectively (30 out of the 50 with widespread pain were healthy prior to their infection) with a mean pain intensity of 5.16 in the latter. 40 out of 100 fulfilled fibromyalgia criteria, of them 22 indicated being healthy before their infection. Previous comorbidities were found to be associated with generalized pain and with fibromyalgia. Health-related quality of life was decreased in more than 80 percent of individuals, not surprising considering the study's inclusion criteria. |
| Fibromyalgia after COVID-19 | Ganesh et al. (52) | Descriptive paper of 108 patients seen at a Post-COVID clinic at Mayo Clinic during 2021. Clinical symptoms were analyzed and assigned to one of six phenotypes: dyspnea, chest pain, myalgia, orthostasis, fatigue, and headache predominant. Patients with no evidence of tissue damage on testing were determined as likely to have a central sensitization phenotype, which was treated with a virtual treatment program aimed at patient education with elements of cognitive-behavioral therapy, health coaching, and paced rehabilitation. The fatigue-predominant, myalgia-predominant, and orthostasis-predominant phenotypes were considered together as central sensitization phenotypes. | 108 fibromyalgia-type patients were seen (75% female, median age of 46 years, 16% were admitted for acute COVID-19). Patients were evaluated on average 148.5 days after the onset of symptoms (interquartile range, 111.5 to 179.3 days). The most common comorbidities were obesity (39%), anxiety (33%), depression (28%), and gastrointestinal disease (25%), while only 5% had irritable bowel syndrome. At the time of evaluation, the most common symptoms were fatigue, shortness of breath, brain fog, anxiety, and unrefreshing sleep. Patients were classified into six phenotypes: fatigue predominant (n=69), dyspnea predominant (n=23), myalgia predominant (n=6), orthostasis predominant (n=6), chest pain predominant (n=3), and headache predominant (n=1), with more women being predominant for fatigue, orthostasis, and chest pain. The "central sensitization" phenotype (n=82) had statistically significantly higher IL-6 levels ($P=.01$), and higher proportion of women (82%) compared to other phenotypes of post-covid symptoms (54%; $P<.0001$). Age was not significantly different. |
| Fibromyalgia after COVID-19 | Jennifer et al. (129) | A 2022 population based retrospective cohort study using data from electronic healthcare database and a COVID registry, during March 2020 and May 2021, investigating incidence rates of several medical conditions following COVID-19 (including deep vein thrombosis, lung disease, fibromyalgia, diabetes, cerebrovascular accident, myocardial infarction, ischemic heart disease, acute kidney disease, hypertension, use of antidepressants/anxiolytics as an indication of depression/anxiety, and use of benzodiazepines as an indication of sleep disturbance). Records for out-patient and community-based physician or other health profession visits were used. Diagnosis of fibromyalgia was based on hospital and community-based physician visit diagnoses. | Slightly higher crude incidence rates were found for depression/anxiety, sleep disturbance, fibromyalgia (0.28% new cases compared to 0.24% in controls $p = 0.034$ in non-hospitalized cases), deep vein thrombosis, lung disease, and diabetes among convalescent persons after COVID-19. |
| Fibromyalgia after COVID-19 | Martin et al. (125) | Patients recruited from a post-COVID-19 infection clinic were assessed at 6 months using the widespread pain index (WPI), symptom severity scale (SSS), 10 point visual analogue scale for fatigue severity (VAS-F) and a 9-item, 7-point fatigue severity scale. (congress abstract) | At six months following infection, five patients out of 25 recruited in total met criteria for fibromyalgia based on the WPI and SSS. Female patients and patients younger than 60-years-old had higher scores. |
| Fibromyalgia after COVID-19 | Miladi et al. (131) | A web-based cross sectional survey during February 2022 to estimate the prevalence of fibromyalgia in patients who recovered from COVID-19 and to identify associated factors. ACR Survey Criteria and the Fibromyalgia Rapid screening Tool (FIRST) questionnaire were used. (meeting abstract) | A total of 150 respondents (66% women) at an average of $6 \pm 3$ months after the COVID-19 diagnosis, majority in the age group of 21-30, 31% of responders had comorbidities, median BMI was 24.7. Median COVID-19 duration was 7 days with 0,7% of patients requiring hospital admission. ~19% screened positive with the FIRST questionnaire for fibromyalgia. Seven of the 29 subjects with fibromyalgia had seen a physician after the occurrence of widespread pain. Post-COVID-19 fibromyalgia was significantly associated with females ($p = 0.003$), comorbidities ($p = 0.01$) and obesity (0.03). |
| Fibromyalgia after COVID-19 | Scherlinger et al. (118) | A prospective observational study from France that aimed to describe the clinical and biologic characteristics of post-acute COVID-19 syndrome. Consecutive patients seeking medical help for persistent symptoms self-attributed to COVID-19 during the first wave (February to April 2020) of the pandemic underwent a multimodal evaluation. Results were compared to convalescent COVID-19 individuals without persistent symptoms. The study also aimed to investigate the potential underlying mechanisms, including autoimmunity and psychological distress. | 30 patients (60% women) were included in total (7 visited the emergency department, 1 was hospitalized for COVID-19). Patients were clinically evaluated after a median of 152 days following the reported onset of initial symptoms (symptom persistence was median of 6 months duration). Seventeen (56.7%) reported a resolution of initial symptoms after a median of 21 days (IQR 15–33), followed by a resurgence at a median of 21 days later (IQR 15–44). Persistent symptoms had a cyclical pattern in 28 (93.3%) patients and were mostly represented by fatigue, myalgia and thoracic oppression. Fatigue was severe for most patients and rated at a median of 7 (IQR 5–8) on a 10-point scale, with pain rated at 5 (IQR 2–6). The DN4 questionnaire screening neuropathic pain was positive ( $\geq 4/10$ ) for 50% (15/30) of patients, and the FiRST questionnaire screening for fibromyalgia-like symptoms was positive ( $\geq 5/6$ ) for 56.7%. For most clinical features there was no significant difference between immunized and non-immunized individuals. A clinical examination, including neurologic examination, was unremarkable. Nasopharyngeal and stool samples for SARS-CoV-2 RT-PCR tests were negative. Routine biologic test results were within normal limits for all but one patient (iron-deficiency anemia). Screening for autoimmunity revealed low (1/160) and medium (1/320 to 1/640) titers of anti-nuclear antibodies in 12 and 3 patients,<br>respectively. Low to medium anti-nuclear antibody titers were numerically more prevalent in SARS-CoV-2 immunized than non-immunized patients (66.7% vs. 33.3%, $p = 0.067$). 10% (3/30) and 26.7% (8/30) of patients had a previous history of depression and anxiety disorders, respectively. HADS screening for anxiety and depression was positive for 11 (36.7%) and 13 (43.3%) of patients, respectively. Only half of post-acute COVID-19 syndrome patients had cellular and humoral immunity for SARS-CoV-2. |
| Fibromyalgia after COVID-19 | Shani et al. (134) | A retrospective cohort study that investigated the associations between the BNT162b2 vaccine, infection with coronavirus, and the incidence of new registered diagnosis of autoimmune disease within ~1 year follow-up using electronic medical records of a large healthcare database. Vaccinated and unvaccinated individuals were compared as a cohort, and infected and uninfected as another cohort. The minimum follow-up time was four months, during the first phase of the pandemic. Included were individuals 12 years of age and above. Findings were reported in hazard ratios (HR) and incidence rates per 100,00 person years. Statistical analysis included incidence rate ratio tests (univariate), and multivariate Cox proportional hazards models with time-dependent exposure status. Correction for multiple comparisons was applied using the False Discovery Rate (FDR) method to account for the investigation of multiple clinical outcomes. | More than 3 million were included with considerable differences between group characteristics. Vaccination did not influence rates of new registered diagnosis of fibromyalgia in any age group within the timeframe of the study. Infection with COVID-19 increased the risk for fibromyalgia (HR=1.72 95 % CI: 1.36–2.19 in individuals aged 18–44, HR = 1.71, 95 % CI: 1.31–2.22 in individuals aged 45–64) and hypothyroidism (in individuals aged 65 or older). The authors note that the process of reaching diagnoses in the primary care setting in many circumstances is not immediate, therefore, the results should be interpreted with caution. |
| Fibromyalgia after COVID-19 | Sørensen et al. (65) | A nation-wide cross-sectional survey collecting data on self-reported symptoms using web-based questionnaires in Denmark via national e-Boks system (with access to 92% of all residents aged 15 years and above). Cases were chosen based on RT-PCR positive tests that were recorded in the national COVID-19 tracking system. Data were collected from August 1, 2021 to December 11, 2021. Questionnaires evaluated post-COVID symptoms and register-based information supplemented data on risk differences in new onset diagnoses of anxiety, chronic fatigue syndrome, depression, fibromyalgia and post-traumatic stress disorder (PTSD) confirmed by a medical doctor since the test (onset between time of testing and questionnaire completion). | 153,412 individuals fully completed the questionnaire, 61,002 participants tested positive for sars-cov-2. There were significant differences in population characteristics between the study groups (in age, sex, physical activity, and more). Based on the statistical analysis of the findings, at least one diagnosis of depression, anxiety, chronic fatigue symptom (CFS), fibromyalgia, or post-traumatic stress disorder (PTSD) of new onset within the first 6, 9, or 12 months after the test was reported by 7.2% of individuals with a positive COVID test, compared to 3.3% of negatives. Risk for fibromyalgia was not found to be significantly different between the groups, amounting at 1% in COVID positive compared to 1.1% in COVID negatives (risk difference 0.02 95% CI: -0.09-0.14). |
| Fibromyalgia after COVID-19 | Ursini et al. (10) | A cross-sectional online survey via social network among Italian speaking adult individuals ($\geq 18$ years) who developed COVID-19 three or more months prior to the study, with the objective of estimating fibromyalgia prevalence after COVID-19. The Fibromyalgia Symptom Scale, based on the ACR 2016 criteria, was used to identify fibromyalgia by a cutoff score of 13. | 616 eligible individuals completed the survey, (77.4% women). In total, 30.7% fulfilled criteria appropriate for classifying fibromyalgia, 6 months on average after contracting COVID-19. Fibromyalgia was associated with a more severe acute infection, obesity, and with males. |
| Fibromyalgia and LC | Calvache-Mateo et al. (104) | A cross-sectional study from Spain aiming to assess clinical and psychological variables among non-hospitalized adult patients with LC, compared to recovered patients after COVID-19 and healthy controls. Outcomes were assessed using questionnaires such as the brief pain inventory, CSI, insomnia severity index, Tampa Scale for Kinesiophobia, Pain Catastrophizing Scale, Depression, Anxiety and Stress Scale, and Fear Avoidance Beliefs Questionnaire, and analyzed using chi-squared and ANOVA tests to identify group differences. | 170 participants in total (healthy control group $n = 58$, successfully recovered group $n = 57$, and LC group $n = 55$, mean age $\sim 45$ years for all groups). Mean CSI (indicating fibromyalgia-type features with a conventional cutoff score of 40) for the LC group was $54.53 \pm 17.10$ at $105 \pm 12$ weeks since infection on average, which was significantly higher compared to recovered patients ($18.81 \pm 16.24$) and healthy controls ($17.69 \pm 14.30$). Brief pain inventory and insomnia severity index were also significantly higher in the LC group, as well as rates of pharmacological treatment. Scores for the pain catastrophizing scale components relating to helplessness were $9.93 \pm 5.70$ compared to $2.16 \pm 3.09$ in the COVID-19 recovered group. No statistically significant differences were found between the healthy control group and the successfully recovered COVID-19 patients in any variable. |
| Fibromyalgia and LC | Hackshaw et al. (135) | A pilot study aiming to develop a metabolic fingerprint approach for diagnosing clinically similar LC and fibromyalgia using Fourier-transform mid-infrared spectroscopic techniques for analysis. Fifty fibromyalgia and 50 LC patients were recruited. Spectral data were split into two sets randomly to train and externally-validate a predictive algorithm for making diagnosis. The relative percentage area of each IR band in the region of 1500 to 1700 $\text{cm}^{-1}$ was taken. Chemometric analysis was then done to analyze the spectral data. | 50 subjects in the LC group (64% female) and 50 in the fibromyalgia group (100% female), and 6 controls. The deconvolution analysis of spectral data identified a unique spectral band at 1565 $\text{cm}^{-1}$, linked to glutamate which was present only in fibromyalgia patients. The Orthogonal Signal Correction Partial Least Squares Discriminant Analysis classified spectra with high accuracy and specificity in the subgroup of external validation. Differences in the population demographic characteristics and medication history potentially introduced confounding factors. |
| Fibromyalgia and LC | Haider et al. (128) | A cross-sectional study to characterize pain, fatigue, and function in individuals reporting post-COVID-19 and compare the clinical phenotype to those with fibromyalgia and ME/CFS. Included were adult individuals that self-reported a physician diagnosis of LC, fibromyalgia, and/or CFS, and were given self-report outcome measures to assess fatigue, dyspnea, pain, quality of sleep, catastrophizing, kinesiophobia, depression, anxiety, cognitive function, | 707 respondents included in the final analysis: 203 had LC, 99 fibromyalgia, and 87 ME/CFS, while the rest had more than one of these diagnoses in combination. Individuals with post-COVID-19 reported mild to moderate fibromyalgia symptom severity, with 8.4% scoring 13 or higher on the fibromyalgia severity score. The fibromyalgia severity score was significantly lower in the post-COVID-19 group when compared to all groups with fibromyalgia and was similar to those with ME/CFS. |
| | | and physical function. The study's survey was distributed by e-mail and research websites targeting patients, faculty, and staff through the University of Iowa. | Individuals with post-COVID-19 reported lower multisensory sensitivity compared to fibromyalgia and fibromyalgia+ME/CFS ($p<.001$). Overall LC respondents reported multiple symptoms that overlap with fibromyalgia and ME/CFS, but with less severe fatigue compared to ME/CFS and less severe pain compared to fibromyalgia. |
| Fibromyalgia and LC | Hyland et al. (64) | A study that conducted a symptom network analysis of LC, ME/CFS irritable bowel syndrome (IBS), fibromyalgia, severe asthma, and a healthy control group to provide insight into the etiology of medically unexplained symptoms. Participants completed a 65-item questionnaire assessing psychological and somatic symptoms, and a network analysis was conducted taking the 22 symptoms that best discriminated between the six groups. Connectivity, fragmentation, and the number of symptom clusters were then assessed to determine relationships between symptoms and underlying causes. | Among 2,164 subjects, the symptom networks of LC and ME/CFS differed. When compared to LC, there was significantly lower connectivity, greater fragmentation and more symptom clusters in ME/CFS, IBS, and fibromyalgia. Although the symptom networks of LC and ME/CFS differed, the variation of cluster content across the groups was inconsistent with a modular causal structure but rather consistent with a connectionist biological basis of medically unexplained symptoms. |
| Fibromyalgia and LC | Nuguri et al. (136) | A follow-up study of the 2023 Hackshaw et al. (135) study to differentiate fibromyalgia from LC based on a metabolic signature. Venous blood samples were collected from LC and fibromyalgia patients using both dried bloodspot cards and volumetric absorptive micro-sampling tips. Data were acquired using surface-enhanced Raman spectroscopy (SERS). | Amide groups, aromatic and acidic amino acids patterns could help discriminate between fibromyalgia and LC. The study demonstrates the potential of SERS in identifying unique metabolites that can be used as spectral biomarkers to differentiate fibromyalgia from "LC". |
| Fibromyalgia and LC | Saito et al. (81) | A study of immunological dysregulation, chronic inflammation, and impaired erythropoiesis in LC individuals compared to recovered and healthy individuals, that used two independent cohorts of LC patients. Cohort 1 was from a previous study, while participants in cohort 2 were chosen if they had LC with ME/CFS symptoms. Clinical evaluation involved serial assessments using questionnaires such as the De Paul Symptom Questionnaire (DSQ), FACIT Fatigue scale, and Cognitive Failure Questionnaire (CFQ) Fibromyalgia (FM) diagnosis was based on ACR criteria, using the Widespread Pain Index (WPI) and Symptom Severity (SS) scale. The study used peripheral blood mononuclear cells (PBMCs) isolated for flow cytometry analysis, clinical tests (CBC, CRP, autoantibodies), cytokine and | The LC and recovered (R) groups were well-matched in age. Hospitalization rates during acute phase were 22.6% LC vs.16.6% R in cohort 1 and 11.8% LC vs. 11.7 % R in cohort 2. Comorbidities in cohort 1 were 15.9% and 16.6 % for LC and R, respectively. In cohort 2, co-morbidities were 8.8% for LC and 5.8% for R. The odds of females having LC were 3 times higher than males in both cohorts. The study noted that the majority of patients with LC suffered from comorbid fibromyalgia (72.7% in cohort 1 and 67.6% in cohort 2) and cognitive dysfunction. The LC group showed a relative increase in absolute neutrophils and monocytes but a decrease in lymphocyte counts. There was a significant reduction in the absolute number of naïve T cells in the LC cohorts, indicating selective T cell exhaustion with reduced naïve but increased terminal effector T cells. Pro-inflammatory |
| | | chemokine multiplex analysis, and ELISA assays. Statistical analysis included Wilks-Shapiro test, Mann-Whitney U test, and Kruskal-Wallis ANOVA | cytokines/chemokines were significantly elevated in both LC cohorts. LC was associated with elevated levels of plasma pro-inflammatory cytokines, chemokines, Galectin-9 (Gal-9), and artemin (ARTN). The presence of autoantibodies was found in 54.5% of the first LC cohort and approximately 55.8% of the second LC cohort. Multiple regression model revealed an increase in CD4TE, ARTN, CEC, Gal-9, CD8TE, and MCP1, and a decrease in TGF-β1 and MAIT cells that distinguished LC from the recovered group. |
| Fibromyalgia and LC | Zhang et al. (77) | Using microarray data of blood transcriptome from 75 fibromyalgia patients (GSE67311 dataset) and 29 covid-19 patients (GSE177477 dataset), authors sought to identify differential expression among the groups and potential drug targets. Machine learning was used for identifying key diagnostic genes for COVID-related fibromyalgia (LASSO algorithm and random forest in packages of R software) | Pathways of neuroactive ligand receptor interaction, ECM receptor interaction, and calcium signalling were significantly activated in the fibromyalgia group. The authors of the study concluded that these findings provide new evidence for the central sensitization hypothesis. Further analysis linked the differentially expressed genes to multiple various signalling pathways. Some commonality in differentially expressed genes between fibromyalgia and covid-19 was found. A diagnostic nomogram was developed (area under the curve of the receiver operating characteristic was 0.746) but was not validated on a new external sample of patients. |
| Fibromyalgia in rheumatoid arthritis during pandemic | Foti et al. (122,183) | A population of Italian patients diagnosed with pre-existing rheumatological disease (rheumatoid arthritis (RA) or psoriatic arthritis) was screened for fibromyalgia via The Fibromyalgia Rapid Screening Tool questionnaire and assessed for pain, depression, anxiety, and disease impact using questionnaires, during the lockdown period of the COVID-19 pandemic via telemedicine. | High rates of fibromyalgia (21.1% in RA and 24% psoriatic arthritis) were found. Patients with a fibromyalgia positive screening had a higher median RA impact of disease. These figures are similar to those known in literature regarding fibromyalgia's increased prevalence in RA populations. |
| Fibromyalgia in rheumatoid arthritis during pandemic | Upadhyaya et al. (130) | A cross-sectional study aimed to evaluate the rates of depression, anxiety, and fibromyalgia using questionnaires, among a group of RA adult patients in New Delhi, India, between June 2020 and June 2021. Fibromyalgia was assessed using the Polysymptomatic Distress Scale (range 0–31) (as the sum of the WPI and SSS). | 200 patients were included. Comorbid fibromyalgia with RA was associated with more disease activity, less remission, more functional disability, and poorer quality of life. Although it is known that fibromyalgia is more common in populations with rheumatological diseases, the study found that 31% of RA patients had fibromyalgia compared to 4% of the control group, which is higher than the 15-21% prevalence reported in pre-pandemic literature for RA (130). |
| LC in rheumatoid arthritis patients | Michaud et al. (120) | A study of RA patients with physician-diagnosed RA and self-reported COVID infections evaluated at 6-month intervals. Database used for medical and demographic data. Questionnaires were | LC in the RA population was found to be associated with severe acute COVID-19, use of antibiotics, more severe RA disease, fibromyalgia prior to infection, other comorbidities, and hospitalization for |
|  |  | used to assess depression, anxiety, fibromyalgia, fatigue, pain, and sleep problems. (Congress abstract) | COVID-19. Pre-existing fibromyalgia was not found to be statistically significant in the multivariate regression model while age, number of infections, pain, and depression were significantly associated with LC. |
| Vaccination | Di Stefano et al. (167) | Due to reports that vaccination might trigger harmful effects on the somatosensory nervous system, the authors investigated the relationship between adverse effects of coronavirus vaccination, quantitative sensory testing (QST), autonomic symptoms, and small fiber pathology on skin biopsy. They recruited adult female patients between January and June of 2022 that experienced generalized sensory symptoms and pain as long-term complications after COVID-19 vaccination (for more than 6 months). Skin biopsy was taken from the distal leg to calculate intraepidermal nerve fibre density according to the European Federation of Neurological Societies and Peripheral Nerve Society guidelines. | 15 female individuals (mean age 48.5 years), most of them had received mRNA vaccine and did not have a previous diagnosis of chronic pain or fibromyalgia, or concomitant peripheral or central nervous system diseases, experienced generalized sensory symptoms and pain and fibromyalgia-type features after vaccination. eleven met diagnostic criteria for fibromyalgia. Orthostatic intolerance was found in the majority. Nerve conduction studies were unremarkable, and most participants had normal QST. They were also found to have a normal skin biopsy. |

Goudman et al. (2021) (146) from Belgium conducted a cross-sectional study by online survey distributed through social media to investigate the possibility of “central sensitization symptoms” (i.e., fibromyalgia-type features) following COVID-19 infection. They used three validated questionnaires and assessed the impact of chronic pain, health-related quality of life, and functional status. Among approximately 500 respondents who self-reported a “post-COVID infection state” (86% females, mean age 46.5±11.4, mean time since COVID-19 was 287±150 days), 70 percent had a score consistent with fibromyalgia features (central sensitization inventory score ≥40), and more than 90 percent were classified as medium to high level of “central sensitization-related” symptom severity. A positive correlation was found with both BMI and the time elapsed since infection. The authors also found a significant correlation between central sensitization inventory scores and post COVID-19 functional status scores (F = 46.17, *p* < 0.001) in a one-way ANOVA test among 486 individuals. However, this finding is expected because the items of both these questionnaires inquire into overlapping manifestations of chronic pain/fibromyalgia-type clinical impact, and do not necessarily reflect two separate variables, which seems to raise an intrinsic problem in the study’s analysis stage.

Bierle and colleagues (2021) from the Mayo Clinic in Minnesota developed clinical criteria to diagnose patients with “post-COVID syndrome” as a syndrome consistent with central sensitization, through a modified Delphi process (144). Using their new developed diagnostic/screening method, they identified new-onset “central sensitization characteristics” (i.e., persistent new-onset fibromyalgia features) in 9% of patients that scheduled an appointment in the general context of a coronavirus infection, from November 2019 to early May 2020. These patients, if after a comprehensive evaluation, are shown to have no objective evidence of organ dysfunction, would be suitable to be diagnosed with LC, according to Bierle et al. Jennifer et al. (2023) analysed data from a large healthcare database (∼2.5 million patients) in a retrospective cohort study and compared COVID-19-positive patients to matched COVID-19-negative individuals based on medical records and healthcare utilization history. The incidence of several medical conditions was the outcome measured between date of recuperation from COVID-19 and end of study period (May 2021). Fibromyalgia incidence was found to be slightly higher after COVID-19 (0.28% new cases compared to 0.24% in controls, p=0.034 in non-hospitalized cases) (129). However, considering that delayed diagnosis of fibromyalgia is extremely common (30), reaching 6.4 years or even longer since the initial onset of symptoms according to a 2018 study (222), the above findings likely do not reflect the actual true state of fibromyalgia incidence after COVID-19.

Next, Shani et al. (2024) (134) conducted a retrospective cohort analysis using a large database of electronic medical records to investigate relationships between the BNT162b2 vaccine, SARS-CoV-2 infection, and the onset of immune-mediated diseases. Follow-up periods ranged from 4 to 12 months for vaccinated individuals and up to 16 months for those infected with SARS-CoV-2. The study defined its outcomes as the first diagnosis of an immune-mediated disease, identified through ICD-9 codes and diagnostic descriptions. According to their results, vaccination did not affect new diagnosis of fibromyalgia in any age group. On the other hand, patients aged 45–64 years or older who were infected with SARS-CoV-2 had a significantly increased risk for new diagnosis of fibromyalgia within the timeframe of the study. Specifically, the incidence rate of fibromyalgia in those infected with SARS-CoV-2 aged 45-64 was 587.9 per 100,000 person-years, compared to 313.2 in those not infected. In those aged 18-44, the incidence was 259.7 per 100,000 person-years, compared to 157.7 per 100,000 person-years in individuals not infected. These findings are translated to hazard ratios (HR) of 1.71 (95 % CI: 1.31–2.22) in the 45-64 age range, and HR of 1.72 (95 % CI: 1.36–2.19) for individuals in the age group of 18-44.

Nevertheless, Sørensen et al. (2022) (65) in their nationwide questionnaire study found conflicting results: the risk for fibromyalgia was not found to be significantly different between infected and uninfected individuals, amounting at 1% in COVID positive compared to 1.1% in COVID negatives (risk difference 0.02 95% CI: −0.09-0.14).

Studies that specifically recruited LC populations provide some more insight. Bileviciute-Ljungar et al. (2022) (16) and Scherlinger et al. (2021) (118) found high rates of positive fibromyalgia diagnosis/screening (using the ACR criteria or the FiRST questionnaire) among LC individuals. In both these studies, fibromyalgia rates were as high as 40% and 56.7%, respectively. Remarkably, of the 40% of those who fulfilled criteria for fibromyalgia in Bileviciute-Ljungar et al.’s study, 55% indicated being healthy before their infection.

In a widely cited study by Ursini et al. (2021) from Italy, which collected data via an online survey distributed among adult individuals (≥18 years) who developed COVID-19 three or more months before the survey publication (a total of 616 eligible individuals completed the survey, 77.4% women), 30.7% fulfilled the ACR survey criteria for classifying fibromyalgia 6 month on average after contracting COVID-19. Only 23 of 616 had a pre-Covid diagnosis of FM. Fibromyalgia was associated with a more severe acute infection (hospitalization), obesity, and with males. The survey was distributed on social network and was therefore subject to self-selection bias. Given these methodological constraints, there is restricted generalizability from the findings. Miladi et al. (2023) report high rates (19%) of post-Covid fibromyalgia as well in their study in Australia using a fibromyalgia screening questionnaire (131).

Myofascial-pain-focused studies: case reports indicate the development or worsening of myofascial pain and localized trigger points following COVID-19, and responding to interventions like trigger point injections and dry needling (19,162). Few population-based studies are found (163,223), but considering that gross changes in health system capacity and resources, and individuals’ behavior had also changed during this time in relation to access to primary care, lifestyle, etc., drawing conclusions is limited.

Di Stefano et al. (2023) (167) observed new-onset fibromyalgia-type features in 15 women after COVID-19 vaccination, eleven out of them met diagnostic criteria for fibromyalgia. Orthostatic intolerance was found in the majority. Nerve conduction studies were unremarkable, and most participants had normal quantitative sensory testing (QST). They were also found to have a normal skin biopsy post-vaccination.

In summary, several studies evaluated fibromyalgia prevalence/incidence after COVID-19 or as part of LC by using patient self-reported surveys and/or electronic healthcare databases (for further elaboration see Table 3). Several of the online survey studies recruited patients by self-selection and involve crucial biases that limit the generalizability of the findings, as well as possible confounding factors. Studies using data from healthcare databases (e.g., confirmed diagnosis in medical records) should consider the effect of gross changes in health system capacity and resources, and individuals’ behavior change during the pandemic in relation to access to primary care, lifestyle, and more, and that some infected individuals did not necessarily undergo PCR testing. Based on the available evidence, and according to descriptions in the literature, fibromyalgia features seem to be more frequent after COVID-19 and are consist with the previously known clinical overlap between post-viral-infection syndrome, ME/CFS, and fibromyalgia, though the findings on incidence and prevalence rates differ significantly between studies. These discrepancies can be due to differing inclusion criteria, study population characteristics and comorbidities, hospitalization status, control group chosen, outcome measures, definition used for LC, period of the pandemic and sars-cov-2 variants, and more. Another topic receiving relatively more attention in the literature was fibromyalgia in rheumatoid arthritis patients after SARS-CoV-2 infection (Table 3). It is well known that fibromyalgia syndrome often occurs concomitantly with inflammatory rheumatological disease - this has been termed by authors as “secondary fibromyalgia.”

#### 3.3.2. Observational studies on LC, chronic fatigue syndrome, and overlapping fibromyalgia (molecular mechanisms, laboratory investigations, and others)

Acknowledging the evident similarity between LC and fibromyalgia, Hackshaw et al. (2023) (135) from Texas, US, set up a pilot study to compare the low molecular weight fraction (aromatic amino acids and peptide backbones) in blood samples of fibromyalgia and LC patients using spectroscopic techniques. The fibromyalgia pattern was linked to the presence of side chains of glutamate at the bands centered at 1560 and 1579 cm^−1^. Even though confounding factors were identified, such as the use of medications in the patient group and a difference in the populations characteristics, and the relatively small sample size questions the strength of their results, it shows a potential for the development of objective diagnosis-specific biomarkers in the future. The group’s research has since been carried further (136).

Although not a study of LC per se, Das et al. (2022) undertook an impressive effort to try to uncover genetic components of chronic fatigue syndrome drawing on samples from the UK Biobank (61). By using a genome-wide association study and a combinatorial approach to analysis, they identified approximately 200 single nucleotides polymorphisms (SNPs) from 2,382 mostly European ME/CFS individuals (by self-reported diagnosis, most of them were in the age range of 61-80 years). When analysed, the total sampled population showed clustering into subgroups that seem to be associated with different phenotypes of ME/CFS. Biological processes suspected to be involved in the genome locations of the SNPs identified included, though aren’t limited to-metabolism, mitochondrial function, stress/infection, autoimmunity, sleep and the circadian rhythm, GABA synthesis, exocytosis, and synaptic vesicle cycle. Few of the SNPs had overlap or known association with other medical conditions including connective tissue diseases, fibromyalgia, multiple sclerosis, and post viral fatigue syndrome (61).

Continuing with another discipline, a 2024 psychology-oriented study aimed to explore the possible association between personality profiles and LC in non-hospitalized non-severe cases, speculating that distinct patterns of coping mechanisms or traits could characterize individuals with LC or render them more susceptible to the syndrome. An association was found between more pronounced fibromyalgia features, a higher burden of depression and anxiety, diffuse pain, attention deficit, memory problems, headaches, perception of lower quality of life, and type D personality (133). Nevertheless, the directionality of such associations should be clarified in the future, since social withdrawal and new anxious and neurotic behaviour due to uncertainty regarding chronic disease and functional impairment might help explain the observed association between high scores on DS-14 questionnaire and new-onset chronic pain.

### 3.4. Central and peripheral nervous system abnormalities

The neurological aspects of LC are another focus of research aiming to elucidate the persistent and often debilitating symptoms experienced by a substantial number of individuals who contracted COVID-19. Table 4 below summarizes the findings on this topic. Nerve conduction studies are being used for investigating neurological involvement in LC and, mostly did not reveal significant abnormalities in non-hospitalized LC. Corneal analysis revealed pathological findings (as elaborated in Table 4). Conditioned pain modulation (CPM), temporal summation, and other neurological abnormalities that may have relevance to the hypothesis of central nervous system sensitization in LC are another topic of ongoing investigations (75,148).

**Table 4:**
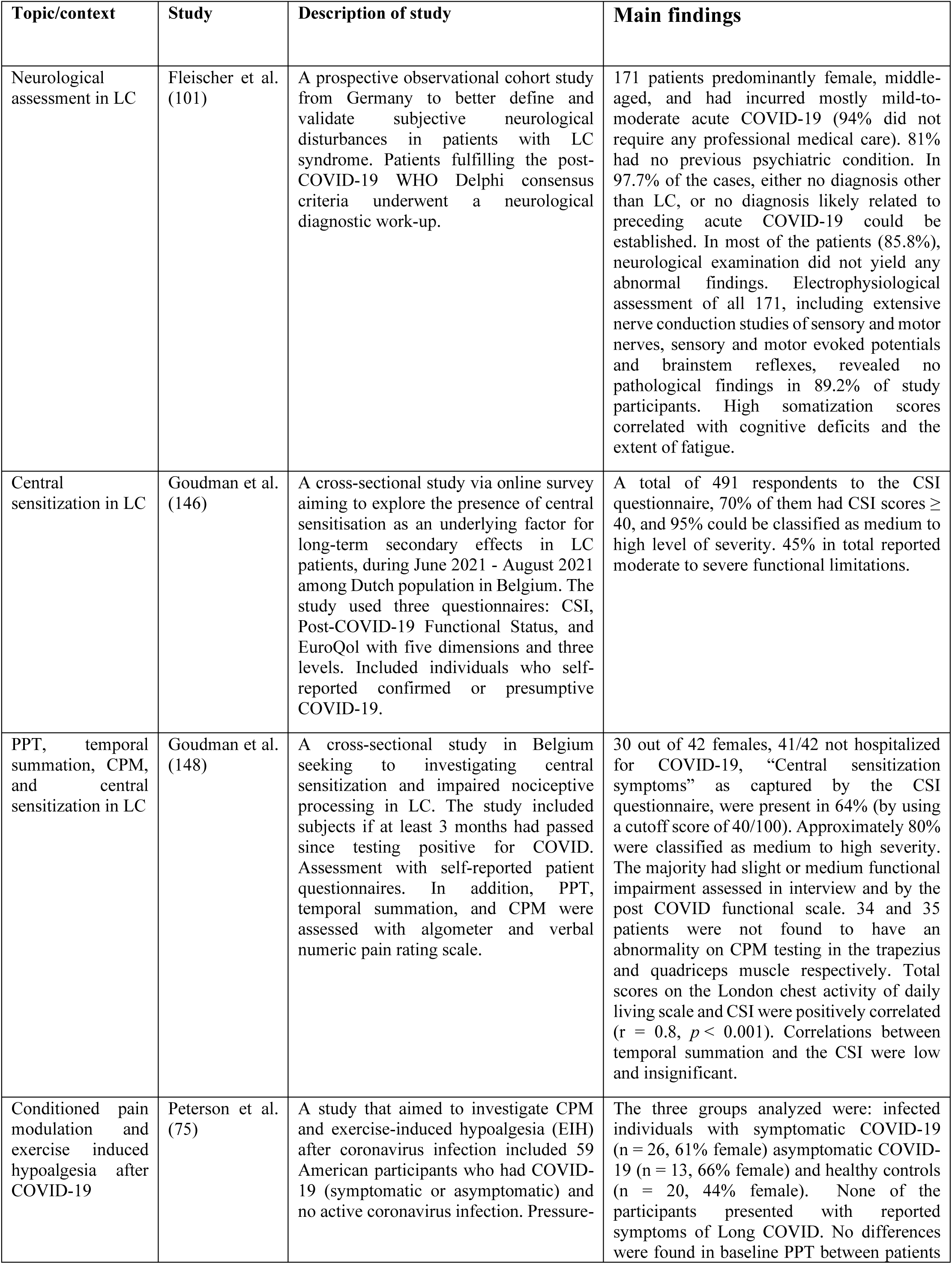
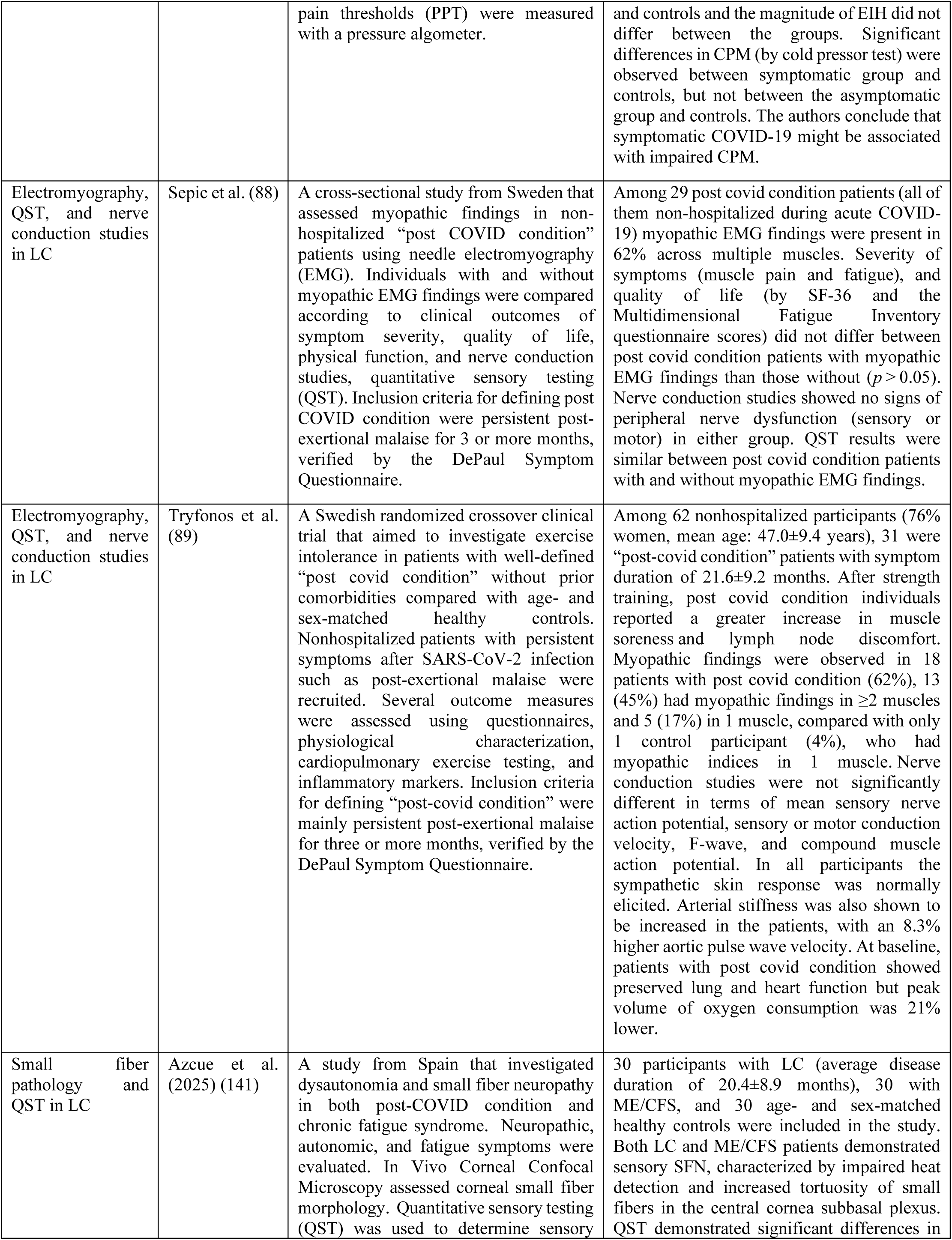
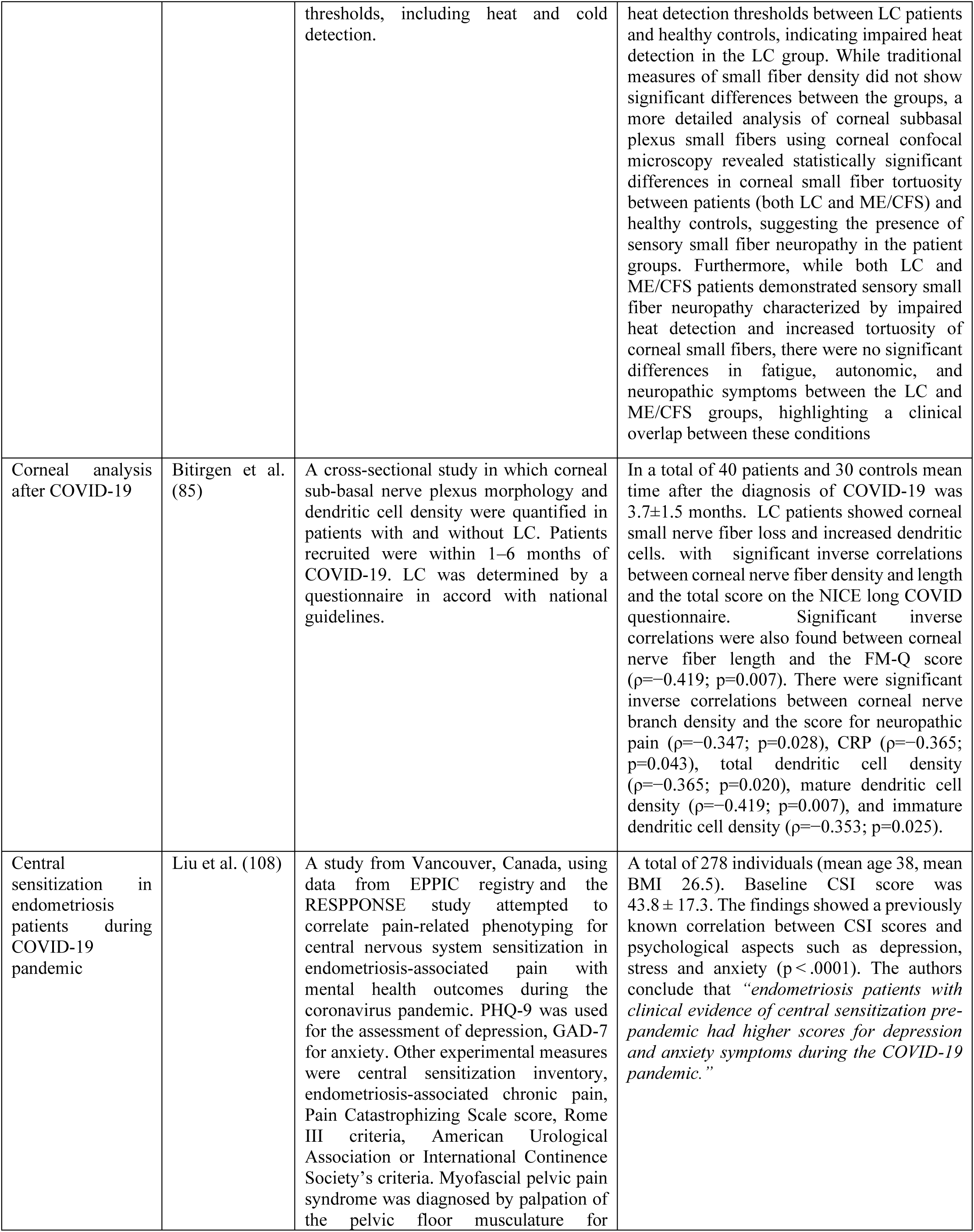
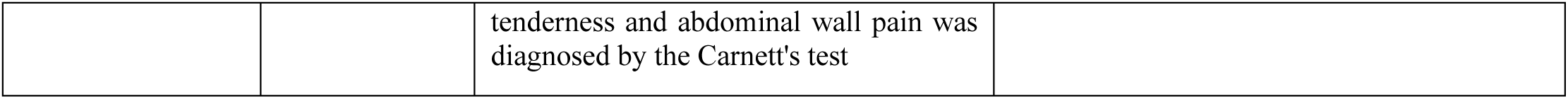
Summary of neurological studies of LC that were included in the systematic review.

However, CPM might be altered after symptomatic COVID-19 even in the absence of long COVID-19, as reported by Peterson et al. (2022) (75).

Overall, while some studies suggest the presence of peripheral nerve abnormalities in certain individuals, others, like the ones involving recuperating non-hospitalized patients with myopathic EMG findings, did not find a clear correlation with symptom severity. Azcue et al. (2025) (141) found that QST demonstrated significant differences in heat detection thresholds between LC patients and healthy controls, indicating impaired heat detection in the LC group. Cold detection showed a different trend. This variability highlights the need for researchers to employ a range of diagnostic techniques. So far, the findings appear to be heterogeneous, possibly reflecting the diverse range of neurological manifestations and the varied populations studied under the title of “Long COVID-19 syndrome.”

### 3.5. Generalized Joint Hypermobility

Generalized Joint hypermobility (GJH) is known to be associated not only with fibromyalgia but functional psychosomatic syndromes in general, and a few authors reported a clinical pattern that they identified lately with regards to joint hypermobility and LC (126,171) based on clinical experiences, as well as a larger observational study. Gavrilova et al. (126) from Saint Petersburg of Russia described what they call a typical clinical observation of theirs, regarding post-COVID fibromyalgia syndrome. They describe a constellation of manifestations involving myalgia, a palpated fibrous cord or thickened/swollen tendons, positive antinuclear antibody test, postural orthostatic tachycardia (POT), and fibromyalgia features, in a hypermobile female patient, starting several months following discharge from hospital admission that was indicated for a non-severe COVID-19 (126).

Eccles et al. (2024) (170) from the UK sought to explore whether GJH was a risk factor for non-recovery from COVID-19 infection. GJH was determined using the 5-part Hakim and Grahame self-report questionnaire by a cut-off score of ≥2 indicating GJH. According to their findings, the presence of GJH was not specifically associated with reported COVID-19 infection risk per se but was found to be significantly associated with self-reported non-recovery from COVID-19 (OR 1.43 (95% CI 1.20 to 1.70) in their study of 2,854 subjects.

A 2024 multisite study by Grach et al. (140) from Mayo clinic compared patients diagnosed with LC and control subjects that had COVID-19 without LC diagnosis, using self-reported questionnaires. GJH was assessed according to the self-assessment 5-part hypermobility questionnaire. They found that 27 percent of LC patients screened positive for GJH compared to 10 percent of controls (*p* = 0.026). Logarbo et al. (2024) (171) noticed a similar relation to hypermobility syndrome and reported findings that are in line with this issue, as further detailed in Table 5 which summarizes studies of GJH from the systematic review.

**Table 5:** Findings regarding GJH as found in the systematic review.

| Topic/context | Study | Description of study | Main findings |
| --- | --- | --- | --- |
| GJH | Eccles et al. (170) | A 2024 UK case-control study to explore whether generalized joint hypermobility (GJH) was a risk factor for self-reported non-recovery from COVID-19 infection. Data was collected through a mobile health application developed with input from physicians and scientists at King's College London, Lund and Uppsala Universities, and Massachusetts General Hospital. The 5-part Hakim and Grahame self-report questionnaire were used to determine GJH by a cut-off score of $\geq 2$ indicating GJH. Based on regression analysis they determined if the presence of GJH was a predictor of non-recovery. | Among individuals reporting incomplete recovery from COVID-19 infection (n=914), 269 patients (254 (29.4%) female) had GJH. In the fully recovered group, 439 of 1940 patients (22.6%, 400 female) had GJH. While the presence of GJH did not show a specific association with COVID-19 infection risk itself, it was significantly associated with non-recovery from COVID-19 (OR 1.43 (95% CI 1.20 to 1.70)). GJH significantly predicted higher fatigue levels, the latter which mediated the link between GJH and non-recovery from COVID-19 (170). |
| GJH | Gavrilova et al. (126) | Case report | Young female patients with joint hypermobility that develop new manifestations after discharge from hospital for COVID-19, including myalgia, postural orthostatic tachycardia (POT), antinuclear antibodies, a noticeable thickened/swollen tendon, and new onset fibromyalgia-type features. |
| GJH | Grach et al. (140) | The objective of this study was to ascertain if individuals diagnosed with LC exhibited a higher incidence of new or exacerbated health conditions. The cohort comprised patients diagnosed with LC at Mayo Clinic facilities in Rochester, Minnesota, and Jacksonville, Florida. Controls had a confirmed history of SARS-CoV-2 infection but had not received a diagnosis of LC. Questionnaires were sent to LC patients and controls to assess new or worsening comorbidities following COVID-19. The questions screened for ME/CFS, GJH, and orthostatic intolerance. The self-assessment 5-part hypermobility questionnaire was used for assessing GJH. | 247 respondents of LC group and 40 controls. The mean age of controls was higher ($p = 0.021$). 94.4% of LC individuals reported that they had a pain condition that worsened or started after COVID-19 compared to 0% of non-LC controls. 58.3% of LC patients met the criteria for ME/CFS, compared to 0% of controls. 27.0% of LC patients had GJH (out of them 90.5% female), compared to 10.3% of controls. LC patients showed significantly higher scores for orthostatic intolerance compared to controls. LC respondents were also found to be more likely to report sensitivity to medications (22.5% vs. 4.9%, $p = 0.007$). Other reported findings in LC group included neurological (92.4%), sleep (82.8%), skin (69.8%), genitourinary (60.6%), allergies/sensitivities/intolerances (31.8%), mood disorders (31.6%), gastrointestinal disorders (21.9%), viral reactivation (e.g., Epstein-Barr virus, herpesvirus, or other viruses) (18.7%), autoimmune diagnoses (16.2%), pulmonary disorders (15.8%), and cardiac disorders (12.1%). |
| GJH | Logarbo et al. (171) | Case report | Five female patients, aged 33 to 51 years, with no known history of hypermobility, presented with persistent and debilitating fatigue, cognitive dysfunction, dysautonomia, and diffuse joint pain, among other neuromusculoskeletal symptoms, over a period of 3 to 15 months<br>following acute SARS-CoV-2 infection. Clinical evaluation confirmed generalized joint hypermobility using the Beighton Score, receiving scores at or above the age-appropriate diagnostic threshold. Notably, all patients exhibited C677T or A1298C polymorphisms in the methylenetetrahydrofolate reductase (MTHFR) gene, which was previously shown to be linked to the development of hypermobile Ehlers-Danlos Syndrome and hypermobility spectrum disorder. Given that up to 35% of the general population carries one of these MTHFR polymorphisms and hypermobility of any etiology may affect up to 57%, the findings suggest a possible predisposition unmasked by SARS-CoV-2 infection (171). Pathophysiological mechanisms suggested may include immune dysregulation, mast cell activation, and connective tissue inflammation. Patients were managed with methylated folate and B12 supplementation, physical therapy, and mast cell stabilization strategies. |

### 3.6. Studies on Interventions

For publications that are dedicated to the topic of LC treatment, the reader is referred to literature on LC interventions (51,80,186,224–226). Although the focus of the work was not to specifically review treatments for LC, and the Cochrane database was not used, a brief overview of treatments that were encountered during the review process is given as follows:

Treatments that were either discussed or investigated in the context of LC or post-viral fatigue syndrome can be categorized into (i) non-pharmacological, (ii) pharmacological and nutritional, (iii) other interventional, (iv) and multimodal. These include: (i) mindfulness training (172), cognitive behavioral techniques (137), pain neuroscience education (110), balneotherapy (149), (ii) creatine (56), Ginseng (151), coenzyme Q10 and alpha lipoic acid supplements (155), L-arginine and vitamin C supplementation (80), synbiotics SIM01 or other supplement/probiotic complexes (159), melatonin (58), low-dose naltrexone (168), metformin (150), antivirals or other immune modulators (51,117), metabolic and other miscellaneous drugs such as colchicine and antihistamines (117), (iii) transcutaneous electrical nerve stimulation (156), pulsed electromagnetic field treatment or other electrophysiological method (161), plasmapheresis (51), hyperbaric oxygen therapy (157,165), (iv) L-acetyl-carnitine in combination with physical rehabilitation (154), multidisciplinary rehabilitation (153,186), multimodal ME/CFS-directed therapy (160), some of which were in non-blinded/non-randomized/non-controlled exploratory studies. While some of these publications reported overall positive results in the short term, others are more speculative.

The following treatments were documented in a follow-up survey of patients attending the interdisciplinary post-COVID care clinic in Mayo clinic of Minnesota (48), which offers a multidisciplinary approach to the treatment of LC (combining use of off-label medications and nonpharmacologic rehabilitative approaches, many of which have previously been used for POT syndrome, ME/CFS, and fibromyalgia) for evaluating the response to different treatments. These treatments were either prescribed or self-used by patients: non-pharmacologic interventions (e.g., a “post COVID treatment program,” physical therapy, occupational therapy, biofeedback), off-label medications (low-dose naltrexone, propranolol, gabapentin, pregabalin, amitriptyline, guanfacine, N-acetyl cysteine, colchicine, midodrine, fludrocortisone, aripiprazole, pyridostigmine, etc. while additional therapies that physicians have been offering more recently, such as guanfacine, NAC, and L-arginine, were not captured in the survey), supplements (fisetin, coenzyme Q10, ginseng, ashwagandha, Reishi mushrooms, nattokinase, specialized pro-resolving mediators), and other interventions (plasmapheresis, transcranial magnetic stimulation, vagal nerve stimulator). These are mentioned here more as initial anecdotal evidence for investigators or stakeholders that may wish to study them further in the context of LC.

Meanwhile, Blanchard et al. (2022) developed a mobile health application for fibromyalgia-type LC (164,173,177). Bileviciute-Ljungar et al. (2022) (16) from Sweden’s Karolinska Institute recruited individuals for their study of a multidisciplinary rehabilitation program, focusing on those with functional impairment and persistent symptoms after COVID-19. In their cohort of 100 individuals with post COVID-19 functional impairment, 40% met criteria appropriate for fibromyalgia diagnosis, while 68% reported being completely healthy before COVID-19. In their randomized controlled study of an eight-week telerehabilitation program, they report positive results for functional status, activity, pain, and health-related outcomes compared to the waiting list group after six months (152,153).

Kjellberg et al. (2022) set up a double-blinded randomized controlled clinical trial to evaluate the therapeutic efficacy of hyperbaric oxygen therapy in LC patients who were healthy prior to COVID-19 (165). They have yet to publish their findings at the time or writing. Meanwhile, Zilberman-Itskovich et al. (2022) (157), in a randomized sham-controlled trial with 37 LC participants (of which only 10.8% were hospitalized during COVID-19) reported an improvement in clinical outcomes (sleep, memory, information processing, pain interference, anxiety, somatization, energy, total taste score, health-related quality of life) and brain perfusion, after hyperbaric oxygen therapy. Data were collected at 1-3 weeks after completion of 40 daily sessions, five sessions per week, with 100% oxygen by mask at a pressure of 2 ATA.

Interestingly, turning to a study on prevention, in a 2024 multicenter randomized clinical trial by Bramante et al. (150), metformin, given to patients in the COVID-19 outpatient setting, was shown to reduce risk of developing LC when assessed at 300 days. The study included adults (30-85 years-old) with overweight or obesity and found a reduced incidence of LC diagnosis by approximately 40 percent (absolute reduction of 4.1 percent) compared with placeb. However, the HR did not appreciably change when adjusted for other a-priori baseline variables. In their study, LC was primarily ascertained by participant-reported receipt of a long COVID diagnosis from a medical provider.

In summary, a wide range of potential treatments for LC are being investigated, encompassing non-pharmacological, pharmacological, interventional, and multimodal strategies, reflecting the syndrome’s complex and heterogeneous nature. While many treatments remain speculative or require further rigorous study, preliminary findings suggest some benefit from interventions such as multidisciplinary rehabilitation, probiotic supplements, and hyperbaric oxygen therapy, which seem to improve symptoms such as fatigue, cognitive and neurological symptoms, and quality of life, at least when examined in the short term, highlighting the need for continued research to establish efficacy and optimize management strategies.

### 3.7. Reviews (systematic reviews, meta-analyses, and narrative reviews)

Table 6 summarizes the systematic reviews included in this scoping review. Particularly germane are the following:

- Fowler-Davis et al. (2021) conducted a systematic review of studies of interventions for post-viral fatigue (59). They found a range of treatment modalities that have been studied so far but conclude that more research involving heterogenous populations is needed to properly assess their effectiveness in the context of post-viral fatigue syndromes.
- Cohen and colleagues (2022) published a comprehensive review on the relationship between chronic pain and infections, elaborating on mechanisms that could be relevant to LC-associated pain (92).
- Rao et al. (2022) conducted a systematic review and meta-analysis (41 studies, 9,362 patients in total) to evaluate the prevalence and prognosis of post-COVID-19 fatigue (60). They found that fatigue prevalence was 44.9% (95% CI 0.329 - 0.575, *I*^2^ = 70.57%) within the first 3 months post-recovery according to a small number of relevant studies, but substantial differences existed among studies. Female patients, inpatient setting, and individuals recruited through social media and in Europe had a higher prevalence of fatigue.
- A systematic review and meta-analysis by Kerzhner et al. (2024) (124) sought to evaluate rates of LC’s persistent pain manifestations, as well as the impairment to health-related quality of life and data on laboratory inflammatory markers in LC. In their analysis, a substantial level of heterogeneity was found and funnel plots demonstrated considerable asymmetry. The pooled proportion of individuals experiencing general body pain symptoms up to one year after COVID-19 acute phase resolution was found to be higher in the nonhospitalized compared to hospitalized individuals (0.306 vs. 0.089, respectively, I^2^ = 95%, p_(subgroup)_ = 0.009). They also discuss the increased associations related to young age, females, and less severe acute COVID-19, as well as a progressive temporal-proportional trend instead of the usual subsiding nature of most other symptoms (124). On that note, Ebbesen et al. witnessed a similar trend in their findings from a nationwide cross-sectional study (105).
- A systematic review and meta-analysis by Hwang et al. (2023) (62) appraised viral infections as an etiology of ME/CFS.

**Table 6:** Overview of systematic reviews included in the scoping review.

| Study<br>(year) | Databases used | Topic |
| --- | --- | --- |
| Fowler-Davis et al. (2021) (59) | CENTRAL, CINAHL, MEDLINE (EBSCO), ProQuest (APA PsycINFO), SCOPUS, SportDISCUS, the International Clinical Trials Registry Platform (World Health Organization) the UK Clinical Trials Gateway (NHS, National Institute for Health Research). | Interventions for fatigue and post-viral fatigue |
| Arienti et al. (2022) (70) | Cochrane Systematic Reviews | Fatigue, post-exertional malaise and orthostatic intolerance |
| Cohen et al. (2022) (92) | MEDLINE, Embase, OVID, and Google Scholar | Comprehensive review on the relationship between chronic pain and infection (mechanisms, causes, treatments, and more) |
| Fernández-de-Las-Peñas et al. (2022) (116) | MEDLINE, CINAHL, EMBASE, Web of Science, medRxiv and bioRxiv | Prevalence and time-trends of musculoskeletal pain (e.g., myalgia, joint pain, and chest pain) after COVID-19, in hospitalized and non-hospitalized cases (systematic review with meta-analysis) |
| Kocyigit & Akyol | Web of Science, Scopus, and MEDLINE | The COVID-19 pandemic and fibromyalgia - the review focuses on prevalences, the suggested relevance of |
| (2022)<br>(143) |  | psychological and psychosocial factors, the role of inflammation, and vaccination. |
| Pires et al.<br>(2022)<br>(106) | MEDLINE (via PubMed) and Google Scholar | Musculoskeletal manifestations of acute and long COVID-19 and the effect of COVID-19 on bone, joints, muscle, rheumatological diseases, and susceptibility to musculoskeletal infection |
| Rao et al.<br>(2022) (60) | MEDLINE, Embase, PsycINFO, CINAHL, Web of Science, Scopus, trial registries (i.e., NIH clinical trials registry, Cochrane Central Register of Controlled Trials, and ISRCTN registry), and Google Scholar. Pre-print servers (MedRxiv and Psycharxiv) were also included and a manual search of OpenGrey. | Post-COVID-19 fatigue (a systematic review and meta-analysis) |
| Yong et al.<br>(2022) (11) | PubMed, SCOPUS and Web of Science | Proposed subtypes of LC: non-severe COVID-19 multi-organ sequelae, pulmonary fibrosis sequelae, ME/CFS, POT syndrome, post-intensive care syndrome, and medical or clinical sequelae |
| Ciaffi et al.<br>(2023)<br>(107) | MEDLINE and Web of Science | Post-acute COVID-19 musculoskeletal manifestations review aimed for rheumatologists |
| Hwang et al. (2023)<br>(62) | MEDLINE and Cochrane Library | Viral infections as an etiology of ME/CFS (systematic review and meta-analysis) |
| Lewthwaite et al.<br>(2023)<br>(158) | MEDLINE, Embase, CINAHL, Scopus and Cochrane | Two reviews: the first is an umbrella review of systematic reviews of LC symptoms, prevalence, and complications grouped into eight treatable trait clusters. The second is an umbrella systematic review of randomized control trials of interventions for LC prevention or management. |
| Kerzhner et al. (2024)<br>(124) | MEDLINE and EMBASE | Pain in LC after hospitalized and non-hospitalized COVID-19 (a systematic review and meta-analysis) |
| Silva-Passadouro et al.<br>(2024) (24) | MEDLINE, Embase, CINHAL, PsycINFO and Web of Science | Resting-state surface quantitative EEG findings in fibromyalgia, ME/CFS and LC compared to healthy controls. |
| Skare et al.<br>(2024) (94) | Pubmed, Scielo and Embase | Ear abnormalities in fibromyalgia, ME/CFS, COVID-19, LC, POT syndrome, and more related syndromes |

Narrative (non-systematic) reviews and topical articles were the most frequent type. Paroli et al. (2024) (79) detail the potential role of infection and inflammation in the etiopathogenesis of fibromyalgia and LC, and Choutka et al. (2022) (13) summarize the current understanding of unexplained post-acute infection syndromes, covering epidemiology, signs and symptoms, patho-mechanisms, and prognosis. A 2024 publication by Stefanou and colleagues (80) gives an updated overview of the neurological and psychiatric manifestations of LC, analysing recent advances in understanding its pathophysiology and the clinical presentation (including prevalence, risk factors, and temporal dynamics of neurological aspects), in addition to covering issues of prevention and vaccination, and exploring potential diagnostic and treatment implications. The authors also propose a standardized framework for the clinical approach and management of patients who have neurological manifestations of LC, plus recommendations for future research. They emphasize that in the absence of a standardized diagnostic framework, the development of comprehensive clinical practice guidelines is significantly impeded. They also stress that a thorough and interdisciplinary assessment of persistent symptoms after COVID-19 is imperative to exclude an underlying well-defined cause (e.g., endocrine, neurological, cardiovascular, autoimmunity, respiratory) (80) in order not to erroneously attribute such symptoms to LC.

### 3.8. Findings from preprints and the narrative literature search of subtopics

As a part of the scoping review, subtopics were explored through a non-systematic search, supplemented by a targeted search of the medRxiv preprint database. Among the subtopics searched were small fibre pathology in LC, somatic symptoms, conditioned pain modulation, quantitative sensory testing, and more as elaborated in the methods section. The main findings are mentioned briefly in Table 7 which summarizes the studies found during the preprint and subtopic searches. As shown, more studies are found on GJH and LC.

**Table 7:** Findings from subtopic searches and preprints.

| Topic | Database and search phrase | Study (year of upload/publication) | Description of Study |
| --- | --- | --- | --- |
| GJH | medRxiv (“hypermobility long COVID”) | Torok et al. (2025) (227) | A 2025 preprint by Torok et al. further reveals a link between GJH and LC in their cross-sectional online survey among U.S. and U.K. populations to compare those with self-reported LC to those without, assessing generalized and extreme joint hypermobility via questionnaires. Logistic regressions were then employed, controlling for covariates, to examine the predictive relationship between hypermobility and LC. Among 1,816 respondents, 352 (19.4%) self reported LC. LC individuals reported a higher number of coronavirus infection episodes and more severe infections. In logistic regression analysis it was found that in mild-<br>asymptomatic COVID-19 subgroup, GJH was associated with LC with OR 1.54 (95% CI: 1.05-2.25 $p=0.027$) compared to OR 0.90 (0.64-1.28) in severe infection. Extreme joint hypermobility was associated with LC, with OR 2.59 (95% CI 1.37-4.89 $p=0.003$) in mild-to-asymptomatic COVID-19, whereas in the severe infection there was no statistically significant association between extreme joint hypermobility and LC. Also, 29.6% of LC individuals reported a parent, child or sibling with LC, compared to 11.7% in non-LC individuals ($p<0.001$). Overall, their analysis suggests that hypermobility influences the odds of LC by two pathways: (i) GJH increases the risk that individuals with no or moderate initial symptoms from a COVID-19 infection experience LC, (ii) GJH is a predictor of developing severe initial symptoms from COVID-19, which is independently associated with increased LC risk. |
| GJH | medRxiv<br>("hypermobility long COVID") | Eckey et al. (2025) (228) | A retrospective online survey of 3,925 patients with either ME/CFS or LC. The study aimed to identify treatments with the greatest perceived benefits for these conditions and to understand how these treatments affected different core symptoms. Ehlers-Danlos Syndrome/joint hypermobility was reported as a comorbidity in both ME/CFS (40.2%) and LC (27.2%) patient groups. When the patients were clustered into four distinct subgroups based on their symptoms and comorbidities, Cluster 2 was characterized by a "POT syndrome-Dominant Presentation". Patients in this cluster reported the highest rates of POT syndrome as a primary symptom (75.0%) and comorbidity (95.1%). Compared to Cluster 3, patients in Cluster 2 reported higher rates of certain conditions, including EDS (33.5%), dysautonomia, mast-cell activation syndrome, and craniocervical instability. This suggests an association between POT syndrome-dominant presentation and joint hypermobility within this cohort. |
| GJH | medRxiv<br>("hypermobility long COVID") | Eastina et al. (2025) (229,230) | A follow-up online survey of 526 adults aged 20-65 with a history of LC to assess the duration and severity of autonomic dysfunction and its impact on function and quality of life. Data were collected via the REDCap platform using questionnaires such as the COMPASS-31 and SF-36, and multivariable logistic regression was used for statistical analysis. This study revealed significant associations between GJH and LC-related autonomic dysfunction. A substantial portion of the LC cohort, 29.7%, self-reported joint hypermobility. Moderate to severe autonomic dysfunction (defined as a COMPASS-31 score $\geq 20$) in LC was associated with female sex ($p = 0.012$) and joint hypermobility ($p = 0.019$). Multivariable logistic regression analysis demonstrated that those with joint hypermobility had a 1.68-fold increased risk of moderate-to-severe autonomic dysfunction compared with those without joint hypermobility (OR 1.68, $p = 0.044$). Notably, joint hypermobility emerged as the most robust risk factor for developing POT syndrome following SARS-CoV-2 infection, conferring a 200% increased odds (OR = 2.05, 95% CI: 1.34-3.14, $p = 0.001$). The researchers suggest that the underlying mechanism for this association might involve connective tissue laxity in individuals with GJH, potentially leading to increased venous pooling and reduced venous return, thus contributing to postural tachycardia |
| GJH | medRxiv<br>("hypermobility long COVID") | Wilson (2023) (231) | An investigation into the overlapping clinical and genetic features of Ehlers-Danlos syndrome (EDS) and LC. This study assessed 1,261 EDS outpatients, utilizing a database of 120 history and physical findings, including genetic data from genetic sequencing in 568 of these patients. These data were then compared to 15 |
|  |  |  | cases of LC from an extensive review and to 104 genes previously associated with COVID-19 severity in previous molecular studies. The study identified six identical genes (F2, LIFR, NLRP3, STAT1, TICAM1, TNFRSF13B) and 18 similar genes (including POLG-POLD4, SLC6A2-SLC6A20, and NFKB1-NFKB2) relevant to both EDS and COVID-19 severity. |
| Somatic symptoms and COVID-19 | PubMed (“somatic symptoms COVID”) | Shevlin et al. (2020) (219) | An investigation of the association between COVID-19-related anxiety and somatic symptoms in adult UK population. Among a representative sample of 2,025 individuals, moderate to high levels of COVID-19-related anxiety were significantly associated with general somatic symptoms, particularly gastrointestinal and fatigue symptoms. This remained significant after controlling for factors such as generalized anxiety disorder. |
| Somatic symptoms and COVID-19 | PubMed (“somatic symptoms COVID”) | Liu et al. (2020) (232) | A cross-sectional survey by Liu et al. of China found that concern regarding COVID-19 was positively correlated with the occurrence of somatic symptoms in college students and primary school children (232). The incidence of somatic symptoms among college students was 34.85 (out of them 26.26% mild and 8.59% moderate). While the incidence of somatic symptoms in primary school respondents was 2.39% (all mild). |
| Fibromyalgia incidence trends 2014-2021 | PubMed (“fibromyalgia incidence nationwide”) | Lee et al. (2023) (223) | Lee et al. (2023) (223) report on nationwide data of fibromyalgia incidence from 2012 to 2021 in South Korea population. The data showed a peak in the annual incidence in the year 2019 reaching 109.20 per 100,000 persons compared to 88.07 in the year 2014. |
| Non-recovery after COVID-19 | PubMed (“long COVID”) | Peter et al. (2025) (233) | A prospective longitudinal cohort study followed LC patients into the second year since contracting COVID-19 and found that the majority of working age patients with LC, who demonstrated no major pathology in laboratory investigations, did not recover. Many remained with a non-specific ME/CFS-type clinical picture despite no obvious clinical finding and no viral persistence in blood and stool testing. |
| LC patient experiences and perspective | PubMed (“long COVID”) | Chasco et al. (2022) (234) | A 2022 study by Chasco and colleagues conducted by interviews offers valuable insight into patient perspective on the impact of the fatigue and brain-fog in LC capturing not only physical descriptions but the social, occupational, and interpersonal aspects as well. |
| LC patient experiences and perspective | PubMed (“long COVID”) | Wurz et al. (2022) (235) | Wurz et al. (2022) evaluated LC experiences, noticing that patients had a deep sense of loss over one's prior identity. In their inquiries they found four main themes (numerous and exhausting LC symptoms, pervasive LC effects, physical activity is difficult or too demanding, and asking for help when few are listening, and little is working) commonly reported by 213 participants (88.2% woman, 33.1% aged 40–49, 74% experienced LC symptoms for ≥ 6 months). |
| Skin biopsy and QST in LC | PubMed (“conditioned pain modulation”, “quantitative sensory testing” and “COVID”) | Flaco et al. (2024) (220) | A case-control study that evaluated possible small fibre neuropathy in patients experiencing painful LC. Skin biopsies were taken from 26 patients with painful LC and quantitative sensory testing was done. Both outcomes were compared to individuals with past COVID-19 infection with painless LC, which was characterized mainly by symptoms such as brain fog and fatigue, and compared also to asymptomatic post-COVID-19 non-LC controls. Among the 26 patients with painful LC, twelve had skin biopsy and/or quantitative sensory testing abnormalities compatible with small fibre neuropathy, while the rest did not. Interestingly, approximately 50% of patients experiencing painful LC had small fibre neuropathy, similar to the rates of small fiber pathology among fibromyalgia patients according to a 2018 meta-analysis (236). |
| Immunology in LC | medRxiv (“long COVID”) | Santos Guedes de Sa et al. (2024) (237) | In a preprint by a research group from Connecticut mechanisms related to autoimmunity continue to receive attention in LC research. The study investigates whether autoantibodies are causally linked to neurological symptoms in LC. Total IgG was purified from plasma of LC participants and its reactivity against human and mouse tissues was analyzed. The study found that IgG from LC patients showed increased reactivity against neural tissues such as the human pons and mouse sciatic nerve, meninges, and cerebellum. This suggests that autoantibodies targeting neural tissues may contribute to the development of neurological symptoms in LC. |

## 4. Summary and conclusions

The purpose of this systematic scoping review was to evaluate and map the body of scientific literature and evidence on new-onset fibromyalgia after non-severe non-hospitalized sars-cov-2 infection. It’s well-recognized that infectious agents such as Epstein Barr virus, cytomegalovirus, mycoplasma, Coxiella burnetii, and other pathogens can be triggers for post-infectious chronic fatigue or are associated with ME/CFS (73,86,92) but the pathobiology of such clinical manifestations is not well defined. In fibromyalgia syndrome, bacterial and viral infections, likewise, are recognized as a part of the aetiology (51,96,238). The overlap of fibromyalgia syndrome, post-viral functional somatic illness, and ME/CFS is an unsolved medical puzzle with increasing public and community health importance. Moreover, considering the empirical findings gathered in the review of literature regarding GJH, such as the results of Eccles et al. (2024) (170), hypermobility seems to be another piece in this puzzle.

Research on fibromyalgia continues to primarily focus on pain, but fibromyalgia is a syndrome. Moving towards a wider view of fibromyalgia beyond pain is needed for the scientific community’s proper comprehension of this intriguing condition. The empirical evidence supports an overlap between LC, chronic fatigue syndrome, and fibromyalgia but the molecular mechanisms still remain unclear. Mechanisms such as neuroinflammation, immune dysregulation, viral persistence, glial cell reactivity, and central pain augmentation are studied, yet are under-researched, and with conflicting findings regarding the presence of viral particles, condition pain modulation, central sensitization, autoantibodies and more. The emergence of post-COVID-19 fibromyalgia presents a significant challenge, requiring a collaborative interdisciplinary approach that integrates evolving knowledge.

Various types of methods, tools, and study designs are being used for LC research, with inconsistency in key concepts and definitions. This leads to a fragmented understanding of the relationship between SARS-CoV-2 infection and LC. Studies that recruit participants in open web-based surveys are subject to self-selection bias, therefore, many of these studies which were included in this scoping review do not allow a well-founded conclusion to be made regarding incidence or prevalence rates of new onset fibromyalgia after COVID-19. Such study designs limit the generalizability of the data greatly. Several significant biases were identified in the studies during the review, including recruitment, self-selection, and recall bias, and other methodological issues as stated in the previous sections. Based on the available evidence, there are indications that new-onset fibromyalgia occurs more frequently after COVID-19, but well-designed studies and meta-analyses are warranted to clarify this and to provide more accurate estimates of epidemiology and possible predisposing factors. The strongest evidence so far seems to be a large 2024 retrospective cohort study (134) that found hazard ratios of ∼1.7 for incidence of new clinically made diagnosis of fibromyalgia up to 16 months after COVID-19 compared to non-infected individuals in the age range of 18-64 years, regardless of vaccination status, but since diagnosis of fibromyalgia is a long process that can take several years in the community care setting, these values might conceivably be gross underestimations.

<u>Central sensitization:</u> central sensitization is one of the areas of focus in LC research. While the observed associations between SARS-CoV-2 and chronic “nociplastic”-type pain syndromes highlight a potential pathogenic link, the precise underlying molecular pathways remain poorly understood and are under continuous debate across the literature. Neurocentric hypotheses such as diffuse pain augmentation in the central nervous system interestingly do not seem to align well with empirical evidence that was found during this systematic scoping literature review of LC, specifically in studies of pressure pain threshold, quantitative sensory testing (141), conditioned pain modulation (148), and temporal summation (148), nor do such notions seem to consolidate well with initial descriptions of plasmapheresis being reported as somewhat beneficial by nine out of nine LC patients (48). But with such a paucity of evidence found (75,88,89,101,112,113,141,148,203,206,220) it seems, however, still early to draw conclusions. Be that as it may, in well-designed studies the statistics paint the scientific picture. LC could indeed be a heterogenous condition with subtypes determined by multiple over- and under-represented molecular pathways in each subtype. But if the central sensitization paradigm (38,52,108,137,139,239–242) is as an axiom, there’s no need to adjust the theory (240) or spot out anomalies (68,69,91,101,148,203,243–250).

The development of new onset musculoskeletal pain after COVID-19 is multifactorial, and can involve both direct viral causes, lifestyle and behavioral factors, other pre-existing and new comorbidities, psychological factors, and more. Having said that, simplistic hypotheses such as fatigue/pain catastrophizing, low psychological resilience, and physical deconditioning, might well explain fatiguability and exercise intolerance, but they do not seem to explain the broad symptomatology of the not-so-rare syndrome among mild non-hospitalized young and previously healthy female cases. Medicine, historically speaking, has had an unhealthy tendency to attribute that which it cannot comprehend to psychology, e.g., (251). An increased feeling of bodily stiffness, as seen in studies (189), doesn’t seem very intuitive when viewed from the standpoint of central sensitization.

<u>Problems in LC terminology inconsistencies</u>: there is inconsistency across literature with regards to what LC basically is, besides the choice of nomenclature itself or the number of weeks duration. Using the term “post/long/chronic covid” to refer to any declined health including that due to well defined organ damage can lead to the conflation of “Long COVID syndrome” and other known medical diagnoses. Publications using this term without parsimony will make it difficult for future researchers attempting to review and analyze the body of literature, as well as to unnecessary confusion in the field. It is the author’s suggestion that if, after anamnesis, objective organ-relevant evidence in clinical investigations can explain the dominant new symptoms of a patient, this should not be referred to as LC or any of its synonyms. LC, indeed, may occur alongside other post severe-COVID-19 organ damage, but as a post-viral chronic fatigue-like syndrome it should be differentiated from such. Since the mechanisms are still ambiguous at this time, making an official distinction between persistent symptoms based on four or twelve weeks seems arbitrary and does not abstain from “multiplication of entities” with reference to the principle of Occam’s razor. If it is a syndrome, as with other syndromes, it should be recognized as a phenomenon and diagnosed clinically based on internationally accepted criteria. Chronic breathlessness (53,93), for example, is not a syndrome, it is a presenting complaint. ‘Suspicion of LC’ could be a more accurate term to consider in future studies in case the assessment was based on anamnesis alone without objective clinical testing. Finally, coherent terminology of a responsible international health body should be used with consistency. The term “post-covid” doesn’t seem like the best choice because it is not clear enough if the speaker refers to something happening any point in time after covid infection/pandemic or the syndrome.

Author’s suggestion is: “Long COVID sequelae syndrome”-new-onset persistent medically unexplained multisymptom illness after sars-cov-2 infection, often presents clinically as ME/CFS or insidious fibromyalgia-type features. LC is a post-infection chronic condition that impairs one’s quality of life.

Prospective studies will likely help formulate a comprehensive explanation, and uncover a causal link, if there is such, between infections and fibromyalgia. Ideas such as immune dysregulation, viral persistence, pure psychosomatics, neuroinflammation, glial reactivity, and central nervous system sensitization are, as noted, postulated, but remain to be further investigated in rigorous empirical studies that are designed according to theory-based hypotheses and with reproducible data. Hence, before announcing the discovery of well documented “central pain augmentation” in LC (137,139), studies that can clearly show this, and, preferably, can clearly link the disease with underlying pathogenic mechanisms, are needed. Further research is required to elucidate the relevant neurobiological (or psychobiological) pathways and inform targeted sensible effective therapeutic strategies. Multimodal rehabilitative approaches, as well as other approaches, are being studied in these relatively early stages of LC research, some of which seem to point in a positive direction.

Knowledge gaps: The main knowledge gaps in understanding post-COVID-19 fibromyalgia that were identified from the above literature review are:

1. Etiopathogenesis and theoretical framework: unraveling the underlying pathobiology and disease mechanisms. These crucial knowledge gaps revolve around the fundamental mechanisms that drive fibromyalgia-type clinical features, the role of viral triggering, contribution of dysregulated immune pathways, genetic and epigenetic predisposition, environmental and lifestyle factors, neural mechanisms and their temporal dynamics, and the supposed role of emotional stress. Also, the current evidence concerning the impact of hospitalization history on the incidence of post-COVID fibromyalgia is inconclusive. This ambiguity is likely attributable to methodological inconsistencies, disparate definitions of fibromyalgia and LC, and variations in the selection criteria of study populations.

A comprehensive theoretical framework for fibromyalgia (and LC) is needed that goes beyond explanations focused solely on pain and hyperalgesia or fatigue. It should enable robust theory-based predictions and potentially lead to the development of disease-modifying treatments. Common symptoms that are documented as associated with LC include fatigue, low mood, shortness of breath, persistent cough, autonomic symptoms such as postural orthostatic intolerance, cognitive dysfunction, brain fog, sleep difficulties, low grade fever, and joint pain (212,226,252–254). Additional multiorgan manifestations as described in literature are myalgia, headache, chest pain or chest tightness, poor appetite, sicca, diarrhea, dizziness, sweating, alopecia, insomnia, restless legs, nightmares, and lucid dreams (252,255–257). An online survey conducted across multiple countries found that approximately 85% of respondents with persistent illness reported relapses, primarily due to exercise, physical or mental activity, and stress (252).

Besides chronic pain, other manifestations of fibromyalgia include easy bruising (27,258), urinary urgency (259), functional gastrointestinal disturbances, sleep disturbances, autonomic symptoms, wheezing, brain fog (27), a reportedly distinct brain pattern on functional magnetic resonance imaging (28,139,260), tingling, creeping or crawling sensations (259), reduced skin innervation (261), close association with gastroesophageal reflux disease (262), various autoantibodies among subgroups of patients (263,264), dry mouth, dry eyes, blurred vision, restless legs, multiple chemical sensitivity, fluid retention, and more (27,44,265). In a 2023 meta-analysis that included 188,751 patients, an increased standardized mortality (SMR) ratio in fibromyalgia was found for mortality from infections (SMR 1.66, 95% CI 1.15 to 2.38), accidents (SMR 1.95, 95% CI 0.97 to 3.92), and suicide (SMR 3.37, 95% CI 1.52 to 7.50) (266).

1. Diagnosis and assessment: bridging the gap to objective measures. This includes the need for biomarkers, the standardization of diagnostic criteria, phenotyping, correlation between objective findings and symptom severity, and addressing symptomatic overlap of the related syndromes.
2. Phenotyping: can help clarify the varied underlying biological mechanisms and facilitate the development of subtype-specific therapies.
3. Patient experience and coping. This includes, among other things, the consequences of clinician dismissal of symptoms (8), factors associated with late diagnosis, addressing the problem of medical stigma, and factors associated with over/under-diagnosis.
4. Treatment/rehabilitation and interventions: moving towards effective strategies for treatment, including treatments for addressing specific symptoms (neurological, fatigue, musculoskeletal pain, mood, etc.), non-pharmacological treatments, personalized medicine approaches, understanding mechanisms of treatments and how they relate to the pathophysiology, integration of digital therapeutics, and striving for more patient education. Future research into LC interventions shouldn’t neglect the role of a rehabilitative approach for treating LC and fibromyalgia.
5. Prevention: Need for better knowledge on evidence-based prevention strategies besides the obvious effort of avoiding infection.
6. Prognosis: Need for better knowledge regarding fibromyalgia and post-COVID fibromyalgia-type syndrome prognosis.
7. Methodological synchronization and harmonization in the field. As an evolving relatively amorphous field of research there is heterogeneity and inconsistency in fundamental aspects such as definitions, methods, and instruments used. Also, there are still limited systematic reviews, and there is a need for longitudinal studies.
8. Bridging basic science and clinical research.
9. Another significant knowledge gap concerns the extent to which clinicians are equipped with contemporary evidence-based knowledge regarding the evolving understanding of LC.

## 5. Recommendations for future research

LC research is an evolving field whose fundamentals are still being articulated. Future research should focus on epidemiological population-based studies with representative sampling and improving methodology, refining definitions, elucidating mechanisms in hypothesis driven studies, and developing effective therapeutic strategies. As some of the studies did not explicitly delineate their inclusion and exclusion criteria, and in studies incorporating a control group the criteria for control subject selection were not always clearly described, nor was there given a justification for the sample size taken, adhering to checklists such as STROBE and CONSORT recommendations for reporting observational studies and trials can add to more rigor in this new developing field.

Important methodological issues besides those mentioned above for consideration in the field based on the review are as follows:

- First, accurate assessment of post-COVID symptom trajectories necessitates future research that stratifies analyses by acute phase severity and hospitalization status. An analysis of all patients without making a distinction between severities of COVID-19 can add confounding factors related to hospitalization, antibiotic use, intensive care admission, and cases with well-defined organ damage, which could make it difficult to draw meaning from their results in terms of “LC.” The number of infection episodes, immunization status, behaviour and environmental factors during the initial recovery from the acute phase, and variant type, are also variables that could potentially be relevant to further investigate in the future. During the review process, patient surveys were found that did not corroborate the presence of the outcome being measured prior to acute covid-19, which makes it difficult to infer anything about new-onset or worsening of symptoms, or other self-reported measures.

Also, hypermobility syndrome appears to be another confounding factor that should be taken into account in future epidemiological studies of LC, as this seems to be an important variable for the phenomenon. It is important to emphasize that undiagnosed fibromyalgia and/or GJH may contribute to the development of LC but are frequently overlooked in the clinical setting. This can add confounding to studies that make use of official diagnostic codes and criteria for fibromyalgia. Due to the high cut-off set by the ACR criteria, the absence of fibromyalgia diagnosis, taken as an indicator for absence of fibromyalgia syndrome, may not suffice for choosing controls. For example, an individual with chronic widespread pain and somatic symptom severity score of 4 (that is, not eligible for official fibromyalgia diagnosis) in the control group could confound the results, as the cut-off chosen for fibromyalgia diagnosis by the ACR seems to be biologically arbitrary.

- Secondly, in studies using diagnostic criteria or diagnostic codes that distinguish functional psychosomatic syndromes, the investigator should recognize that making a distinction between chronic widespread pain and fibromyalgia diagnosis (and even ME/CFS), or ignoring their overlap, may confound results if the mechanism is shared, as has been suggested by some authors (43,64). Moreover, researchers conducting a correlation analysis between variables or outcome measures that represent overlapping constructs such as stress and fibromyalgia diagnosis or chronic widespread pain and fibromyalgia-type symptoms, or CSI score and depression (108,120,243,267), will end up with results that seem redundant unless that is what the study was designed to do.
- Third, it’s worth noting that studies that recognize central sensitization as a phenomenon simply based on hypersensitivity in the palmar side of the participants’ dominant hand, for example (112), do not necessarily relate to a mechanism of nociplastic generalized central pain augmentation and sensory hypersensitivity. If the authors of a study conclude that generalized central hypersensitivity and allodynia were found, then they might like to demonstrate that it is, indeed, both central and generalized. A methodological issue (29)was evident in literature in relation to the “central sensitization inventory” questionnaire, which tries to capture the impact of chronic pain conditions such as fibromyalgia (268) or “fibromyalgia-type features.” Authors have mistaken the CSI for central sensitization (111,115,117,139,184).

The following offers a mechanistic explanation for new-onset medically unexplained fibromyalgia-chronic-fatigue-and-somatoform-type manifestations after COVID-19, while attempting to reconcile the main findings from the systematic scoping review regarding fibromyalgia and LC, primarily including:

- A multifaceted etiology.
- Overlap between LC, chronic fatigue syndrome, related functional somatic syndromes and fibromyalgia symptomatology (including multiple medically unexplained multisystemic refractory symptoms, widespread myofascial discomfort and myofascial pain, hyperalgesia, itching, fatigue, post-exertional malaise, POT syndrome and autonomic symptoms, morning stiffness, spasms, irritable bowel, multiple chemical sensitivity, and more)
- Multisystem non-specific clinical findings (subclinical inflammation and immune dysregulation, metabolic abnormalities, low-grade hypoxia, muscle histopathological abnormalities, intraepidermal small fibre pathology, etc.)
- Unremarkable results on routine medical tests.
- The risk factors.
- Significant association with both hypermobility syndrome and low vitamin D.
- A relatively high prevalence of LC in mild and subclinical acute disease cases among previously healthy individuals.
- Insidious and heterogenous nature of the condition.
- Pain varying in anatomical location, and neuroanatomically illogical distributions.
- Other anomalies and counterinstances such as discrepancies between empirical findings and expected findings in nerve conduction studies and pressure pain threshold measurements, dissociation between measures of sensitization and subjective burden (249), low correlations between disease burden and conditioned pain modulation (269), autoantibodies and inconsistent findings regarding them (263,270), evidence of peripheral neuropathy in subgroups, disappointing and poor response to theory-based pharmacotherapies, symptomatic response to weather change (271), discordance between autonomic small fiber pathology and autonomic symptoms (272), and more (248–250).

There are many theories for fibromyalgia (e.g., autoimmunity, the gut-brain axis, nociplastic pain, etc.) (14,32–37,139,244,273–282), each of them leads to specific hypotheses, study designs, and study methods, and each can lead to different conclusions from the same findings. Any suggested theory should explain the broad symptomatology and manifestations of the syndrome besides merely pain and hyperalgesia.

## 6. Part two– synthesis of data and formulating a mechanism for “fibromyalgia syndrome” pathophysiology

Part 1 of this work presented findings from the scoping review which covered empirical evidence of new-onset fibromyalgia-type symptomatology after COVID-19. An in-depth review of the putative pathophysiology of LC was not the purpose of the review and can be found elsewhere (see section 3.2). Part 2 is a synthesis of data and conceptual analysis based on the scoping review, for reconciling the findings and anomalies from part 1. The following sections offer a conceptual framework for fibromyalgia pathogenesis, for the purpose of discussing this mechanism in the context of LC.

### 6.1. Fascial Armouring: a conceptual framework for the etiopathogenesis and cellular pathway of ‘primary fibromyalgia syndrome’

Fibromyalgia-type syndromes (or “functional somatic syndromes”, also called “chronic overlapping pain conditions” or “central sensitization syndromes”) and myofascial pain syndromes are suggested to be overlapping manifestations of a common medical entity with shared molecular pathways (74,139,265,280,283–289), or “two sides of the same coin” (283). The conceptual framework of ‘Fascial Armouring’ offers a non-autoimmune connective-tissue-based mechanism for fibromyalgia-type psychosomatic syndromes that’s based on the cascade of inflammatory myofibroblast force generation in soft tissue and dysregulated extracellular matrix remodeling, which may drive peripheral and central pain mechanisms (290). In its severe form, this suggested mechanism is anticipated to physiologically manifest as a mild-to-moderate global chronic exertional compartment-like syndrome (87,290), which might help explain “central sensitization symptoms” and propel fibromyalgia multiorgan and multisystem manifestations such as: pain, hyperalgesia, mechanical hypersensitivity, tender spots/trigger points, allodynia, general bodily discomfort, itching, muscle spasms, chronic fatigue, cognitive symptoms, autonomic abnormalities, cardiovascular and metabolic alterations, morning stiffness, small fiber pathology, intramuscular collagen organization abnormalities, metabolic abnormalities, various psychosomatic symptoms, overlap with other chronic psychosomatic-functional pain conditions, close association with hypermobility syndrome, various autoantibodies and close association with systemic autoimmune connective tissue diseases, atypical profile of inflammatory biomarkers, low efficacy of central neuroactive pharmacological agents (e.g., tricyclics and gabapentenoids), mostly silent routine medical investigations, signs of longstanding subclinical chronic ischemia and oxidative stress, and more (290). The term ‘psychosomatic syndromes’ within the context of this paper refers to disorders that are usually attributed to mental, emotional, or psychological disorders manifesting somatically in the body top-down (e.g., via neuroendocrine pathways) without tissue histopathological abnormalities, typically regarded in medicine as disorders of organ functionality, not organic diseases, i.e., disorders of function, not of tissue integrity, composition, architecture, or structure. In terms of nosology, what distinguishes primary fibromyalgia syndrome from other functional (psycho)somatic syndromes is simply a matter of clinical consensus of definition, a fashion, since the “diagnosis” is not biologically attached to a specific measurable mechanism. Thus, from the standpoint of molecular biology, what truly distinguishes between these clinical syndromes is still not entirely understood. Fibromyalgia is one of the psychosomatic syndromes.

The suggested mechanism is concisely outlined as follows, formulated by integrating five fundamental building blocks (this section presents the conceptual framework and theoretical model for the pathogenesis of fibromyalgia, and, afterwards, the clinical implications will be discussed):

#### (i) Normal mechanobiology of myofibroblasts

Myofibroblasts are contractile mechano-sensitive cells that can promote long-term contracture in tissue, and they have a complex mechanobiology often compared to smooth muscle cells. By synthesizing alpha-smooth muscle actin (α-SMA) fibres and focal adhesion complexes that grant them the ability to sustain mechanical tension in the surrounding extracellular matrix (ECM), myofibroblasts use a lockstep type mechanism to generate force (291). Myofibroblasts are induced by various signals and are part of the inflammatory and healing process, most known for in scar formation, but are also relevant, though somewhat overlooked, in other conditions including asthma (292), cardiac arrythmias (293), and during infection and events of systemic inflammation (294).

A positive feedback loop is established as myo/fibroblast cell contractility and ECM matrix remodeling stress-shields local mechano-active cells from external force while sustaining surrounding tissue contracture and is largely facilitated by transforming growth factor β1 (TGF-β1) (291). After TGF-β is secreted in a latent form and then activated through interaction with integrins, it binds TGF receptors on the cell membrane which in turn activates a signaling pathway and transcriptional elements that are responsible for α-SMA expression - one of the key players in this mechano-active festivity (291). Mechanical force that is generated by contractile myofibroblast cells expressing α-SMA, is mediated by, and also stimulates, integrins and focal adhesion complexes. This provides further input into the positive feedback of mechano-sensitivity, which leads to more α-SMA synthesis as well as ED-A fibronectin and allows for more force generation in a vicious cycle (291). A distinct cytokine-mediated pathway, the type-2 cytokine axis, may promote fibrosis independently of TGF-β, and involves the alarmin cytokines IL-25, IL-35, and IL-5 and IL-13 (295). Connective tissue growth factor (CTGF) is also a main actor in the signaling pathway of TGF-β-dependent myofibroblast stimulation (296). Nonetheless, clinically relevant factors that can attenuate or inhibit myofibroblasts are estrogen, vitamin D (via vitamin D receptor signaling), resveratrol, and more (297–300). Gut microbe-derived metabolites can influence fibroblast-to-myofibroblast differentiation and induce organ fibrosis (301). The delicate process of myofibroblast de-differentiation/senescence/apoptosis is important for the health of tissue and is influenced by factors such as fibroblast growth factors, prostaglandins, cellular communication network factor 1 (CCN1), metformin, and more (296,302).

Myofascial tissue of normal healthy individuals contains myofibroblasts that are likely to contribute to the development of pain and the manifestation of “myofascial pain syndrome” due to their natural biological activity (303,304). Empirical investigations have demonstrated that myofibroblasts are normally present in fascia and interstitial ECM and contribute to the pre-stress and basal tone of the tissue (303,305). Some authors suggest that abnormal mechanical tension in myofascial tissue can serve as a source of pain and myofascial trigger point-related nociception (306). Schleip and colleagues estimate that forces generated by soft tissue myofibroblasts may reach ∼2 Newtons and generate 1 cm per month of contracture that’s sustained by matrix remodeling, which is not at all negligible (303,307). Myofibroblasts are a phenotype of mechano-active smooth-muscle-like cell which generally have a similar behavior and mechanobiology irrespective of the anatomical location or the tissue (308).

Fibroblasts function as a large network (309). They form an extensive intricate cellular network in soft tissue that may have significant and underestimated physiological and functional importance (310). Fibroblasts can be arranged in nodules and cords and express altered contractile behavior and tensional homeostasis (311,312). Langevin et al. (2004) have shown, using confocal microscopy, histochemistry, immunohistochemistry, and electron microscopy, that fibroblasts form many cell processes and many points of cell-to-cell contact with each other when studied in vitro (310). About 30% of fibroblasts processes were shown to extend continuously from one cell to another using confocal microscopy. Other scholars have reported findings consistent with this when investigating human fibroblasts in vivo (313,314). When fibroblasts experience mechanical stimuli, they initiate a range of cellular responses such as changes in intracellular calcium and adenosine triphosphate release, activation of intracellular signaling, actin polymerization, and gene expression. It is possible that oscillations of calcium waves are a main facilitator of intercellular communication of fibroblasts, through fluctuations in the levels of cytosolic calcium and its effect on downstream cell signaling pathways (310). The nature of these oscillations likely depends, among several different factors, on substrate rigidity (315).

#### (ii) Tensegrity qualities when superimposed on the interconnectedness of the fascio-musculo-skeletal system

Fascia and the extracellular matrix constitute a complex dynamic interconnected extensive fiber-cellular network of connective tissue that undergoes a process of continuous remodeling and transmits and absorbs loads while it exhibits tensegrity-type qualities (290,304,316). ‘Tensegrity’ (the words ‘tension’ and ‘integrity’ merged) is a concept that describes the homeostasis of a complex pre-stressed structure that is stabilized under forces of compression and tension and functions as one connected spatial system (316–318). ‘Bio-tensegrity’ (319) is a biophysical conceptual framework under continuum biomechanics that incorporates the principles of tensegrity for a better understanding of human physiology and kinematics (304,320,321). It is a theoretical concept of biomechanics integrated into our discussion as a simplification. The theory suggested here also stands without the idea of organismal “bio-tensegrity,” though, as with most models, its aim is to simplify. In living tissues there is an ongoing dynamic balance of forces of cell traction and points of resistance within the ECM, with a state of reciprocal isometric mechanical tension (316). The dynamic bio-tensegral system and mechano-transducing signaling enable cells to mechanically sense changes, modify their microenvironment, and promote ECM remodeling in homeostasis and in disease states (316). Figure 3A-D displays tensegrity structures as an illustration of this concept of a pre-stressed structural system in a steady-state that is maintained in a balanced equilibrium of compressive and tensional forces – as an allegory for the human body. Its aim is to illustrate an anatomical situation of mechanical imbalance in the (fascio)musculoskeletal system. The purpose is to demonstrate tensegrity as a pillar in the model, not a specific clinical syndrome.

**Figure 3.**
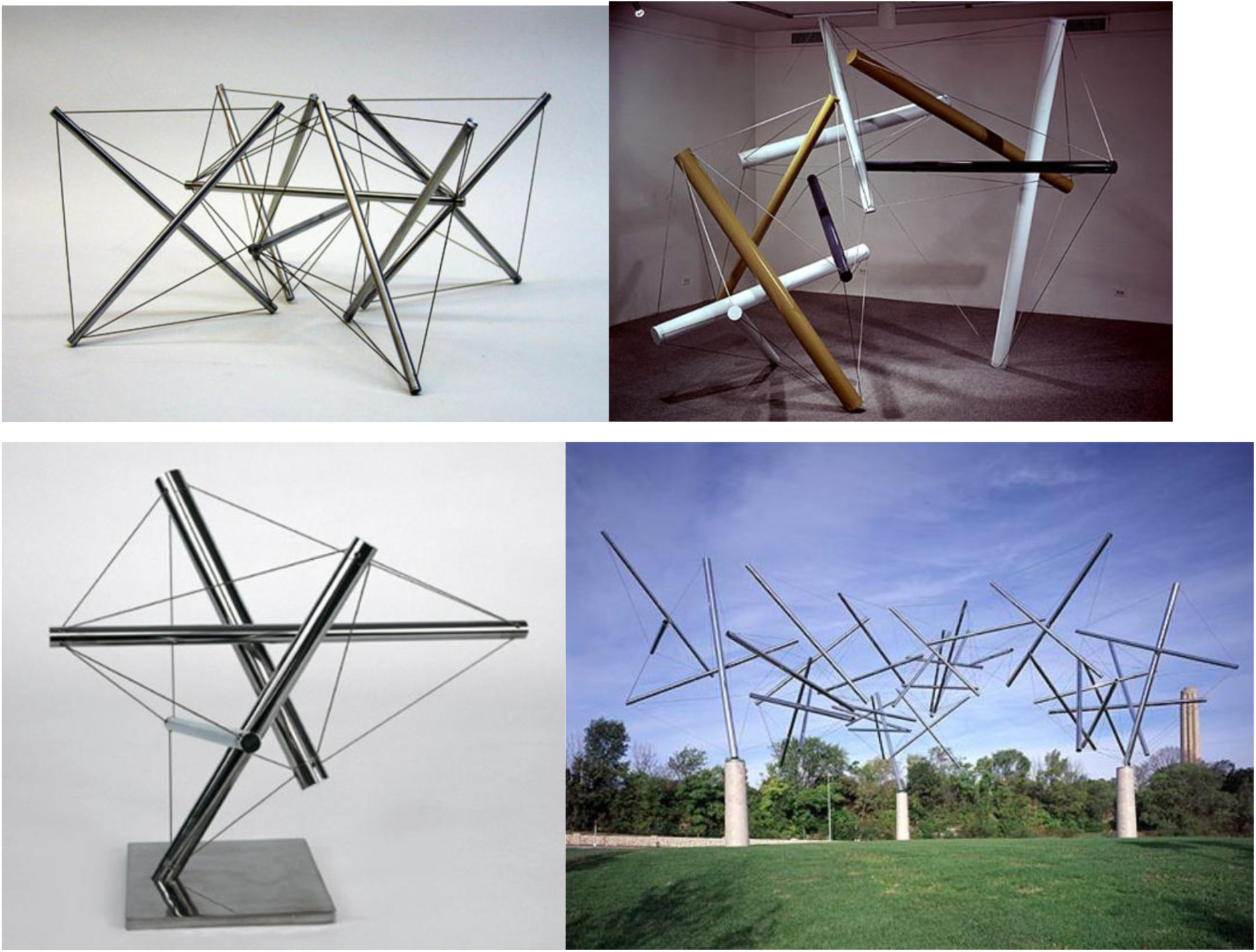
An illustration of the concept of tensegrity. Floating compression elements transmit force through the tension elements. Changes to one node affect mechanical homeostasis of the structure and other nodes as well. In the setting of continuum biomechanics this principle can be called ‘bio-tensegrity’. Kenneth Snelson’s Audrey I 1966 (top right), eight up 1967 (top left), triple crown 1991 kansas city, MO (bottom left). 60.5 degrees 1992 stainless steel tensegrity structure (bottom right). Images from http://kennethsnelson.net.

Observational studies highlighted the relevance of bio-tensegrity mechanotransduction on tumor cells by mediating the cellular response to ECM stiffness (316,319). In addition, existing empirical investigations of ECM (and fascia) in humans in vivo support the tensegrity properties of fascia by demonstrating its role in a continuous myofascial system where tension is balanced across different segments. For example, studies have shown that sustained manual pressure on the lateral raphe in patients with chronic low back pain resulted in an anterior shift of the transversus abdominis musculofascial corset system, suggesting the release of pre-existing tightness or adhesion in the posterior fascia and a change in its elastic properties (322). Manual intervention has also been shown to lead to increased sliding and thickness changes of the transversus abdominis, indicating a redistribution of tension within the myofascial system. Furthermore, research on isometric plantar-flexion demonstrated a strong correlation in stiffness changes between the lower limb muscles (gastrocnemius) and lumbar tissues (thoracolumbar fascia and erector spinae), highlighting a long-distance interaction within the myofascial tensegrity network (323). These findings collectively reinforce the concept of fascia as a force transduction network rather than merely local passive structures, supporting its tensegrity role in maintaining body stability and function. Virtually all organs and tissues are organized as prestressed structural hierarchies that exhibit immediate mechanical responsiveness and increase their stiffness in direct proportion to the applied mechanical stress (324). Molecules, cells, tissues, organs, and our entire bodies use “tensegrity” architecture to mechanically stabilize their shape, and to harmonize structure and function at all size scales (325,326).

Like any other model, the tensegrity model described here is a simplification of the theory. Because tensegrities are composed of discrete networks of support elements, rather than a uniform medium like a chunk of metal or a rubber band, they provide a way to transmit mechanical forces along specific paths and to focus or concentrate stresses on distant sites and at different size scales. These are all features observed at the level of whole organs as well as tissues, cells, membranes, cytoskeletal networks, subcellular organelles, nuclei, mitotic spindles, transport vesicles, viruses, and proteins (325).

#### (iii) Myofascial chains

Fascia constitutes a most ubiquitous tissue that permeates the human body and is capable of transmitting and dispersing mechanical forces to a distance because of the structural connectivity of the (fascio)musculoskeletal system (304,327,328). On a more macroscopic level, as part of normal physiology, internal mechanical forces are transmitted within myofascial tissue along mechanical links called myofascial chains (304,307,321,327,329). In this way, for instance, force in the lower limb can be transmitted to the trunk and affect the lumbar musculature (327). Stretching of the upper limbs can lead to an increased maximal range of motion in the lower limbs, and vice versa (327,330). Cadaveric studies investigating force transfer in the human body indicate that anatomic structures normally described as leg, hip, and pelvis muscles interact with muscles of the spine and arm through the thoracolumbar fascia, thus forming an integrated functional system that allows for load transfer between the spine, pelvis, legs and arms (331). For example, the posterior layer of the thoracolumbar fascia was found to be continuous with the fascia of the gluteus maximus, and some of the superficial lamina fibers were found to cross the midline and fuse with both the lateral raphe and fibers derived from the fascia of the latissimus dorsi (331). Muscle and fascial tissue do not exist in isolation, but rather they function together in synergy to facilitate the body’s movements through mutual connections thus forming a myofascial tensional network that connects all parts of the body as a whole (323). Most skeletal muscles in humans are connected through connective tissue (327).

#### (iv) Innervation and sensory functions of fascia

Fascia contains a densely interwoven network of sensory nerve endings that are involved in the perception of pain (304,332–334), although the relationship between the nervous system and fascia is a relatively neglected field of research. Free and encapsulated nerve endings are located within myofascial tissue (335), including interoceptive receptors and Ruffini and Pacini corpuscles (304). Superficial fascial tissue is associated with skin mechanoreceptors and thermoreceptors, while the deep fasciae are known to affect proprioception (305). Fede & Stecco et al. (2021) showed that in fascia an impressive network of sympathetic nerve fibers is found, as was demonstrated in samples from mice (336). Nociceptor free nerve endings terminate in muscle interstitium. Non-myelinated C fibre receptors in muscle tissue are polymodal and respond to high mechanical pressure and chemical stimuli (337–339), as do A-delta fibres which are related to stretch receptors (337). The biochemical milieu can therefore affect nociception when nociceptive substances accumulate in muscle interstitium (340).

Fascial dysfunction, overuse, strain injury, trauma, and inflammatory changes, are postulated to lead to pain due to pathological ECM remodeling accompanied by chemical and mechanical alterations (341). Pathological changes in fascia are characterized by increased tissue stiffness and changes in the ECM, including changes in both collagens and matrix metalloproteinases levels as well as alteration in myofibroblast activity (332). Abnormal mechanical forces and nociceptive mediators that are secreted by myofibroblasts and local cells (e.g., interleukin 1-beta, tumor necrosis factor-alpha, neuropeptide Y, substance P) may trigger pain via activation of peripheral sensory receptors (304). The transient receptor potential ankyrin cation channel TRPA1, which is widely expressed in sensory neurons, is known to respond to mechanical and chemical stimuli and is involved in acute and chronic pain, as well as in the sensation of itching (pruritus) (342). Among its natural endogenous agonists are products of oxidative stress. Lack of TRPA1 may attenuate the expression of transforming growth factor beta 1, interleukin 6, and α-SMA (342). Transient Receptor Potential vanilloid 4 ion channel TRPV4 is known to be responsive to mechanical stimuli and is likely to be relevant in musculoskeletal pain (343,344).

#### (v) Substrate stiffness & rigidity of ECM

ECM stiffness seems to be a crucial factor in the behavior and function of nerve cells (345). Researchers have investigated the effect of matrix rigidity on neuronal cells in vitro, and found a marked difference in growth dynamics, synaptic density and electrophysiological activity of cortical neuronal networks when comparing cultures grown in substrates with 100-fold differences of young modulus (346). Matrix stiffness may be a significant factor to modulate Schwann cell function and behavior (347). Specialized Schwann cells form a mesh-like network in the subepidermal border of the skin and are intimately associated with unmyelinated nociceptive nerves. This cell type is inherently mechanosensitive and capable of conveying nociceptive information to the nerve. As was shown using transmission electron microscopy, a distinct thick layer of fibrillar collagen is found to envelope their cell processes (348).

The above findings, integrated mechanistically, provide the five main elements for forming the theoretical model of Fascial Armouring. Essentially, it can be summarized as: myofibroblast-mediated bio-tensegrity tension, compression, and ECM stiffness on a background of interrelated myofascial tissue and myofascial chains. Basically, it is a myofibroblast-driven disease of the fasciomusculoskeletal system, whose severe manifestations would be comparable to a mild-to-moderate global chronic exertional compartment-like syndrome (or a “myofascial pain syndrome of the whole body”) (290).

Empirical findings that, when taken together, may support this mechanism for fibromyalgia can be found in several studies and are listed in Table 8 below. A more elaborate analysis of this model in relation to fibromyalgia and LC can be found in a recent study (87,290). A myofascial-based mechanism might help explain several manifestations of LC and fibromyalgia (and the overlap of both), particularly when initial infection was mild or asymptomatic and when medical evaluation reveals no prominent organic abnormalities such as pulmonary fibrosis or other organ damage.

**Table 8:** Summarizes empirical findings that may support a tensegrity connective tissue-based mechanism for fibromyalgia.

| # | Abnormality | Findings |
| --- | --- | --- |
| 1 | Increased intramuscular pressure | High intramuscular pressures in trapezius muscle of fibromyalgia patients were documented in a study by Katz et al. (2021). The patient group had a mean pressure value of 33.48 mmHg compared to 12.23 mmHg in controls (32). The authors of that study assert that such muscular pressure abnormality might be a critical finding that contributes to diffuse muscle pain in fibromyalgia and may be an intrinsic feature of the disease. They conclude that the idea of fibromyalgia as a disorder driven exclusively by central mechanisms without any peripheral organic causes should be reevaluated (32). This musculoskeletal finding is interesting because in common practice (349) chronic exertional compartment syndrome usually exhibits silent routine medical investigations and is diagnosed if at least one of the following intramuscular compartment pressure measurements is made: (a) pre-exercise intramuscular pressure of more than 15 mmHg (b) one minute post-exercise pressure of more than 30 mmHg, or (c) five-min post-exercise pressure of greater or equal |
|  |  | to 20 mmHg. Studies measuring intramuscular pressure of other muscles besides trapezius in fibromyalgia were not found |
| 2 | Muscle tension and myofascial taut bands | Significantly higher measurements of muscle damping were found in fibromyalgia patients which reflects increased muscle tension (350). Myofascial taut bands (i.e., palpable hardened and contracted muscle fibers) are very common in fibromyalgia patients (351). According to a patient survey, morning stiffness was ranked as being bothersome to a similar degree as pain in individuals suffering from fibromyalgia (271). A 2022 observational study from Spain found a weak but significant association between elastic properties of soft tissue measured by ultrasound elastography at the tender point areas, and psychological factors (Pain Catastrophizing Scale scores) in 42 female patients with fibromyalgia (352). Zetterman and colleagues (2021) report that 51 female fibromyalgia patients in their study had significantly higher mean %EMG and shorter EMG rest time values than the controls (247). Fibromyalgia disease burden, as evaluated via patient self-reported questionnaire (CSI), is significantly correlated ($r=0.546$) with muscle stiffness in patients rating pain 4 or higher on a numeric rating scale (353). |
| 3 | Imaging | A study by Müller et al. (250) found no evidence for functional or structural alterations in brain areas typically involved in acute pain processing that could reflect chronic stimulus-independent pain in FM patients. |
| 4 | Trigger points | The local pain of chronic widespread pain and fibromyalgia patients is often related to the presence of myofascial trigger points (354). For the most part, myofascial trigger points make up the topography of fibromyalgia tender spots ($r = 0.78$, $p$-value $<0.001$) (355). It is recognized that in a substantial amount of cases symptoms attributed to “central sensitization” are |
|  |  | actually maintained by bottom-up sensory signals originating in peripheral tissue such as soft tissue and the musculoskeletal system (275). |
| 5 | Fascial/epimysium stiffness | Kawakita and colleagues from Japan in their 1991 observational study (356) investigated deep pain measurements in fibromyalgia tender spots by using needle electrodes. They describe that the needle was inserted to a depth where the experimenter felt some physical resistance or stiffness against the needle insertion (what they have termed “needling stiffness”). The minimum pain thresholds under the tender point occurred at the depth of the needling stiffness. Observations via ultrasound scan suggested that the needle tip at this depth was located on or near the fascia. |
| 6 | Chronic ischemia (357) and muscle histopathological findings | Histological analysis of fibromyalgia patient biopsies reveals nonspecific muscle changes, such as segmental fiber necrosis, accumulation of lipids and glycogen, and subsarcolemmal mitochondrial clustering, which were hypothesized to stem from sustained muscle contraction and ischemia of unknown etiology (358). In muscles of fibromyalgia patients, chronic contraction appears to contribute to increased DNA fragmentation and structural changes within the muscle tissue (359). Hénriksson et al. (1982) investigated biopsy specimens from muscle of fibromyalgia patients. They described finding moth-eaten fibres that were evenly distributed over the whole cross-section and only type I fibres were affected. Electronmicroscopy of specimens revealed mitochondrial abnormalities (such as electron-dense inclusions and lack of inner membrane), myofibrillar Z-streaming, and cytoplasmic bodies, and glycogen concentrations were below normal. There was an abnormal relation between mitochondria and myofibrils. The authors |
|  |  | stated that the biopsy findings described may be of diagnostic importance (360). |
| 7 | Serum cytokines, elevated levels of fibroblast TGF-β expression and ECM abnormalities | A 2020 gene expression analysis study of peripheral blood samples from ten fibromyalgia patients identified a transcriptional profile related to the immune system (361). It also found significantly higher levels of two long non-coding RNAs and higher serum levels of IL-17, TGF-β, IL-6, IL-21, and IL-23 in the fibromyalgia group using ELISA method. The results of the study were validated on a group of 50 fibromyalgia patients (48 females, mean age: $49 \pm 20.5$ years) (361). A biopsy study of fibromyalgia patients indicated a significant increase in TGF-β gene expression in peripheral fibroblasts compared to controls (362). The expression of proteins associated with ECM remodeling and oxidative processes appears to differ in fibromyalgia fibroblasts, potentially explaining a heightened inflammatory state (363). Findings from a 2002 study indicate significantly elevated levels of interleukin 1-beta, interleukin 6, and tumor necrosis factor alpha in skin of a subset of fibromyalgia patients, detected in biopsy samples taken from the deltoid region of female patients, by using RT-PCR and immunohistochemistry (364). Immunoglobulin G deposits in dermis were also previously demonstrated and could be related to the patho-mechanism or merely an epiphenomenon (364). The observed decrease in intramuscular collagen levels could predispose fibromyalgia patients to muscle micro-injuries and result in non-specific signs of muscle pathology (365). |
| 8 | Metabolic alterations | The interstitial levels of metabolic substances in individuals with fibromyalgia were evaluated through microdialysis (366). Concentrations of glutamate, lactate, and pyruvate were found to be significantly elevated. The response to acetylcholine |
|  |  | stimulation was shown to be associated with increased dialysate lactate in fibromyalgia (367).<br>Increased pyruvate levels and lower adenosine triphosphate and creatine phosphate are seen in muscle interstitium in fibromyalgia patients (368). More studies have revealed findings in line with this result (340,369). |
| 9 | Oxidative stress | An abnormality of mitochondrial function is suspected to occur in fibromyalgia (368), and may help explain the pain of the syndrome. A transmission electron microscopy study revealed morphological alterations in mitochondria of peripheral blood mononuclear cells from fibromyalgia patients. This finding is suggestive of mitochondrial dysfunction and consequential inefficient oxidative phosphorylation, metabolic and redox disorders, and increased reactive oxygen species levels, which may play a pathogenetic role in fibromyalgia (370).<br>Also, fibromyalgia is characterized by a hypomethylated DNA profile with an overrepresentation of genes linked to stress response and DNA repair/free radical clearance (371). Cellular energy metabolism abnormalities seem to promote fibromyalgia and possibly other chronic pain conditions, indicative of a role of oxidative phosphorylation in pathophysiology of chronic pain (372). Oxidative stress seems to have an important role in fibromyalgia's clinical picture (373,374). A clinical study demonstrated that hyperbaric oxygen therapy significantly alleviates fibromyalgia symptoms and improves patient reported quality of life significantly (375). |
| 10 | Cardiovascular system | Evidence of decreased peripheral blood flow in individuals with fibromyalgia suggests underlying functional impairments in their cardiovascular system (376). There is evidence indicative of decreased transcapillary permeability in fibromyalgia |
|  |  | patients (377) as well as abnormalities in peripheral arteries (378). |
| 11 | Differential modulation of disease course by invasive surgery | Surgery deeply affects fibromyalgia manifestations in a differential manner: whereas some surgeries improve the course of the disease, other operations are associated with triggering or worsening fibromyalgia (379–382). For example, while fibromyalgia is a risk factor for less satisfaction, more postoperative pain and opioid prescription, worse functional outcome, and higher rate of medical and surgical complications following orthopaedic surgery (383), a 2008 study found that laparoscopic Roux-en-Y surgery is associated with resolution or improvement of fibromyalgia (384). A 2014 study found that following parathyroidectomy, fibromyalgia medication use decreases remarkably, and quality of life is significantly improved (385). In hip and knee arthroplasty, fibromyalgia patients that had pre-operative lower body symptoms experienced general improvement, while patients with upper body pain reported a worsening of symptoms, when studied at one and six-months postoperatively (275). Irritable bowel syndrome (IBS) (286), a functional gastrointestinal disorder which has much clinical overlap with fibromyalgia (55), was shown to be relieved to below Rome II criteria in 80 percent of patients after laparoscopic anti-reflux surgery (380). Meanwhile, fibromyalgia patients have a higher incidence of IBS after appendectomies (386). Laparoscopic cholecystectomy has been shown to deeply influence fibromyalgia symptoms (379), and hysterectomy, with or without oophorectomy, seems to worsen fibromyalgia (387,388). |
| 12 | Correlations between disease burden and other variables | A systematic review and meta-analysis investigated the association between the CSI questionnaire scores (evaluating fibromyalgia-type disease burden) and quantitative sensory testing (QST) in different chronic pain populations and found |
| | misalign with theory-based expectations | that for conditioned pain modulation, the best correlation found with the CSI score was $r=-0.13$. Temporal summation, pressure pain threshold, heat pain threshold, and cold pain threshold, showed correlation coefficients of $r=0.16$, $r=-0.36$, $r=-0.17$, and $r=-0.27$, respectively. |
| 13 | Other | Studies suggest that fibromyalgia patients have higher prevalence of carpal tunnel syndrome (389), tension type headache (27), gastroesophageal reflux disease (262), and reduced optic disc perfusion (390). Additional anomalies and counterinstances are also found (249). |

### 6.2. “Fascial armoring” as a fascio-musculoskeletal medical entity of continuum biomechanics

Let us now analyze this theoretical mechanism as a medical entity with clinical reasoning, for instance: one may infer from the mechanism that if myofibroblast-mediated “bio-tensegrity” tension and fascial stiffness develops in the temporal fascia, it is expected to manifest as a tension-like “primary headache disorder” (87). If tensegrity imbalance and compression transfer to thoracolumbar fascia, the expected manifestation would be a chronic “none-specific” low back pain (290). If it affects the palms and hands, it is expected to predispose to a Raynaud-like phenomena and/or carpal tunnel syndrome. Tension transferred to the spinal denticulate ligaments or dura-spinal-cord-associated abnormalities. Prevertebral fascia-spinal stiffness. If in the pelvic fascia-urinary urgency. Chest and torso-chest tightness, shallow breathing, and if severe, non-cardiac chest pain. Diaphragm and abdomen-predisposition to gastroesophageal reflux. Neck and pretracheal fascia-muscle tension dysphonia and dysphagia. Cervical fascia, parotid fascia, and superior cervical ganglion-dry mouth. Baroreceptors and stellate ganglion-autonomic imbalance. Neurovascular bundle and perivascular nerve plexus or nervi nervorum-sensory and vascular irregularities. Jaw-temporomandibular tension, dysfunction, and pain. Tympanic membrane and inner ear-hearing abnormalities. Proprioceptors-impaired coordination, impaired balance, microsomatognosia, and new onset clumsiness. Joint capsule or tendons-decreased joint range of motion, and if myotatic reflex is involved-muscle spasms. Muscle spindles-increased resting tone and activation of the stretch reflex, and sustained tonic muscles during sleep. Epimysium-compression of striated muscle. Increased rigidity of subcutaneous fascia - activation of TRPA1 channels and itching sensation, and substrate stiffness dependent neurite alterations. Celiac plexus, mesentery, visceral fascia, abdominal muscles and/or the gut wall-disrupted peristalsis, subsequent bloating, distension, and stretching of the gut wall, alterations in gut microbiota, and so forth. Abnormal pendulousness affects gait and kinesthetics (e.g., robotic gait). Altered absorption and dissipation of kinetic energy throughout the fascio-musculoskeletal system would lead to a subclinical “functional” movement impairment without classic neurological signs (e.g., upper or lower motoneuron pathology or cerebellar dysfunction). Increased intra-abdominal pressure has the potential to compromise blood flow in the gonadal arteries/veins. Sleep impairment is a non-specific complaint easy to disregard as a psychological issue. Morning stiffness, a general feeling of heaviness, hypervigilance, mild pallor, low mood, and constant exhaustion, are naturally deduced if the abnormality is widespread. Recall that the guiding theme when clinically interpretating this entity is: myofascial tension, compression, and high ECM substrate stiffness. Worth noting, the mechanobiological cascade of myofibroblasts is not necessarily propagated as a classic inflammatory leukocyte-driven disease in its nature, therefore, it can be mostly silent when examined in routine medical investigations. Systemic subclinical chronic oxidative stress is achieved by this theoretical model as it portrays a global chronic exertional compartment-like syndrome. Unmyelinated muscle nociceptors are activated by hypoxia of muscle tissue which is exacerbated by muscle contraction (354,391).

Sugawara et al. (1996) (392) report that mechanical compression of the dorsal root ganglion by a stimulus decreases the threshold needed to trigger a neuronal response, leading to the generation of action potentials. The same (in vitro) study also suggests that these action potentials can continue after the stimulus is removed, indicating increased mechanical sensitivity (392). Results from another (in vivo) study seem to be in line with these findings, revealing an ectopic spontaneous discharge generated within chronically compressed ganglia (393). It is interesting to note that dysfunction of the thoracolumbar fascia has been described as a chronic compartment syndrome of the paraspinal muscles (394).

Some of the variability of this mechanism relates to fibroblasts being a diverse cell family. They secrete cytokines, growth factors, and various inflammatory mediators (295), neurotrophins (395), and matrix metalloproteinases, and can uptake cellular signaling molecules and serotonin which affect molecular biological pathways and metabolism (291,295,396). In certain conditions myo/fibroblasts can express major histocompatibility complex class II and CD74 and stimulate CD4+ T-cells in an antigen-dependent manner via T-cell receptor ligation (397). Also, substrate stiffness affects the function of monocytes/macrophages, dendritic cells, B-cells, and other immune cells (398). Low-grade inflammation is implicated in this mechanism. To the best of the author’s knowledge, no study has examined fascial myofibroblast concentration or blood CTGF levels in fibromyalgia. Box 1 summarizes T-cell dysregulation and its relevance to this framework.

#### Box 1: T helper 17 cells and T regulatory cells and their relevance to the pathophysiological model

T helper 17 (TH17) and regulatory T cells (Treg) are two distinct CD4+ T cell subtypes with opposing functions in the immune system. TH17 cells, regarded as proinflammatory, produce IL-21 and IL-22, as well as signature cytokines IL-17A and IL-17F, and are implicated in the pathogenesis of various autoimmune and inflammatory diseases (399). TH17 cells not only trigger B-cell proliferation but also promote the formation of germinal centers together with isotype switching (399). In contrast, Treg cells are known for their immunosuppressive properties, mediated by the expression of FoxP3 and the production of TGF-β, and play a crucial role in maintaining immune homeostasis and preventing autoimmunity (400). The differentiation of naive CD4+ T cells into TH17 or Treg cells is critically influenced by the cytokine milieu present during T cell activation. In addition to these effector T-cell subsets, a specialized T helper cell subset, called follicular B helper T cells has been identified, which plays an important role in B cell induction of induction of germinal centers and isotype class switching.

The development of TH17 cells is driven by a combination of pro-inflammatory cytokines, including IL-1β, IL-6, and TGF-β. These cytokines activate the transcription factor RORγt, which is essential for TH17 cell differentiation and the production of IL-17 family cytokines (400). IL-23, while not required for the initial differentiation, is crucial for the survival and expansion of TH17 cells (400). Treg cell differentiation is primarily induced by TGF-β and mediated by the transcription factor FoxP3 (400). A delicate balance between TH17 and Treg cells is tightly regulated by the interplay of cytokines. While TGF-β together with the inflammatory cytokine IL-6 can induce the differentiation of naive T cells into the Th17 phenotype, TGF-β, favors Treg induction (400). The dysregulation of the Th17/Treg balance has been implicated in various autoimmune and inflammatory diseases (401).

Interestingly, TH17 cells, known for their pro-inflammatory cytokine production, can promote myofibroblast activation and collagen deposition, contributing to fibrosis (402). IL-17A increases and stabilizes TGF-βRII expression on fibroblast, and the TH17-associated cytokine IL-22 similarly enhances TGF-β signaling in fibroblasts (295). It was shown that TGF-β in turn induces the expression of IL-17A when produced concurrently with the pro-inflammatory cytokines IL-1, IL-6, or TNF (295). Some of the abovementioned cytokines known to be produced and secreted by myofibroblasts (e.g., IL-6) (401), can influence TH17/Treg balance (399,400).

Since LC is a newly recognized syndrome, empirical studies have yet to fully investigate whether similar abnormalities occur in post-acute cases of sars-cov-2 infection. Myofascial tissue in LC is a relatively neglected field of research. When searching MEDLINE for the term “COVID myofascial” only 20 items were found, mostly in the field of physiotherapy.

Figure 4 outlines fascial armouring as a medical entity of rheumato-psycho-neurology for explaining fibromyalgia-type syndromes. The main motif to keep in mind is substrate rigidity of ECM, myofascial tension, mechanical compression, and tensegrity imbalance. The clinical presentation would depend on multiple factors and does not necessarily depend strictly on the occurrence of pain. Pain in this framework is a manifestation of the entity, but it isn’t the actual entity itself. The suggested theoretical model, intrinsically, has variations.

**Figure 4.**
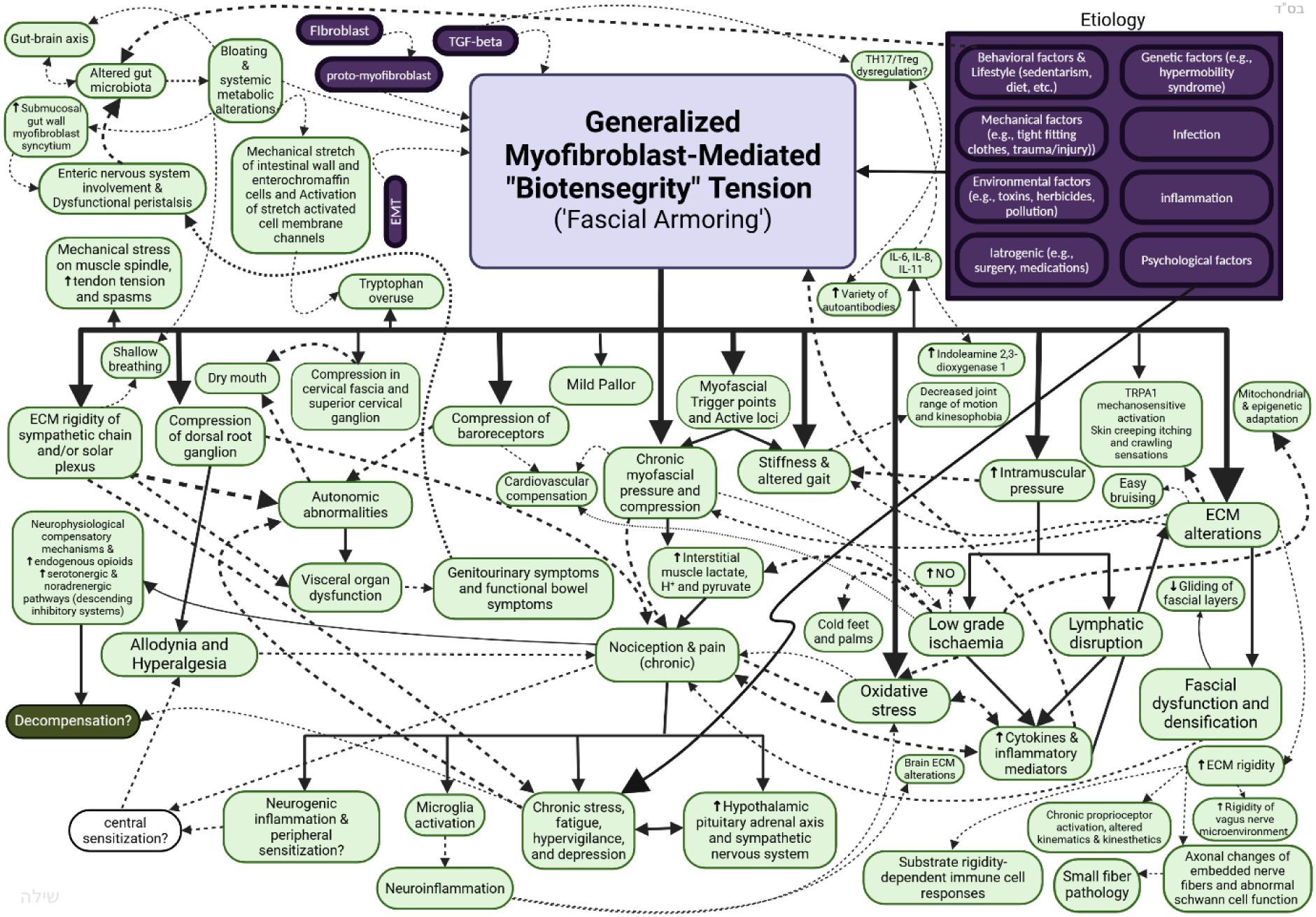
An outline of Fascial Armouring as a multifactorial rheumopsychoneurological medical entity. The clinical manifestation de facto depends on several factors (environmental, genetic and epigenetic, behavioral, psychological, co-morbidities, the specific cytokine profile, etc.) as well as the severity and location of the armouring and anatomical structures involved, and compensatory mechanisms. Involvement of the abdominal fascia, enteric nervous system, psoas muscle, and intestines navigates the abnormality more towards an irritable bowel-like syndrome. Involvement of the pelvic fascia navigates more towards genitourinary symptoms. Tensegrity forces affecting the lower back area will manifest as a tension-like “non-specific” low back pain, and so forth. A widespread disorder would manifest initially as multiorgan medically unexplained symptoms or a “somatic symptom-like disorder” (later in severe cases- “fibromyalgia syndrome” or a “myofascial pain syndrome of the whole body”). Some susceptible individuals might be more prone to neurophysiological dysfunction and mood disorders in chronic cases. The boundary of a medical entity goes as far as the boundary of the biological process which underlies it. An unhealthy lifestyle is an important contributing factor to this entity, as seen in the top right rectangle representing the etiology. Sedentary individuals would have a larger component contributed by immobilization stress especially during lockdowns. Pain leads to the release of neuropeptides, cytokines, and chemokines, and neurogenic inflammation. It is accepted by neurobiology that peripheral sensitization may lead to central sensitization, a process mediated by neurotransmitters such as glutamate. Many factors lead to sleep disruption. The gut-brain axis is likely involved as well. Persistent cough, as often seen following COVID-19, would likely be related to pulmonary injury during the acute phase. Olfactory/gustatory disfunction would be related to injury of olfactory/gustatory system during COVID-19. Pancreatic injury could lead to metabolic abnormalities, and so forth. This scheme focuses on the psychosomatic subtype of long COVID-19, where no well-defined abnormality is found following a mild or asymptomatic infection, and symptoms and complaints seem out of proportion and don’t make much sense clinically. Any mechanism suggested for this entity must be able to explain its broad symptomatology besides merely pain. Dashed lines and arrows have no unique significance in this schematic and are simply meant for visual clarity. Not all links and relationships are depicted in this scheme. ECM-extracellular matrix, IBS-irritable bowel syndrome, MMP-matrix metalloproteinase, POTS-postural orthostatic tachycardia syndrome.

### 6.3. Soft tissue myofibroblasts in the context of Covid-19

Figure 5 outlines the positive feedback loop of myofibroblast force generation in ECM alongside various factors that may provide enhancing or suppressing input to regulate the pathway. For simplicity, latent TGF-β and the proto-myofibroblast phenotype are not shown. Worth note, lifestyle is one of several etiological factors in this framework. A cytokine storm that may occur during infectious diseases is expected to fuel this pathway, leaving behind a remodelled and less healthy fascia. During infection with SARS-CoV-2, as with other infections, pro-inflammatory and pro-fibrotic processes are activated and involve various inflammatory mediators including TGF-β (403). TGF-β was mentioned as a main cytokine that fosters the differentiation and cellular activity of myofibroblasts (404). After SARS-CoV-2 infection, fibrotic changes facilitated by myo/fibroblast are seen in several tissues and organs (including lungs, heart, kidney, liver, intestines, and more) (254,405–414). Muscle biopsies in post-COVID-19 patients with persistent complaints of fatigue, myalgia, and/or weakness lasting for up to 14 months revealed myopathic changes, including muscle fiber atrophy, mitochondrial abnormalities, subsarcolemmal accumulation, inflammation, and capillary alterations, suggesting skeletal muscle as a target of SARS-CoV-2 (178,179). The angiotensin converting enzyme 2 (ACE2) and TMPRSS2, which are the key mediators that allow viral invasion by SARS-CoV-2 (415), are found in extrapulmonary and musculoskeletal tissue including muscle cells, smooth muscle cells, pericytes, endothelial cells, macrophages, chondrocytes, synovium cells, osteoblasts, and osteoclasts (416–423). Mast cell-mediated activation of fibroblasts can contribute to fibrotic changes seen in LC (11).

**Figure 5.**
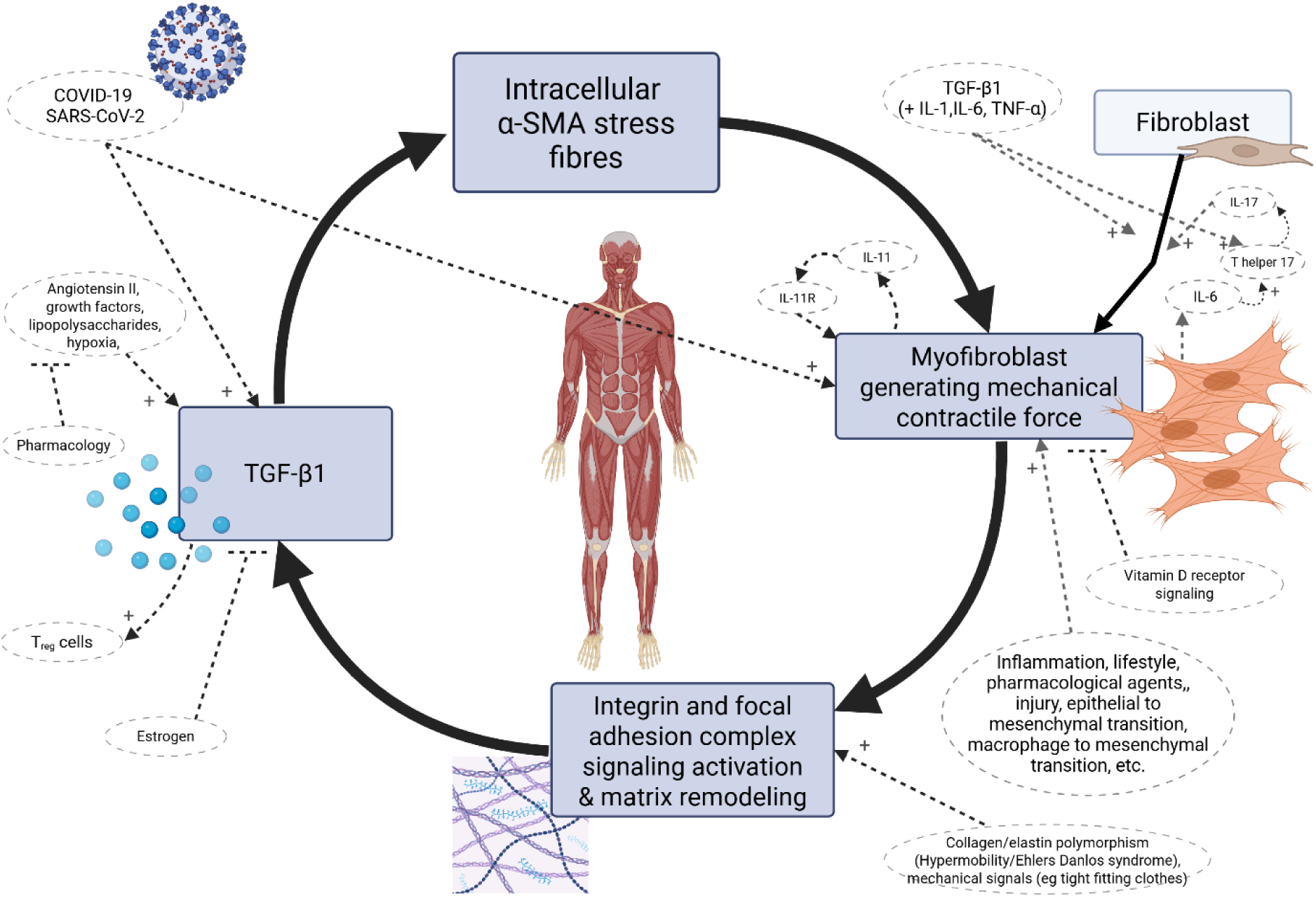
The vicious cycle of myofibroblasts transforms the osteomyofascial tensegrity-like system into a high pre-stress biomechanical system: This is suggested to be the core cellular pathway of primary fibromyalgia syndrome. Various factors provide input into the cycle of myofibroblast contractility and matrix remodeling. Insofar as COVID-19 involves systemic cytokines, pro-inflammatory, and pro-fibrotic signals including TGF-β1, and stimulates fibroblast-to-myofibroblast differentiation in various tissues, it is part of the etiology. Studies have shown that estrogen inhibits TGF-β1 and myofibroblasts and is associated with lower fascial stiffness (404,424,425), therefore, estrogen is expected to be a protective factor whereas low estrogen states are anticipated to be a risk factor (87). Myofibroblasts secrete cytokines such as IL-11, IL-8, and IL-6 (295,401), some of which are crucial to the balance of TH17 Treg cells (400,402). IL-17A has been implicated in pathways of fibrosis in fibroblasts (295). IL-11 acts both in paracrine and autocrine signaling and has downstream effects on nearby cells. The proto-myofibroblast phenotype is not shown for the purpose of simplicity. Open arrows signify stimulation/enhancement while closed arrows signify suppression. IL-interleukin, TGF-β1 – transforming growth factor beta-1, α-SMA-alpha smooth muscle actin. Figure created with BioRender.com.

While this paper presents a conceptual framework for elucidating fibromyalgia-type manifestations of LC, it has not been empirically tested. Figure 6 below illustrates how several mechanisms may possibly contribute to persistent symptoms - functional and non-functional - after SARS-CoV-2 infection, and its infection-associated organ damage, e.g., pulmonary fibrosis, renal injury, myocardial injury, neuroinflammation, etc. The plausible contributing mechanisms may include immune cell dysregulation and autoimmunity, persistence of viral particle shedding in peripheral tissue, latent neurotropic pathogen reactivation, vagus nerve dysfunction or autonomic nervous system neuroinflammation, endothelial damage, hypercoagulability, muscle atrophy, immune-mediated myopathy, vascular disruption in the blood–brain barrier, and other abnormalities (1,7,12,426,427) including, possibly, a connective tissue abnormality that involves the myofascioskeletal bio-tensegrity-like system. These abovementioned suggested mechanisms are not necessarily mutually exclusive.

**Figure 6.**
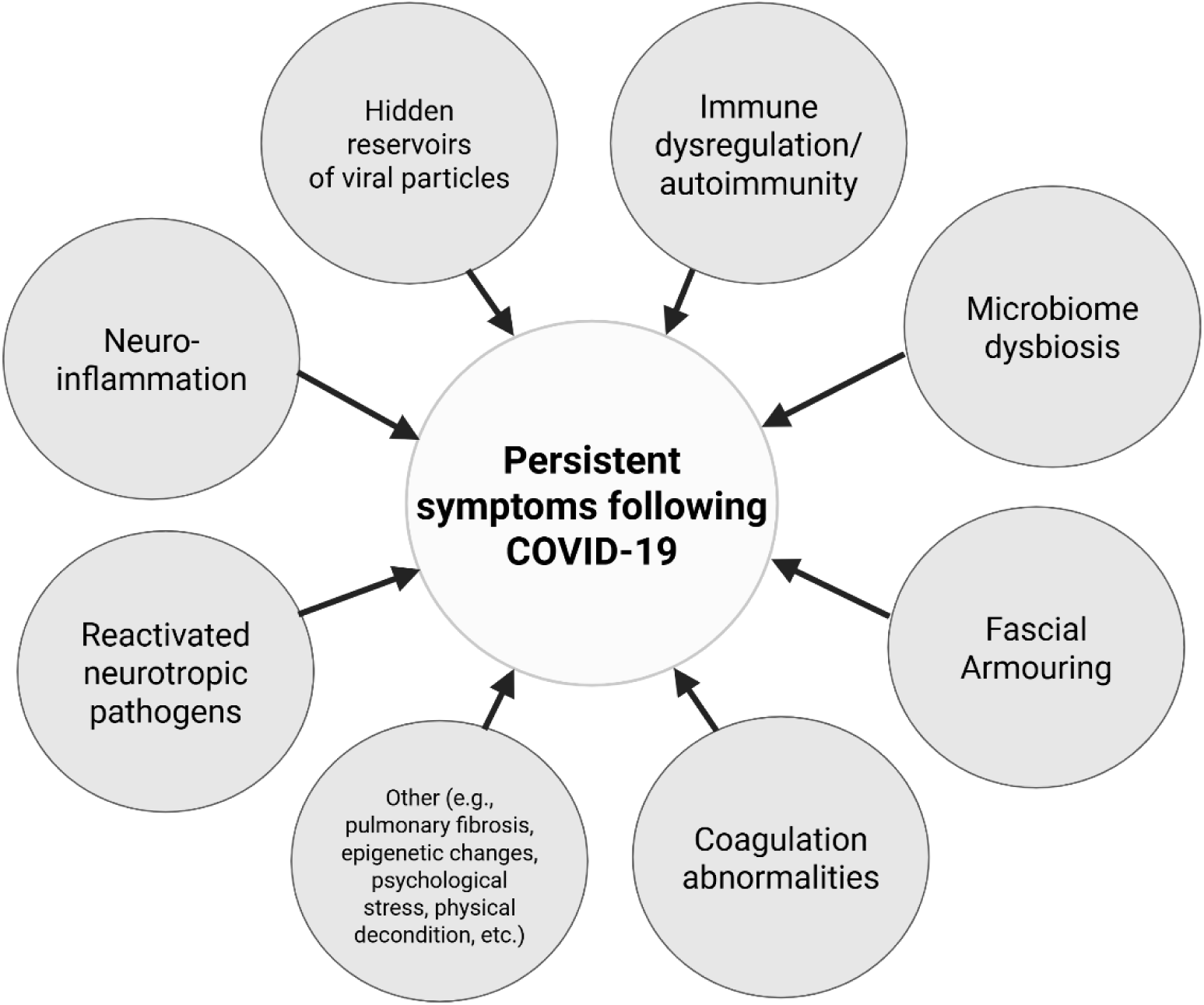
Persistent symptoms after COVID-19 have a multifaceted pathogenesis that may result from multiple cellular processes and organ systems involving various abnormalities. Even though lung fibrosis is noted, the term “long COVID-19 syndrome” should be reserved to multisystem medically unexplained symptoms after COVID-19 with no demonstrated well-defined organic damage to explain them, with no alternative organic diagnosis for them and no comorbidity to account for them, of the kind that is typically dismissed or labeled medically as “psychosomatic”. This syndrome should not be conflated with persistent symptoms after hospitalization or known lung or cardiac damage nor with post-intensive care syndrome, which are not the focus of this paper. For practical purposes, individuals who have pulmonary fibrosis after COVID-19 should be labeled as having “pulmonary fibrosis” (which can be covid-19 induced and which has its etiopathogenesis) rather than “LC syndrome”. Myocarditis induced cardiomyopathy, likewise, should be designated as “myocarditis induced cardiomyopathy.” Figure created with BioRender.com.

## 7. Interpreting LC manifestations and drawing theory-based predictions

In 2020 the World Health Organization declared covid-19, caused by sars-cov-2, a global pandemic (428). Approximately 10-30 percent of individuals who had covid-19 experience persistent bothersome symptoms after recovery from the acute phase (2,7,226,429) in what is officially termed “long covid” or post-acute covid-syndrome. While the social and economic burden of LC is a matter still being figured (430), many individuals with LC experience heavy symptom burden and a persistent medically unexplained multisymptom illness, termed by some scholars as “functional long covid” as opposed to an “organic long covid” (50). The observed similarity between LC and psychosomatic syndromes such as fibromyalgia and ME/CFS has led researchers to suspect a shared underlying mechanism (9,10,16,21,51,55,139,146). Some experts agree that LC should be defined as a psychosomatic disorder (30). Nevertheless, attributing the condition to deconditioning or labeling patients with normal results on medical investigations as “functional long COVID-19”, as opposed to an “organic long COVID”, is problematic.

Although it is true that improving respiratory muscle function can alleviate symptoms in cases where muscle atrophy and deconditioning are contributing to LC symptoms (431), attributing the condition solely to deconditioning and psychological factors fails to comprehensively address its complexity (432). Enck & Mazurak (2018) (433) emphasize that biopsychosocial models should not neglect organic biological aspects, even when applying them to somatoform-type disorders.

To date, controlled trials and large-scale cohort studies have not shown that current pharmaceutical therapies effectively reduce symptoms or improve radiological and biomarker profiles in LC (432), and there is an urgent need for effective treatments (226,434). While existing theories offer some insights, they often fall short in fully explaining the broad symptomatology of fibromyalgia. This underscores the imperative to explore novel mechanistic frameworks that can provide a more integrated explanation. Understanding the patho-mechanisms involved in LC may potentially lead to the development of better treatments. The following sections explain predictions that are derived from the mechanism suggested here for the pathogenesis of fibromyalgia syndrome.

### 7.1. Mechanistic Predictions of ‘Long COVID-19’ Manifestations Based on the Suggested Biomechanical Model

From the suggested mechanism of myofibroblast-generated tensegrity tension and ECM alterations in myofascial tissue, predictions regarding LC may be drawn, based on a mechanistic analysis. Any factor that enhances the cycle of myofibroblast mechanobiology and contributes to myofibroblast activity, theoretically leads to more cell contractility (and inherent reactive feedback regulation), and may advance the disorder, while factors that inhibit this cycle are generally expected to be protective factors. Based on the model, predictions can be made, explained as follows:

Risk/protective factors and relieving factors:

- Hypermobility syndrome/Ehlers-Danlos syndrome: Collagen microarchitecture affects mechanosensitive signaling in cells followed by an induction of myofibroblasts and secretion of proangiogenic factors (vascular endothelial growth factor and IL-8) when studied in human adipose-derived stem cell culture (435). Hypermobile Ehlers-Danlos syndrome is associated with ECM disarray and increased myofibroblast phenotype when studied in vitro (436). Hypermobile Ehlers-Danlos syndrome and hypermobility spectrum disorders are probably not separate entities but rather appear to be both on a continuum characterized by altered ECM homeostasis and a chronic inflammatory state (436). If ECM microarchitecture augments myofibroblast activity in patients with hypermobility syndrome, it is mainly for this reason that GJH is expected to increase their risk for fibromyalgia-type symptoms, a relationship that should be weakly explained statistically by psychological stress levels alone (290).
- Mechanical tension on the skin has been shown to enhance myofibroblast activity (437). The application of mechanical forces such as by use of splints further corroborates this finding (438). The development of myofascial pain is linked to tight-fitting clothes (290,439,440). Tight-fitting clothes and accessories are expected to predispose individuals to fibromyalgia due to input into the integrin-mediated yes-associated protein cascade of myofibroblast mechano-activity.
- Lifestyle and exercise (movement): Though not performed on myofascial tissue in vivo, a study showed that cyclical mechanical stretch reduces myofibroblast differentiation of primary lung fibroblasts (441). Tissue stretch reduces TGF-β1 and type-1 procollagen in mouse subcutaneous connective tissue (442). Immobility leads to fibrosis and an increase in myofibroblasts in knee joint capsule when studied in vivo (443). Immobility allows for the development of abnormal cross linking between connective tissue fibres (444). These findings provide, in general, the biological rationales for the role of exercise (movement) versus sedentarism according to the suggested myofascial-based mechanism. It is interesting in this respect that yoga involves cyclical stretching of almost all body parts as an integral aspect of the practice.
- The effect of taking hot showers during the acute induction phase of myofibroblasts following infection, and the long-term effect of a possible activation of heat shock proteins in a subcutaneous population of myofibroblast requires further investigation. It is not necessarily expected to be inert.
- The effect of weather changes on symptoms will be facilitated by the biophysical effects of temperature, electromagnetics and humidity on myofascial tissue and hyaluronic acid.
- If factors such as tattoo ink or smoking induce subcutaneous myofibroblasts, sedentary people with whole-body tattoos and smokers are expected to have worse and more prolonged psychosomatic symptoms after covid-19. A similar link is expected for those using cosmetics and topical creams containing substances that upregulate pathways of subcutaneous fibroblast-to-myofibroblast differentiation.
- Environmental factors such as pollutants, chemicals, and microbes can trigger or protect from fibrosis (295,445).
- Diet and the gut brain axis (446) fit in the mechanism of LC from multiple angles and not only in relation to connective tissue.
- Obesity: in obesity, connective-tissue fibrosis is induced and mediated by mechano-transducing signaling pathways (447). Fibrotic processes mediated by myofibroblasts can transform the mechanical properties of subcutaneous tissue, increasing its rigidity and connective tissue stiffness (447). For this reason, obesity is predicted to be a significant risk factor for LC.

#### Other manifestations based on the model

- <u>Hair loss:</u> adipocyte to myofibroblast transition is a possible cause of alopecia (448). The connective tissue sheath and follicular papilla can use gap junctions to form a communicating network. During hair cycling, this network plays a part in the control of hair follicle dynamic structural changes (449). Hair loss is therefore expected to occur and be weakly explained by psychological stress levels.
- <u>Pallor</u>: might be an overlooked manifestation, reflecting impaired peripheral perfusion due to autonomic and non-autonomic or hydrostatic causes.
- <u>Explaining Morning stiffness</u>: Tomasek et al. (2002) (291) describe the dynamics of fibroblast populations in three-dimensional collagen lattices and the process of generating traction and tension in their surrounding matrix of collagen fibrils. Over several hours the forces increase until a plateau is reached. If a similar process occurs in fascia in vivo, then a period of immobility would be comparable to this process of allowing the cells to reach the plateau of a higher tension state uninterrupted.
- <u>(Fascio)Musculoskeletal</u>: altered pendulousness of the legs. If the physician is searching for an objective mechanism-based sign of the disease to test bedside, this might be a relatively good one.
- <u>Cardiovascular</u>: A mild chronic compartment-like syndrome affecting multiple muscles should, by a chronic contraction of skeletal muscles, impair perfusion and lymph flow and alter starling forces which could exacerbate pre-existing subclinical cardiovascular issues. The typical presentation would, by reasoning, include changes in blood pressure regulation, fatigue on exertion or after a heavy meal, palpitations, higher resting heart rate, cold feet and palms, sub/clinical impairment of sexual function, and absence/impairment of morning erection in males, due to impaired blood flow to various organs. Chronic compressive forces in the periorbital fascia would lead to subclinical reduced optic disc perfusion. Idiopathic fluid retention might also be derived mechanistically.
- <u>Active loci</u>: possibly due to mechanical stress on the muscle spindles as well as sympathetic overactivity. Tonic slow adapting receptors in nuclear chain fibers of the muscle spindle would activate gamma motoneurons via the stretch reflex in prestressed myofascial tissue. Also, afferent input from gastrocnemius-soleus muscle C-fibres produces long-lasting excitability of the biceps femoris/semitendinosus α-motoneuron efferent fibers through the flexion reflex in an animal model (450). A mechanistic discussion of myofascial pain syndrome and active loci in the context of this framework is available in a recent study (290).
- <u>Immune system:</u> based on a finding that substrate stiffness affects immune cell function (398). Fibroblasts and inflammatory myofibroblasts secrete cytokines as part of their natural activity (295). An overactive (or “irritable”) state of immune cells due to paracrine proinflammatory cytokine secretion, chronic low-grade inflammation, and increased substrate rigidity of the ECM would likely predispose the immune system to over-reacting in intolerance to “irritant” antigens. Such immune hyperirritability could be evident in the form of predisposition to gluten intolerance, multiple chemical sensitivity, association with autoinflammatory reactions, or other clinical or subclinical immune dysregulation. Fibromyalgia and LC are associated with mast cell dysregulation (451). A mechanistic explanation, among several, can be related to findings (452) that tissue stiffness affects mast cell behavior and function.
- <u>Metabolism:</u> Myofibroblasts secrete IL-6, IL-8, and IL-11 (295,401). The cytokine IL-6, besides its effect on CD4+ T lymphocytes, can activate indoleamine 2,3-dioxygenase, as shown in different cell types (84,453,454), and therefore is potentially intimately related to the metabolic balance of the tryptophan-indoleamine 2,3-dioxygenase 1-kynurenine and serotonin pathway. Metabolites of this pathway (e.g., the neurotoxic metabolite quinolinic acid) (455), some of which can cross the blood brain barrier (456), were observed in altered systemic levels in fibromyalgia (457,458), and are linked to cognitive impairment and depression (84). Besides cytokines, the gut microbiome has the capacity to modulate indoleamine 2,3-dioxygenase 1 too, for example via butyrate production (84,459).
- <u>Mood and psychosomatic disorders:</u> post-traumatic stress disorder, anxiety, and depression are known manifestations of ‘long COVID-19’ (460). “Post-traumatic stress disorder” in this framework (not only in the context of LC) is expected to have a bio-mechanical aspect involving the (fascio)musculoskeletal system. Any acute sympathetic or inflammatory reaction which leads to a simultaneous abrupt contraction of multiple muscles and of the osteomyofascial tensegrity structure would cause a sudden shift in its biomechanical and energetic elastic state. The energetic shift and the mechanical tension locked in the ECM by contracting cells would lead to an increase in widespread tension in the body irrespective of alpha motoneurons. Sympathetic nerve fibers embedded in fascia would also be affected, which is a relevant interface with emotion and cognition. If the musculoskeletal tension is not released after this acute event, overtime fascia and ECM will be remodeled in this higher-tension state which initially was supposed to be a temporary sympathetic defensive reaction. This is followed by myo/fibroblasts remodeling the ECM and stress shielding themselves to mask the tension while, importantly, they form “supermature” focal adhesions and upregulated expression of α-SMA. In their resolution phase, the balance of proliferative and apoptotic signals is crucial for the outcome of myofibroblast cells (296). They can either undergo apoptosis (mediated by fibroblast growth factor 1, prostaglandin E2, and IL-1beta), evade apoptosis and persist in the tissue, or enter senescence (mediated by CCN1 with upregulated intracellular p16 and p21, and characterized by the acquisition of a senescence-associated secretory phenotype, specifically the secretion of TGF-β1 and pro-inflammatory cytokines and chemokines such as IL-6, CCL2, IL-1α, IL-1β, IL-8, PDGF, and ECM proteins), or other possible fates (296). Myofibroblasts become much more active above a certain threshold of matrix rigidity (461). Higher ECM pre-stress in the tensegrity-like structure crosses the threshold for myofibroblast activity and propels their cascade of mechanobiology and stress shielding, but once fascia is remodeled this way, it is much more difficult to resolve. Fibromyalgia does not typically erupt in patients overnight. The systemic implications aren’t limited to myofascial tissue, and include changes in metabolism and secretory profile of myofascial cells, changes in vasculature, effects on the immune system, and more. Interestingly, circulating systemic fibroblast growth factors can deeply affect brain physiology (462). Also, the intracranial ECM is suggested to be implicated in the pathophysiology of stress-induced depression (463).
- <u>Overlap with “myofascial pain syndromes”</u>: The clinical overlap of myofascial pain and associated psychosomatic and “non-specific” pain conditions (or “central sensitizations symptoms”) is likely to be evident in relation to LC. Figure 7 illustrates in general the clinical overlap reflected by the mechanistic overlap, as suggested by this conceptual framework (not all relationships are depicted in this scheme).

### 7.2. Predictions of results on investigations and means for testing the hypotheses

The myofibroblast-based model can be tested by several different methods.

- A preliminary non-invasive straightforward approach would be to measure muscle damping (350) which should reflect increased muscle tension. Pendulousness of the legs or arms of fibromyalgia-type LC patients compared to controls could be a relatively simple clinical test to start with, after controlling for age, sex, and body mass index. The inclusion of subjects would focus more on persistent “fibromyalgia-ness” patients who had mild acute infection, rather than dyspneic patients who had severe acute pulmonary covid-19. Shear wave elastography/strain elastography or magnetic resonance elastography can help measure the stiffness of tissue for comparing fibromyalgia-like LC patients and healthy controls. Again-focusing on those with multiple multiorgan psychosomatic complaints after mild/asymptomatic infection and excluding subjects who were hospitalized during the acute phases. The resonance of tissue and its response to internal organ oscillations might also be found to be altered.
- Sub/clinical decreased joint range of motion should be seen on careful examination when taking into consideration hypermobility syndrome. Demonstrating an inappropriately normal joint range of motion in a hypermobile individual is false-normal and pathological.
- Biopsies can be used to measure myofibroblast density, or to examine if fascial cells express elevated levels of α-SMA. But since myofibroblast can de-differentiate and leave behind a remodeled dysfunctional fascia, testing only by this method might actually be deceptive. Needle biopsy may be sufficient for this (464). Smokers are expected to have a higher density of myofibroblasts in myofascial tissue compared to non-smokers.
- Overall, pharmacological agents that enhance myofascial fibroblast-to-myofibroblast transdifferentiating are expected to predispose to LC and fibromyalgia.
- Microdialysis of muscle, for example of the trapezius muscle (368) or vastus lateralis (340)-not all patients necessarily have increased concentrations of algesic substances and signs of anerobic metabolism in the same muscles because not all patients necessarily have the trapezius (or vastus lateralis) deeply affected. The clinical variability is derived from a mechanistic variability regarding which anatomical structures and layers are more involved. Laser doppler fluxmetry and isotope washout methods can also be used (465).
- Quality of radial pulse on palpation could be a fairly useful clinical sign for the condition.
- Severe cases might have abnormally elevated serum and urinary creatine as a result of muscle breakdown and oxidative stress, if muscle cells fail to compensate.
- Increased physiological response to the Valsalva maneuver and exertional dizziness would be characteristic of a mild-to-moderate global chronic compartment-like syndrome.
- Activation of the stretch reflex due to diffuse involvement of the muscle spindles or tendon would manifest as increased muscle tone not mediated by alpha motoneurons.
- Biophysical tests-strain elastography, atomic force microscopy, optical coherence elastography, dynamic mechanical analysis, etc., of fascial/myofascial tissue might be insightful, although these would have to take into account the complexity of the model and possible confounding factors. Age, sex, pH, temperature, hydration, hyaluronic acid composition, elastin and collagen polymorphisms, adipocytes, cell phenotype and density, are all variables that may affect the properties of fascia in vivo.
- Heterogenous clinical complaints are derived from the mechanistic variability. When the transformation of fascial ECM reinforces the cycle of myofibroblast force generation, myofascial degree of stiffness increases and muscles are then subjected to low-grade chronic ischaemia. Over time, in the absence of full muscle relaxation and due to insufficient nutrients and oxygen, muscle mass and muscle cells experiencing longstanding low-grade hypoxia will undergo long term structural, metabolic, and genetic/epigenetic level adaptations and compensations, while the immune system is continuously drawn into the process due to ongoing tissue injury. Sedentarism reinforces atrophy of skeletal muscle. Afterwards, matrix material such as collagen replaces atrophic skeletal muscle mass and at this stage fatigue and weakness become more prominent. Meanwhile, fascia can either continue in its positive feedback, or break the cycle and proceed to a stress-relaxation failure stage, where it experiences mechanical creep and has lower shear modulus. A higher myofascial Young’s modulus is expected if pain, tension, and stiffness are the main complaint, and lower fascial stiffness is expected if fatigue, weakness, and pain are the predominant complaint.

**Figure 7.**
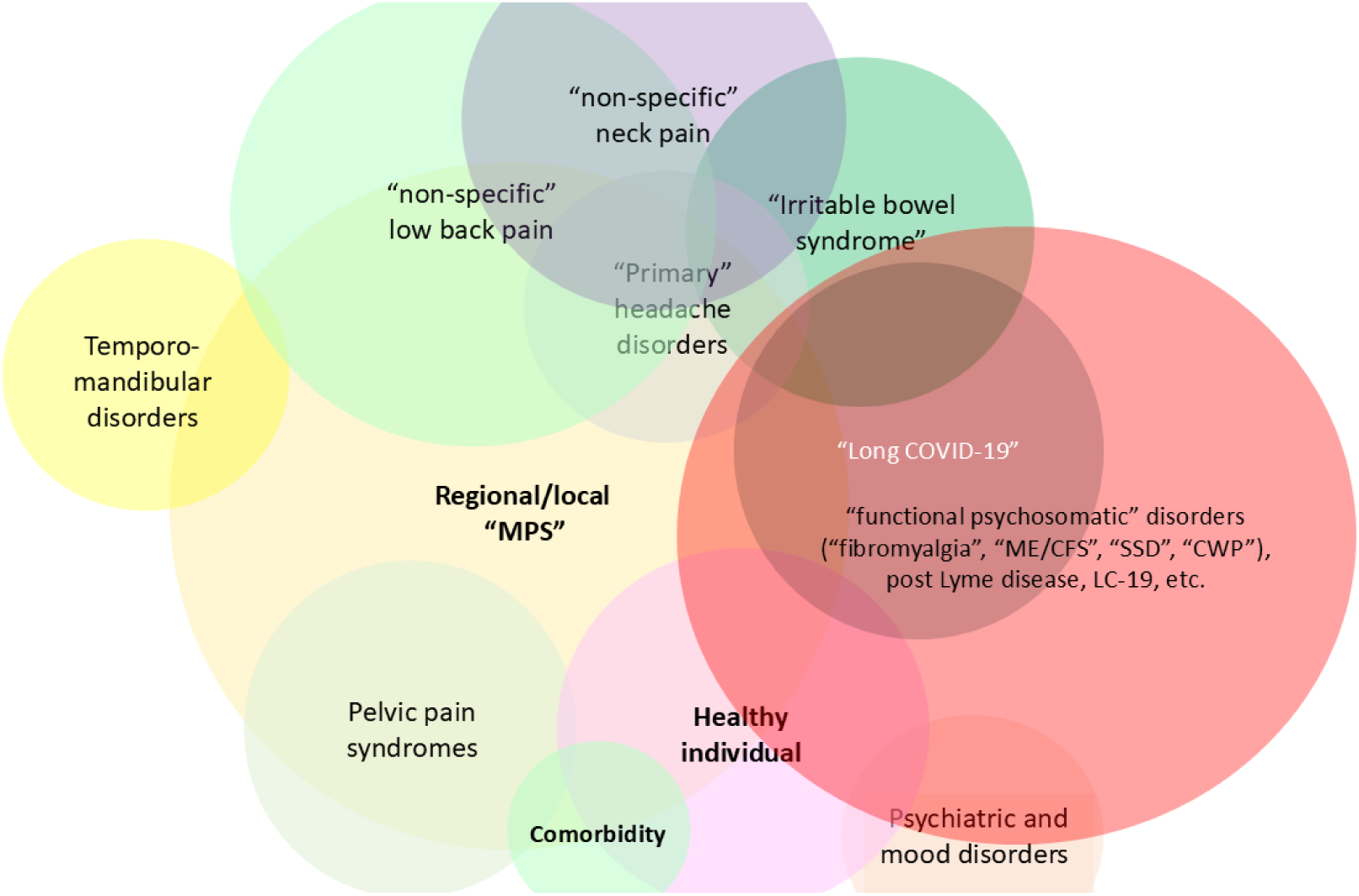
Overlaps of psychosomatic fibromyalgia-type conditions. The syndrome of long covid-19 refers here to ME/CFS-fibromyalgia-type persistent clinically unexplained symptoms. The clinical manifestation de facto depends on multiple factors including environmental, genetic, behavioral, psychological, iatrogenic, and which anatomical structures are more involved etc. Individuals with known organ damage after severe hospitalized covid-19 or those with lung parenchymal scarring after infection or those hospitalized with hypoxia are not the focus of this illustration. Involvement of the abdominal fascia and intestines navigates the abnormality more towards a functional gut disorder or irritable bowel-like syndrome. Involvement of the pelvic fascia navigates more towards genitourinary symptoms. Masseter and temporalis muscle fatigue during mastication would be associated with temporomandibular involvement. Involvement of the lumbar and abdominal region navigates more towards “non-specific” low back pain, and so forth. In clinical terms, “SSD” and “CWP” can be said to be a prodrome of fibromyalgia. Not all relationships are depicted in this diagram. Colors are for visual clarity and have no special meaning. The term “healthy individual” is subject to the reader’s interpretation. CWP-chronic widespread pain. LC-long covid-19. ME/CFS-myalgic encephalomyelitis/chronic fatigue syndrome. MPS-myofascial pain syndrome. SSD-somatic symptom disorder.

Cadaveric studies might not be the best choice to investigate the tensegrity dynamics. Mathematical computer models might be more useful in this case.

## 8. Conclusions

Long COVID-19 is increasingly becoming a public health concern and, similarly to fibromyalgia, still lacks a comprehensive biology-based definition, diagnosis, staging, epidemiology, pathophysiology, prognosis, prevention, and treatment. The term “LC syndrome” and its synonyms should be reserved for patients with a persistent clinical picture that does not fit other known organic diseases, when no well-defined organ damage is found to account for the patient’s clinical picture. Several plausible mechanisms have been hypothesized in literature so far to explain LC such as direct viral toxicity, dysregulation of the immune system, persistence of viral particles in peripheral tissue, latent viral reactivation, neuroinflammation, microbiome alterations, an imbalance of serotonin in the brain, vascular disruption in the blood–brain barrier, brainstem dysfunction, dysregulated circadian rhythms, microvascular endothelial damage, vascular thrombosis, deranged endocrine functions, tissue infiltration of amyloid-containing deposits, epigenetic changes, exercise-induced myopathy, sarcopenia and physical deconditioning, and more (1,7,12,426,427,429,466). Here, a different perspective is proposed for the pathophysiology of LC as a disorder of immuno-rheumo-psycho-neurology involving the (fascio)musculoskeletal system and the cascade of myofibroblast force generation and ECM remodeling in soft tissue.

The suggested neuro-mechanobiological physiological model predicts a link between LC and myofascial proto/myofibroblast phenotype cells and is also used to make testable experimental predictions on investigations, and it predicts risk and relieving factors in LC, as well as effectiveness of treatment options. Still, this pathophysiological model clearly needs to be adjusted according to empirical studies.

Physicians should be careful not to assume by default that a patient presenting with unexplained chronic pain and multiorgan medically unexplained symptoms is malingering or over exaggerating as this is a major barrier to treatment for fibromyalgia patients in many clinical settings and can easily become a similar obstacle for long covid patients. The conceptual framework of “fascial armouring” links between unhealthy lifestyle and pain and suffering. As with many other medical scenarios, it is multifactorial. An unbalanced body will hold an unbalanced mind. Holistic therapies targeting both body and mind are under-utilized in the treatment of chronic psychosomatic pain conditions. Biology does not separate or segregate itself into distinct medical specialties as we do in our profession, and the “body” and the “mind” are one being, one flesh. Further research is needed to better understand the post-acute sequala of covid-19 and how to best manage new onset psychosomatic symptoms in patients recovering from coronavirus infection.

## 9. Limitations

The main limitation of this scoping review is that a sole researcher evaluated the studies. Being a new field, the review aimed to report the extent and range of studies on the subject. Much less was the focus to appraise the quality of the studies (in agreement with Peters et al., (2015) (467)), which when done by a sole author can introduce biases. The exclusion of studies published in journals ranked Q4 according to JCR aimed to give a fair balance between quality and quantity, implementing an objective criterion. As a consequence, some literature was not covered. Another drawback could be that the H-index and other parameters weren’t taken into consideration. A rounded average of the quartile in the three years adjacent to the publication time might be a more reflective marker of a journal’s quality. Two major databases, Web of Science and Medline, were searched systematically, and the search was limited to the period of inception to end of the year 2024. Cochrane library was not used, which is another weakness. Keywords focused on fibromyalgia and covid, not chronic fatigue syndrome despite a notable overlap between the two syndromes. As stated, the review focused on investigating fibromyalgia-type symptomatology, and the issue of fatigue was not expressed in the systematic search. Fatigue after covid can occur due to several reasons (physical deconditioning, muscle atrophy and sarcopenia, lung scarring, endothelial damage, genetic, metabolic, and mitochondrial level alterations, etc.). The terms “conditioned pain modulation” or “quantitative sensory testing” were not used in the systematic search phrases, and the term “medically unexplained” or “somatic symptoms” was not used either. To understand the mechanisms involved in LC better, future scoping reviews on this topic should have a broader scope and include terms related to somatic symptom disorder sand the general phenomenology of functional somatic syndromes. These are possibly phenotypes of a medical entity which putatively share a pathogenesis linking them mechanistically, as some researchers have suggested. Focusing this current review on pain narrowed the reviewed literature to primarily pain-related studies, though, being a scoping review, a specific narrow question was not asked. Rather the question of “what is the evidence on fibromyalgia manifestations post-covid as a phenomenon” was the aim of the review as a precursory step for future empirical research and systematic reviews. Excluding studies that recruited patients with a history of hospitalized covid-19 could be a disadvantage, but it was meant for minimizing confounding by organ damage-related symptoms or post-intensive care syndrome. Fibromyalgia-type manifestations were the focus, which are typically described as non-organic, functional, medically unexplained, or non-physiologic. Also, as the field is relatively new, valuable studies might have been missed if not published yet, and the preprint database search was not systematic and included only one preprint database (medRxiv). This is a drawback especially when reviewing a subject which is still new and emerging. Congress abstracts or proceedings have been likewise not purposefully searched. “Fibromyalgia features” were a criterion for inclusion of a study in the review but the validity of the definition as provided in the methods section is debatable. Also, the exclusion of studies investigating the effects of the covid pandemic on fibromyalgia symptoms in patients that were already diagnosed with fibromyalgia prior to the pandemic, likely omitted potentially relevant data. The mechanism of exacerbation of fibromyalgia might reasonably be linked to the pathogenesis of the disease, especially when considering contemporary medical community’s common belief that emotional stress triggers fibromyalgia. Due to the overlap between myofascial pain syndrome and fibromyalgia (in terms of trigger points and tender spots etc.) the term myofascial pain was part of the search, but not the term “musculoskeletal pain” (or “pain” in general). This was due to time and resource constraints. Consequently, relevant studies likely have been missed, though some were included because they were identified in the cited references of included studies. As central sensitization is the most accepted theory for the mechanism of fibromyalgia, this term was integrated into the search strategy but other putative theories for fibromyalgia were not. Several theories, all of which are still under dispute, have been suggested for fibromyalgia pathophysiology, though none of them are currently considered good enough to attribute the pathophysiology to entirely. Future research should take this into consideration.

## Data Availability

All data produced in the present study are available upon reasonable request to the authors

